# Inferring genetic variant networks by leveraging pleiotropy shows trait relationships drive massive pleiotropy in GWAS

**DOI:** 10.1101/2024.06.01.24308193

**Authors:** Martin Tournaire, Asma Nouira, Mario Favre Moiron, Yves Rozenholc, Marie Verbanck

## Abstract

Genetic variants have been associated with multiple traits through genome-wide association studies (GWASs), but pinpointing causal variants and their mechanisms remains challenging. Molecular phenotypes, such as eQTLs, are routinely used to interpret GWAS results. However, much concern has recently been raised about their weak overlap. Taking the opposite approach with PRISM (Pleiotropic Relationships to Infer the SNP Model), we leverage pleiotropy to pinpoint direct effects and build variant-trait networks. PRISM clusters variant-trait effects into confounder-mediated, trait-mediated, and direct effects, and builds individual variant networks by cross-referencing results from all traits. In simulations, PRISM demonstrated high precision in identifying direct effects and reconstructing variant-trait networks. Applying PRISM to a set of 70 complex traits and diseases representative of the phenome from the UK Biobank, we found that direct effects accounted for only ∼11% of significant effects, yet were highly enriched in heritability. Multiple lines of evidence showed that PRISM networks are consistent with established biological mechanisms.

## Introduction

Over the past 20 years, Genome-Wide Association Studies (GWASs), which test the association between several million genetic variants and a trait of interest, have established a myriad of associations between genetic variants and human complex traits^1–3^. According to GWAS catalog^4^, 7,714 publications and 1,142,122 unique genetic variant-trait associations have been reported as of June 2026. However, GWASs suffer from multiple limitations and biases. First, a major limitation is pleiotropy, *i.e.* a single genetic element affecting more than one trait, known to be pervasive throughout the human genome^5–7^. Because of pleiotropy, the same genetic variants are often associated with multiple traits in GWASs^8^. Second, it has been shown that most identified genetic variants are intergenic and involved in the regulation of the expression^9^. Consequently, the link between genetic variants and genes, and the underlying biological mechanisms, remain largely unresolved. A systematic review from 2022 reported only 309 experimentally validated non-coding GWAS variants^10^. Moreover, because of Linkage Disequilibrium (LD), referring to the non-random association of alleles at different loci on a chromosome, candidate causal variants in a genomic locus are indistinguishable^11^. Therefore, precisely pinpointing true causal genetic variants to the complex traits they affect proves to be a tremendous challenge^12^, hence the need for computational approaches complementary to GWASs.

Extensive follow-up analyses have become essential for prioritizing likely causal variants identified from GWASs^13,14^. These analyses typically include: 1) Pathway enrichment, which aims to identify biological pathways enriched with associated variants^15,16^. 2) Functional characterization, which aims to highlight the functional roles of GWAS variants^17,18^. 3) Fine-mapping, which aims to prioritize between correlated candidate variants^12,19–22^. However, all of these follow-up analyses heavily rely on genome annotations. And yet recently, much concern has been raised about the fact that GWAS variants overlap very little with annotations like molecular quantitative trait loci (QTL), particularly expression QTL (eQTL)^23,24^. To the point that GWAS and cis-eQTL variants show systematic structural differences^25^. Likewise, GWAS and EWAS (epigenome-wide association studies) do not find the same causal genes^26^.

Here, we take the opposite view to current methods, in order to completely sever our approach from annotations to further refine GWAS findings. Instead, the idea is to take advantage of the omnipresence of pleiotropy to disentangle variant-trait associations obtained from GWAS. We hypothesize that an associated locus from GWAS can result from 3 distinct underlying biological mechanisms: 1) *confounder* pleiotropy when the association can be explained by a confounder between traits; 2) *vertical* pleiotropy when the association can be explained by a variant-trait effect on another trait, which in turn affects the primary trait; 3) *direct effect* when the association is caused by a true unmediated direct effect from variant to trait. Therefore, to disentangle variant-trait associations, we propose to leverage pleiotropic relationships between traits by rerouting an integrative Mendelian randomization (MR) method.

MR aims to infer the causality of an exposure trait *X* on an outcome trait *Y*. MR uses genetic variants as instrumental variables that are robustly associated with the exposure of interest and tests whether the effects of the variants on the exposure result in proportional effects on the outcome. Classical MR relies on three assumptions: 1) the genetic variants must be strongly associated with the exposure *X*; 2) the genetic variants cannot directly affect the outcome *Y*; 3) the genetic variants must not affect the confounder *U* of the exposure-outcome relationship. It has been shown that traditional MR massively suffers from pleiotropy, which violates the assumptions and biases the results of MR^27^. This led to the development of multiple integrative MR methods that take into account pleiotropy, notably by modelling a latent confounder (*e.g.* LHC-MR^28^, CAUSE^29^, MR-CUE^30^) or by clustering genetic variants (PWC-MR^31^, MR-Clust^32^). However, MR only uses genetic variants as instrumental variables, and no conclusions are drawn *a posteriori* on individual variant-trait associations. Even MR methods designed to handle pleiotropy predominantly treat variant pleiotropy as an issue to eliminate rather than a phenomenon worth leveraging. Therefore, we chose to reroute MR to explicitly focus on the relationships between genetic variants and traits.

Here, we propose PRISM, which stands for Pleiotropic Relationships to Infer the SNP Model, a genome-wide method to disentangle variant-trait associations from GWASs, into confounder, vertical, or direct effects. The rationale behind PRISM is to re-examine a given variant-trait association from GWAS through the prism of all other traits. Concretely, PRISM runs an MR model between a given trait of interest and all other traits, while including pairwise confounders, to capture different pleiotropic trait contexts. Then, by aggregating the results, PRISM predicts significant variant-trait effects that are consistent regardless of the pleiotropic context. Finally, based on relationships between traits, PRISM assigns labels to all significantly consistent variant-trait effects and constructs a network for each variant. To assess the performances of PRISM, we simulated GWAS summary statistics for a complex network of traits. Then we processed a set of 70 representative heritable traits from the UK Biobank^33^ spanning 12 broad phenome categories through PRISM and disentangled the effects of ∼4 million variants to infer variant networks.

## Results

### PRISM disentangles the associations of genetic variants from GWASs by leveraging pleiotropy

We hypothesize that the observed associations in GWAS can be attributed to 3 distinct underlying biological mechanisms: 1) *confounder pleiotropy* which occurs when the variant has an effect on the confounder of the relationship between traits; 2) *vertical pleiotropy* which arises from the variant-trait effect caused by another trait; 3) *direct effect* which occurs when the association is due to a direct effect from the variant on the trait (Fig. 1A). In other words, variant-trait associations in GWASs are induced by direct effects and their pleiotropic ripple effects. It is worth mentioning that we can also define “horizontal pleiotropy” which occurs when a variant has several distinct direct effects on more than one trait. To comprehensively disentangle these associations, PRISM analyzes every single variant-trait effect across multiple pleiotropic contexts using GWAS summary statistics for multiple traits (Fig. 1B). Specifically, the effect of a given variant on trait *X* is reassessed by jointly modeling the effect of the variant on trait *X* with each one of the other traits (*A*, *B*, *C* and *D* in Fig. 1B). By aggregating information through the prism of all other traits, PRISM predicts whether the variant-trait effect on *X* is consistent across the different contexts, and subsequently clusters the effect as null, pleiotropic, or direct. Concretely, the “confounder pleiotropy” label indicates that the variant was identified as having an effect on trait *X* only through a confounder shared with another trait. In contrast, the “vertical pleiotropy” label denotes that the variant effect on trait *X* is only mediated through another trait causally related to *X*. Conversely, a “direct effect” conveys a significant effect that was not mediated by pleiotropy. Once this procedure is completed for all traits, the obtained direct and pleiotropic labels are integrated to construct a network for each variant and the traits it impacts.

**Figure 1:**
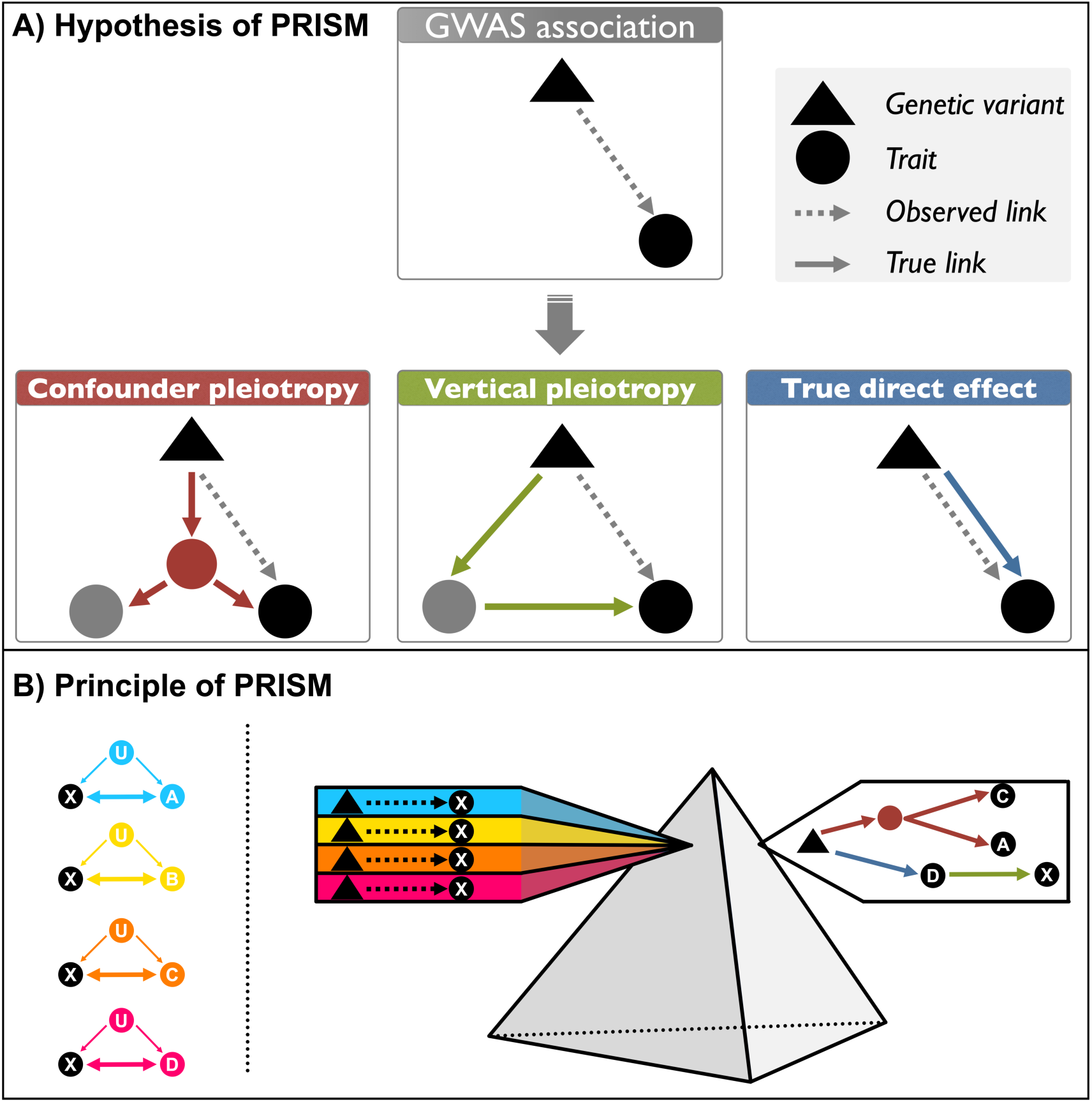
Overview of PRISM | **A)** 3 distinct mechanisms underlying GWAS associations: 1) confounder pleiotropy, i.e. effect mediated by a confounder; 2) vertical pleiotropy, i.e. effect mediated by a causal relationship between two traits; 3) direct effect. **B)** PRISM principle. 1) Each variant-trait association is evaluated in a model of trait X in different contexts relative to trait A, then to trait B, and so on. Each model takes into account a unique latent confounder U (left panel and Supplementary Fig. 1). 2) PRISM assesses the effects of all studied genetic variants on a trait X, through the prism of multiple trait contexts. Then, by cross-referencing the information and combining all traits, a full pleiotropy network can be built for each variant (right panel).

### PRISM accurately distinguishes direct from pleiotropic effects in simulations

We constructed a pleiotropic network consisting of 60 simulated traits organized into four subnetworks of 15 traits with a similar structure (See Fig. 6). Briefly, each simulated subnetwork consisted of different groups of traits *A*, *B*, *C*, *D* and *E*. *A* connected directly to *B*_3_ and *B*_4_, while *B*_1_ and *B*_2_ had mutual causal influences and *D* was causal to *A*. *B*_4_ also connected to traits from the *C* group, which made *B*_4_ the most pivotal trait in the subnetwork. Traits from group *E* had no causal relationships with any other traits. All traits shared a unique confounder with every other trait in the subnetwork. Moreover, to link the subnetworks, a confounder was simulated between each pair of *B*_4_ traits across subnetworks. This global structure emphasized the complex interplay of direct, vertical, and confounder relationships among traits. In total, the simulated network was composed of 60 traits, and 426 confounders. 100,000 genetic variants were simulated for each of the 60 traits generating GWAS summary statistics.

In addition, this network encompassed a wide array of scenarios consisting in modulating the polygenicity and the heritability of traits, the strength of causal effects between traits, and the strength of the confounders (Supplementary Table 1). Then, we processed the simulated data through PRISM, and compared the true variant-trait effects with those predicted by PRISM. We calculated precision and recall to assess the performance of predicting each type of effect using two complementary approaches (See Methods). In the first approach, referred to as *all effects*, a prediction was considered correct if it matched any of the potentially overlapping true effects. This approach was used to evaluate whether PRISM made genuine classification errors from a statistical standpoint, but is not as informative for interpretation. In the second approach, referred to as *prioritized effects*, each true variant-trait effect was assigned a single label according to a predefined hierarchy prioritizing the most interpretable effect from a biological standpoint: 1) direct, 2) vertical, 3) confounder. Under this more stringent approach, a prediction was considered incorrect if it did not match the prioritized true label, even when it corresponded to another overlapping true effect. This evaluation is therefore more relevant for interpreting real data.

First, as shown in Fig. 2 (*i.e.* traits from groups *A*, *B*, *C*, *D*, and high causal effects between traits), Supplementary Fig. 2 (*i.e.* traits from groups *A*, *B*, *C*, *D*, and low causal effects between traits) and Supplementary Fig. 3 (*i.e.* traits from group *E*), PRISM consistently achieved high precision in detecting direct effects in all scenarios. When considering the *all effects* approach, precision was also high for vertical pleiotropy and confounder pleiotropy, indicating that PRISM correctly predicted all effects. When considering the *prioritized effects*, precision remained high for direct effects in all scenarios. Precision for vertical pleiotropy was also high in low and medium polygenicity, but decreased with high polygenicity, where individual effects were weaker and overlapping pleiotropic effects were harder to disentangle. Confounder effects were the most delicate to classify, especially in the case of low and pervasive confounders, since these effects were very weak. Regarding recall, PRISM recovered direct effects well, but recall decreased with increasing polygenicity, as expected: since the total trait heritability was fixed, when polygenicity increased, individual variant effects were smaller. The same trend was observed for vertical effects. Recall for confounder effects was lower overall, particularly for low and pervasive confounders and at higher polygenicity. Overall, we prioritized precision over recall, ensuring that PRISM stringently controls for false positives. As shown in Supplementary Figs. 4-6, these results are robust to lower trait heritability. Importantly, by design, PRISM prioritizes the capture of direct effects and their ripple vertical effects. Consequently, variants tend to be classified as confounders when their pleiotropy cannot be clearly disentangled, which explains the comparatively lower precision to detect confounder effects, which are the least interpretable effects. Having assessed genuine false-positive classifications with the *all effects* evaluation, the *prioritized effects* approach will be used in the following analyses, as it provides a more interpretable framework to discuss simulation results and their implications for real data.

**Figure 2:**
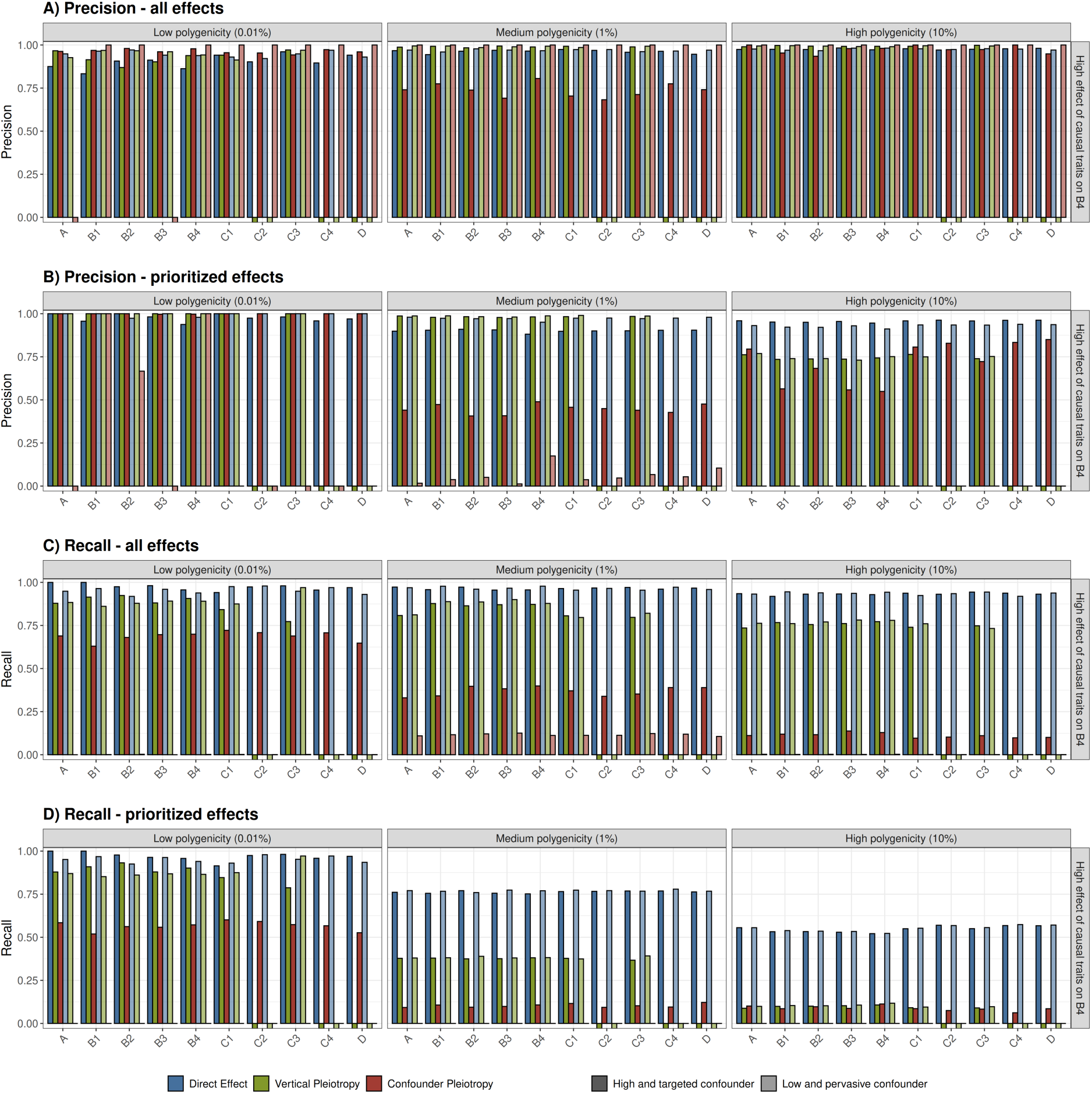
**A) B)** Precision and **C) D)** recall of PRISM predictions for significant variant-trait effects, on simulations. **A)** and **C)** represent the performance of PRISM when multiple predicted and true labels are allowed (i.e. all effects, see Methods). **B)** and **D)** represent the performance of PRISM when each true variant-trait effect is assigned a single prioritized label for interpretation (i.e. prioritized effects, see Methods). The x-axis represents the simulated traits, grouped across subnetworks for visualization purpose (See Fig. 6 and Methods). Significant effects are defined with *P* < 5 × 10^-8^/59, the recommended threshold from PRISM (See Methods). Bars are colored according to predicted labels: direct effect (blue), vertical pleiotropy (green), or confounder pleiotropy (red). Six scenarios are represented across facets, with varying parameters of polygenicity and type of confounder. Polygenicity represents the proportion of variants with a direct effect on each trait. Causal effect on *B*_4_, the most pivotal trait in each subnetwork, represents the proportion of effect passed to each one of the four *B*_4_ traits, for all traits with a non-zero vertical effect on *B*_4_. High targeted confounder (darker shades) means that few variants (0.01%) have an effect on the confounder *U*, but with magnitude of effect rivaling direct effects. Low pervasive confounder (lighter shades) means that a large proportion (5%) of variants have an effect on the confounder, but with low magnitude. All traits are simulated with high heritability (60%). For precision, the y-axis represents the proportion of well-predicted variants among all predicted variants for a given label. For recall, the y-axis represents the proportion of well-predicted variants among all true variants for a given label. NB: In cases where no variant was clustered under a given label, it was impossible to calculate precision or recall; such instances are represented by squares below the x-axis. Results for low causal effect on *B*_4_ can be found in Supplementary Fig. 2. Results for traits *E*1-*E*5 can be found in Supplementary Fig. 3.

### PRISM accurately reconstructs variant networks in simulations

The end goal of PRISM is to use predicted and labeled variant-trait effects to construct a trait network for each variant. A network consists of nodes that represent the variant and the traits it significantly affects, and edges that represent the variant-trait and trait-trait effects labeled as direct or pleiotropic. In our simulations, we applied a weighted Simple Matching Coefficient (SMC) to individually compare the variant networks predicted by PRISM with their true simulated counterparts. First, we measured the accuracy of the predicted networks by matching their edges with those of the true networks. As illustrated in Supplementary Fig. 7, the accuracy of the predicted edges from PRISM was very high in all scenarios, therefore edges that are predicted by PRISM were either identical or very similar to those of the true networks. As shown in Supplementary Fig. 8, the power to predict edges decreased with increasing levels of polygenicity, and was particularly affected in case of a low and pervasive confounder, leading to incomplete networks when the method lacks sufficient power to detect all variant-trait effects. However, this is a deliberate methodological choice to stringently control for false positives. In other words, although the networks built by PRISM might be incomplete, the inferred edges are very likely to represent true effects.

### PRISM reassesses GWAS variant-trait associations and distinguishes between direct and pleiotropic effects

PRISM tests and labels variants for direct and pleiotropic (*i.e*. vertical and confounder) effects on a panel of traits. Contrary to GWAS, an effect is considered as PRISM significant if it remains consistent across multiple assessments in different pleiotropic contexts. We applied PRISM to GWAS summary statistics from the UK Biobank^33^. Our procedure for trait inclusion aimed to ensure that the traits span broad categories representing well the whole phenome, which led us to include 70 traits spanning 12 broad phenome categories (See Supplementary Table 2 and Methods for details). First, to illustrate how PRISM processed GWAS associations, let us take the concrete and representative example of coronary heart disease (CHD) (Supplementary Fig. 9). The majority of the 910 significant GWAS variants were classified as having vertical, confounder or null effects by PRISM, *i.e.* 97, 441 and 366 variants respectively. Only 6 variants were identified as having a direct effect, and all mapped to the non-coding gene CDKN2B-AS1. A recent study^34^ highlighted its potential role in CHD by acting as an RNA-sponge regulating microRNA miR-92a-3p, a known therapeutic target of CHD^35–37^. This explains why PRISM predicted a direct effect. Importantly, the vast majority of highly significant variants in the original GWAS for CHD were labeled as confounder effects. Finally, closely examining the networks of pleiotropic (vertical and confounder) variants revealed that most variant effects on CHD were mediated by lipids.

Generalizing to all 922,942 variant-trait effects predicted by PRISM across the selected 70 traits, we observed that direct effects represented only ∼11% of all PRISM significant effects, while vertical and confounder pleiotropy represented 47% and 42% respectively. Therefore, by reassessing GWAS associations through the prism of pleiotropy, PRISM offers novel insights into the biological mechanisms underlying associations observed in GWAS.

### PRISM reveals that most observed pleiotropic associations in GWASs are caused by relationships between traits

In GWAS, extensive pleiotropy, where a single genetic variant shows significant associations with two or more traits, is massively observed^38^. Since PRISM leverages pleiotropy, we aimed to characterize the pleiotropic nature of the associations discovered in GWASs. Specifically, we identified 187,981 variants exhibiting observed pleiotropic effects from GWASs in the included 70 traits from UK Biobank, and assessed the nature of this pleiotropy using PRISM. Naturally, confounder and vertical effects provide us with straightforward pleiotropic effects. To complement the interpretation of pleiotropic effects, we proposed to introduce an additional type of pleiotropy: horizontal pleiotropy which occurs when a genetic variant has a direct effect on at least two traits. Using PRISM, we found that confounder and vertical pleiotropies were responsible for respectively 65.2% and 34.6% of the observed pleiotropy across all traits (Fig. 3 and Supplementary Fig. 10). Whereas, horizontal pleiotropy, multiple direct effects from a genetic variant, was found extremely rare and represented 0.17%. Therefore, the extensive pleiotropy observed in GWAS may be largely attributed to the relationships among traits. This finding underscores the complexity of genetic effects on multiple traits and highlights the interest of PRISM in elucidating the underlying mechanisms of pleiotropic associations in GWAS findings.

**Figure 3:**
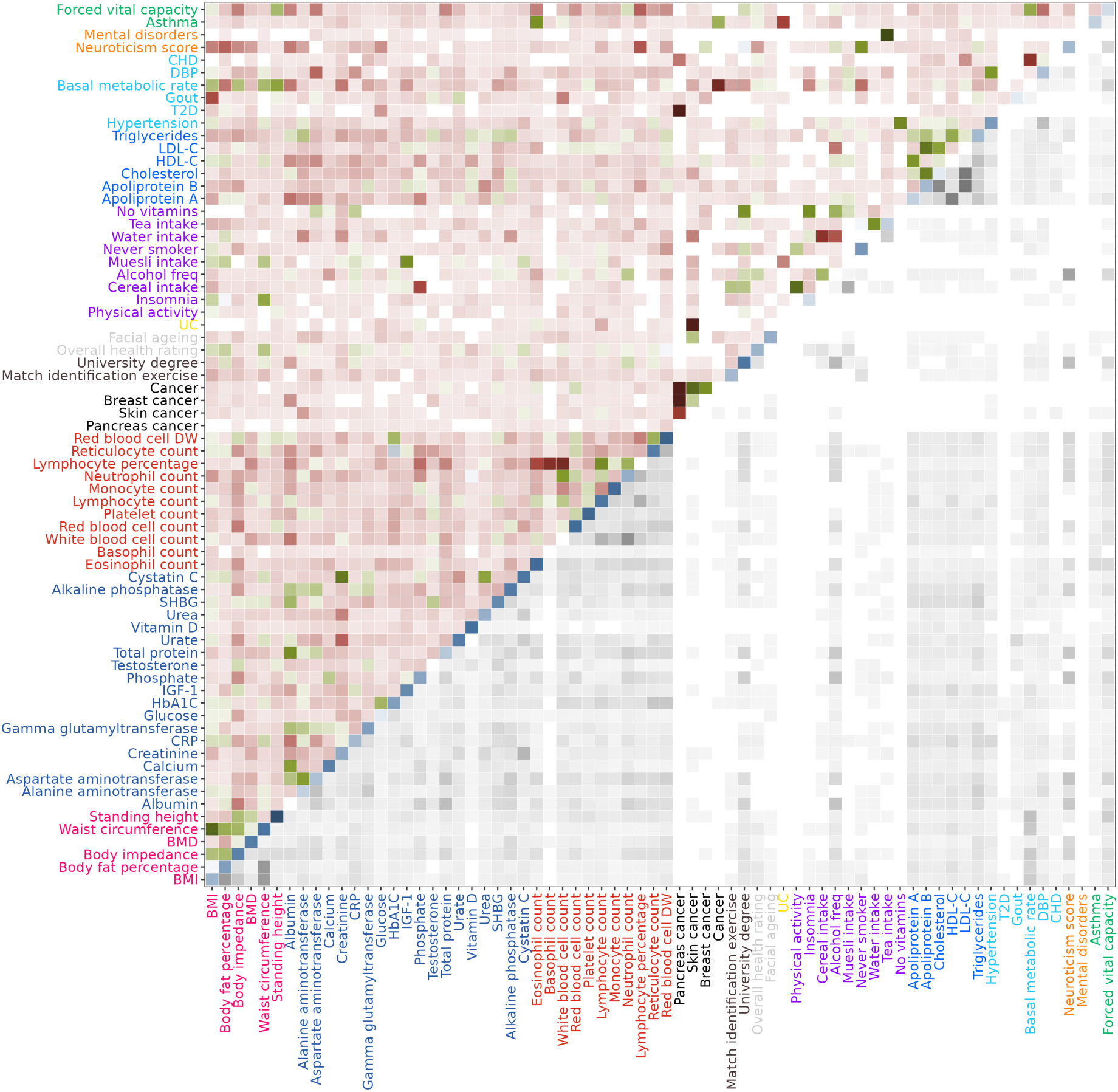
Heatmap of variant effects shared between traits. The bottom-right triangle represents the proportion of variants shared between traits in GWAS. The grey tile gradient represents the observed pleiotropy from GWAS (scaled proportions). The top-left triangle represents the largest proportion of PRISM significant variants classified in one of the three categories of pleiotropy between the pair of traits (confounder, vertical, or horizontal). The red, green and blue tile gradients represent confounder, vertical and horizontal pleiotropy respectively (scaled proportions). The diagonal represents the proportion of individual direct effects for each trait according to PRISM.

### PRISM direct variants are significantly enriched in per-variant heritability compared to GWAS variants and pleiotropic variants

In GWAS, variant heritability measures the proportion of trait variance explained by all measured variant-trait associations. Thus, the per-variant heritability is the contribution of a specific genetic variant to the overall heritability. We applied stratified LD score^39^ (s-LDSC) to estimate per-variant heritability enrichments across labeled variant-trait effects grouped into four categories: three categories for PRISM significant variant effects (direct, vertical, confounder), and a category for GWAS significant associations. Across most traits, we observed that PRISM direct variants demonstrated higher enrichment in per-variant heritability compared to both other PRISM categories (vertical and confounder variants) and to GWAS variants (Fig. 4). Specifically, for almost 80% of the traits, the enrichment was higher for PRISM direct variants than for GWAS variants, with a median fold increase of ∼1.7. As an example, for Apolipoprotein B, the enrichment for PRISM direct variants reached 127, compared to 36 for GWAS-significant variants, corresponding to a 3.5-fold increase. These findings support the hypothesis that indirect pleiotropic effects on traits are weaker compared to direct effects. Importantly, randomly shuffling PRISM labels to check for potential biases indeed eliminated the heritability enrichment for direct variants (See Supplementary Results), providing further support for these results.

**Figure 4:**
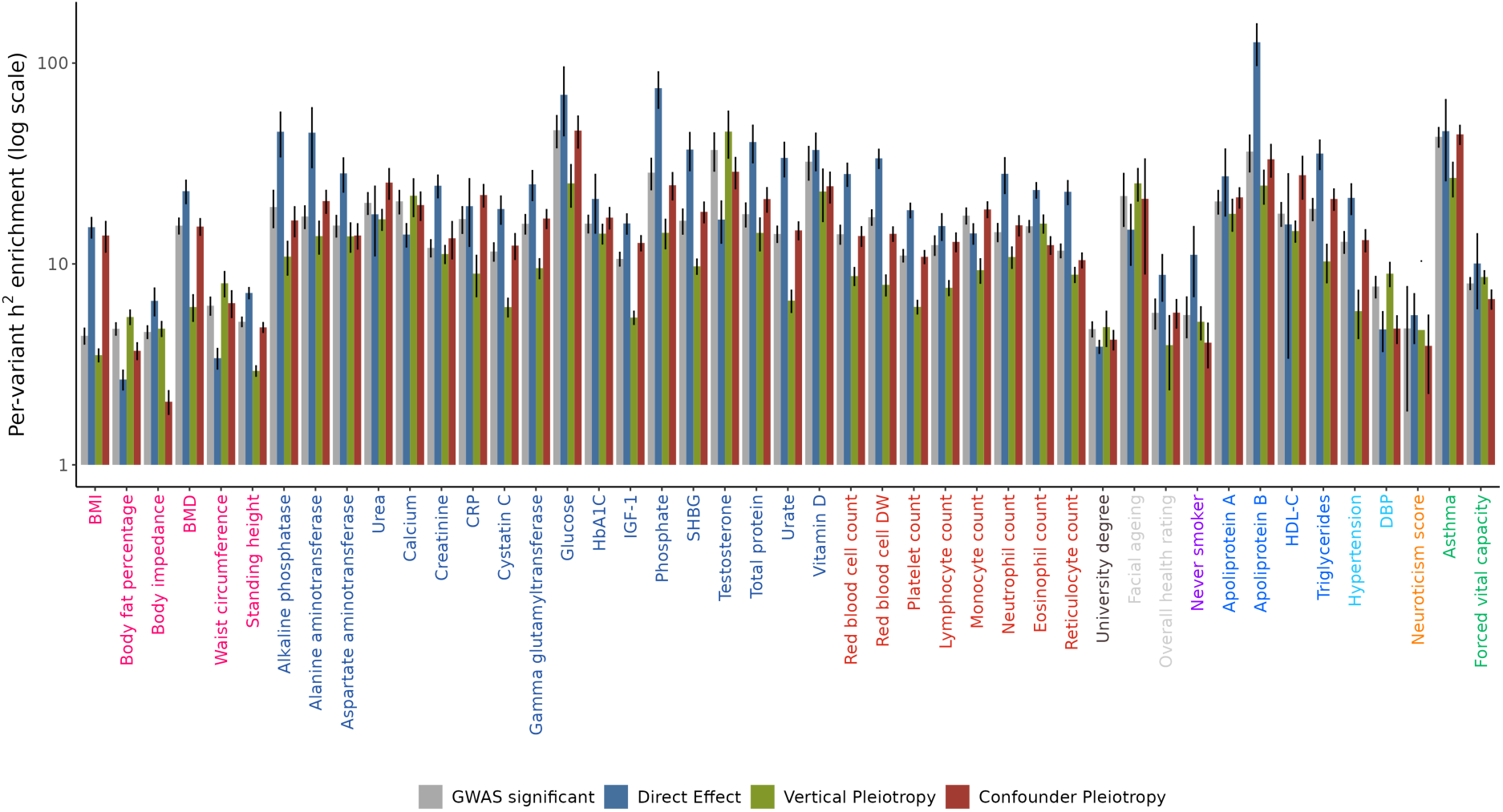
Enrichment in per-variant heritability of genetic variants, according to PRISM categories (direct, vertical, confounder) and GWAS. The x-axis represents the traits colored by broad categories. The y-axis represents the enrichment in per-variant heritability, estimated using stratified LD score regression and plotted on a logarithmic scale. Error bars represent the standard error of each enrichment estimate, also derived from stratified LD score regression.

### PRISM direct variant discoveries are robust to the set of included traits

Vertical and confounder pleiotropies occur when a genetic variant effect on a trait is mediated by another factor, either an identified trait or a latent confounder. Because it is neither conceptually nor computationally feasible to include every possible trait in the PRISM pipeline, we evaluated the robustness of PRISM results to missing traits, and thus potential missing confounders. Although a dependence of PRISM results on the set of included traits could be expected, we undertook several sensitivity analyses to show that results were mostly robust.

To begin, we focused on Overall health rating, a trait very likely to be influenced by causal relationships between traits and therefore particularly sensitive to omitting traits from the PRISM analysis. First, we systematically re-ran PRISM while sequentially removing each trait one by one, except Overall health rating. We compared the direct effects on Overall health rating obtained from these “incomplete” sets with those from the full set and observed that PRISM direct effect predictions remained highly consistent (Supplementary Fig. 11). Second, we ran s-LDSC on the incomplete sets and found that the enrichment of PRISM-labeled variants remained largely unchanged upon omitting traits (Supplementary Fig. 12).

Then, we extended this analysis to study all traits. We sequentially removed all traits that belonged to the same phenome category at once and reran PRISM. For every PRISM-significant variant, we re-assessed its labels under these incomplete sets. Even under these unfavorable conditions, PRISM direct effect predictions remained largely consistent (Supplementary Fig. 13). Moreover, analyzing the incomplete results with s-LDSC indicated once more that the enrichment of PRISM-labeled variants remained largely unchanged, despite ignoring substantial phenome categories of traits (Supplementary Fig. 14).

These findings demonstrate that PRISM results are robust and interpretable, even when not all potential traits are included.

### PRISM variants mapped to eGenes are found in relevant tissues

Standard follow-up analysis in GWAS involves checking whether the associated variants are eQTL in tissues linked to the studied trait. We expect to identify more interpretable and relevant eGenes using PRISM direct and pleiotropic categories. Therefore, we investigated the enrichment of relevant tissues in the expression of genes associated with eQTL variants corresponding to variants identified by PRISM or to GWAS variants. eQTLs and their corresponding eGenes were retrieved from GTEx^40^. Indeed, as an example for C-reactive protein (Supplementary Fig. 15), although the GWAS variants were not enriched in any tissue, we showed that direct variants exhibited a single very strong enrichment the liver, which is consistent with the fact that C-reactive protein is synthesized in the liver^41,42^. In contrast, although no enrichment was observed for either GWAS variants or direct variants for albumin, kidneys showed strong enrichment for vertical pleiotropy (Supplementary Fig. 16). This observation aligns with the role of the kidneys in filtering albumin^43^ and the established association between kidney disease and albumin levels^44,45^. These examples show that PRISM direct and pleiotropic categories are very likely to reflect specific and more directly interpretable underlying biological mechanisms.

### PRISM pinpoints direct variants that are mapped to more trait-specific genes than GWAS

Traditional pipelines for GWAS analysis typically involve examining genes mapped to significant variants, and their associated pathways. In our study, we notably performed annotation analysis to map functionally annotated variants to genes based on their physical positions in the genome and expression Quantitative Trait Locus (eQTL) mapping. Using FUMA^17^, we mapped genes to GWAS variants and to PRISM variants for each trait separately. Supplementary Fig. 17 compared the sets of common mapped genes between traits for GWAS and PRISM direct variants. Strikingly, GWAS mapped genes presented a very strong connectivity of traits compared to PRISM direct mapped genes. Subsequently, we labeled each gene that mapped to a significant PRISM variant with its corresponding PRISM label. In instances where multiple variants with different labels mapped to the same gene, we prioritized labeling in the following order: direct, vertical, and then confounder. Supplementary Fig. 18 illustrated that most genes shared between traits were involved in vertical or confounder pleiotropy. These observations suggest that genes mapped to direct variants may be more relevant, resulting in a more biologically interpretable variant-gene-trait network from PRISM. To further investigate the biological significance of our results, we conducted an enrichment analysis based on DisGeNET^46^ pathways using genes mapped to PRISM direct and GWAS variants. The goal was to identify enriched pathways and compare their significance between PRISM direct- and GWAS-mapped pathways. PRISM direct pathways generally exhibited greater enrichment compared to GWAS pathways in nearly two-thirds of the traits where clear differences were observed (Supplementary Fig. 19). Next, we employed bipartite network analysis to compare the centrality measures of gene-trait networks constructed from PRISM direct and GWAS variants. The gene network inferred from PRISM direct variants demonstrated lower degree and closeness, indicating fewer links between traits, but higher betweenness, suggesting the presence of more central traits (Supplementary Fig. 20). However, the GWAS-inferred network comprised significantly more gene nodes and edges due to the greater number of GWAS variants and their associated mapped genes compared to PRISM direct variants. To ensure a fair comparison, we conducted two distinct sensitivity analyses. First, we randomly removed gene nodes from the GWAS network to match the fewer node counts of PRISM network, without constraining the number of edges. This resulted in centrality measures similar to those of the full GWAS network (black dots). Second, we removed both genes and edges to create GWAS sub-networks with the same number of nodes and edges as PRISM direct network. Remarkably, with networks of the same exact dimension as PRISM, the centrality measures were equivalent for all the traits (red dots), and the network inferred from PRISM still demonstrated lower degree and closeness. This suggests that the network inferred from PRISM direct variants was more denoised and untangled, discarding redundant and biased relations induced by pleiotropy.

### PRISM produces coherent results on a panel of literature-validated variants

The final aim of PRISM is to infer genetic variant-trait networks. To confirm the reliability of PRISM, we aimed to compare its predicted networks to what the scientific literature suggests. We therefore examined whether the variant-trait effects identified and labeled by PRISM had already been previously identified. We reviewed variant-trait associations from a set of validated variants from Open Targets^47^. We chose to highlight two examples and the additional validated networks can be found in the Supplementary Results. First, variant rs7528419, residing in the SORT1 gene locus^48^ coding for the sortilin protein, was strongly associated with coronary heart disease (CHD) in GWAS^48^. According to PRISM (Fig. 5A), this variant displayed a vertical effect on CHD but a direct effect on apolipoprotein B (apoB). Essentially, PRISM suggested that this variant affects CHD only through its direct effect on apoB, which has a causal effect on CHD. Recent studies have indeed demonstrated that sortilin restricts the secretion of apoB^49^, and that apoB is one of the most accurate markers of lipid-induced cardiovascular risk^50^. Second, variant rs2282679, mapped to gene GC^51^, is validated for vitamin D levels. Kew et al.^52^ also highlighted that this gene is involved in white blood cells and neutrophil accumulation. Remarkably, these findings align with the 3 effects identified by PRISM coming from this variant (Fig. 5B). Additional examples are presented in the Supplementary Results.

**Figure 5:**
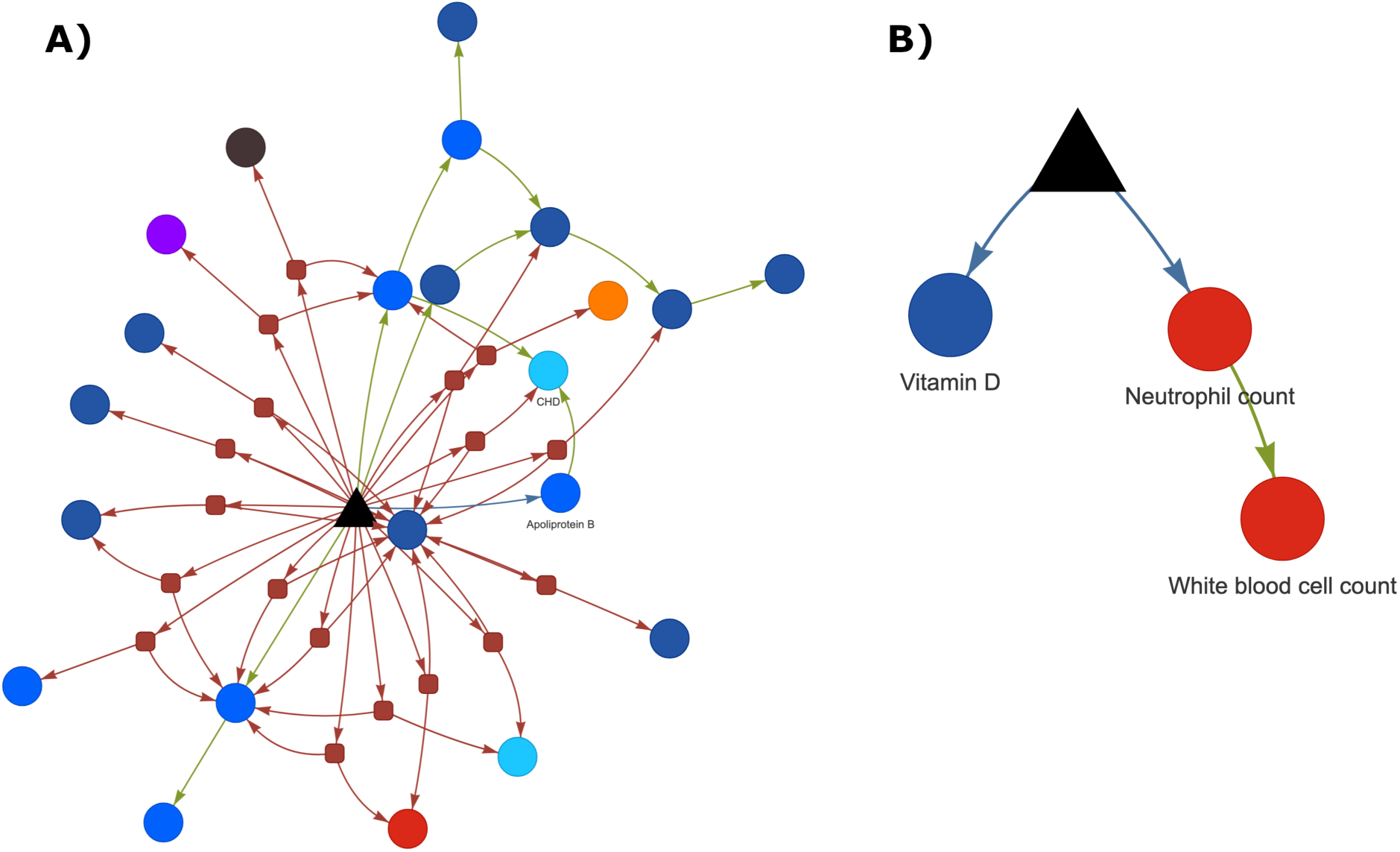
PRISM inferred causal network for variants rs7528419 (A) and rs2282679 (B). Genetic variants are represented as black triangles. Red arrows are effects of the variant through a confounder meaning confounder pleiotropy. Green arrows are effects of variant through a causal trait meaning vertical pleiotropy. Blue arrows are direct effects from the variant to traits. Confounders are represented as red squares and traits are represented as circles colored according to trait categories.

## Discussion

We developed PRISM (Pleiotropic Relationships to Infer the SNP model) that leverages pleiotropy to reassess the associations between genetic variants and traits from GWAS, by distinguishing between direct effects and pleiotropic effects. PRISM takes the opposite view to current methods, and does not resort to any annotation, but instead leverages pleiotropy which is pervasive throughout the human genome, to produce a network for each genetic variant.. We assessed the performances of PRISM on a simulated complex pleiotropic network of 60 traits and 426 confounders. We fine-tuned PRISM to achieve high precision in clustering direct variant-trait effects, which resulted in more limited power to detect significant variant-trait effects in highly polygenic traits. Consequently, variant networks predicted by PRISM could be incomplete, but importantly demonstrated a high level of accuracy. We applied PRISM to a set of 70 heritable traits and diseases from UK Biobank. PRISM was able to build a network for 400,143 unique variants. PRISM showed that most associations were caused by pleiotropy, as only ∼11% of variant-trait effects were direct. Direct variants predicted by PRISM were highly significantly enriched in per-variant heritability compared to GWAS significant variants and pleiotropic variants. Comparing biological pathways derived from PRISM direct variants and GWAS variants, the enrichment was stronger for PRISM. PRISM was able to pinpoint direct variants mapped to more trait-specific genes than GWAS, and the PRISM gene-trait network appeared disentangled and more pertinent compared to the GWAS gene-trait network. PRISM inferred relevant variant causal networks; we could show the concordance between the causal networks inferred by PRISM and networks from the literature for a panel of validated variants. Importantly, each causal network constructed by PRISM is unique and specific to its corresponding variant. All significant edges in these networks are individually computed by PRISM, which is different from integrating variants into pre-existing trait networks.

However, the proposed method has a number of limitations. First, its performance could depend on the set of traits under analysis, as pleiotropy is defined relative to the processed traits. Consequently, adding or removing traits, especially central ones, could influence whether variants reach significance and how their pleiotropy is clustered. We thoroughly investigated this issue both in simulations and real data. In real data, we demonstrated that removing large sets of traits had only a minimal effect on PRISM predictions, particularly regarding s-LDSC heritability enrichment. However, in simulations, omitting a central and pivotal trait could reduce PRISM accuracy in classifying variants involved with that trait (See Supplementary Fig. 21 and Supplementary Results) in the extreme case of a single unique mediator which is very unlike to happen in real data, as suggested in our sensivity analysis in real data. Given our procedure for trait inclusion designed to broadly represent the phenome, we infer that our dataset effectively includes proxies for most conceivable mediators. Therefore, the direct effects identified in our analysis predominantly represent true direct variant-trait effects, supporting the robustness and biological relevance of our findings. Second, PRISM relies on parameters computed by LHC-MR to assess the relationships between traits and to infer direct and pleiotropic labels. We extensively tested PRISM performances by adding noise to all the input parameters, and concluded that PRISM precision and recall are robust to reasonable parameter estimation errors (See Supplementary Results for details). Third, PRISM is geared towards precision to identify direct variant-trait effects. Hence vertical, and in particular confounder precisions are more limited. This is a deliberate choice since we aim to favor precision in the identification of direct effects. Fourth, the precision and recall were calculated from summary statistics simulated from a pleiotropic network of heritable traits. Similarly on real data, we only included heritable traits. Traits that are not heritable might disrupt the inferred causal network. Interpreting results that include very low heritability traits, which will be mechanically disadvantaged in comparison to highly heritable traits, must be performed with caution. However, we did simulate networks without any pleiotropy, and PRISM was still able to identify direct effects correctly (See Supplementary Results for details). Traits with very simple genetic architecture (*e.g.* monogenic diseases) can seemingly be processed without any issue. Fifth, as far as computational time is concerned, PRISM is limited by the obtention of pairwise traits input parameters from LHC-MR. The number of such pairwise computations increases exponentially as the number of traits increases. This is why we could not run PRISM on the full phenome-wide set of traits but instead used a procedure to select a representative set of 70 traits spanning broad categories that we hoped were well-representing the phenome.

On a different note, conventional strategies for disentangling GWAS associations could fall under fine-mapping, which aims at distinguishing between causal genetic variants and non-causal variants, using LD reference panels and genomic annotations^12,19–22^. Simply put, their objective is to attribute the real effect to a minimal subset of top variants in LD within a locus (at best, 1), and disqualify the other variants as LD effects. However PRISM takes an orthogonal approach to traditional fine-mapping methods, and aims not to distinguish between variants in linkage disequilibrium (LD), but to assess whether the observed effect of a variant on a trait, at the level of the locus, can be explained by pleiotropy, *i.e.* by another trait or a confounder. This has two main implications. First, we did not clump genetic variants when presenting biological results, to avoid eliminating the true causal variant effect in favor of an LD effect. Nevertheless, LD is still taken into consideration when calculating the probabilities of genetic variants to have causal effects. Variants with high LD scores are penalized because they are expected to mechanically exhibit larger effect sizes^53^. Second, we think that fine-mapping methods, such as SuSiE^54,55^, can complement PRISM by deciding between multiple candidate direct causal variants in LD. Combining these methods could enhance the identification of true causal variants within a locus and provide insights into the pleiotropy of the variants, which is crucial for future research on putative causal variants.

On a different note, direct effects and vertical pleiotropy are more straightforward concepts, while confounder pleiotropy presents a greater complexity. When a variant-trait effect is attributed to confounder pleiotropy, it indicates that the effect is mediated by an unidentified factor that PRISM could not precisely determine. This factor might be missing from the analysis, or the signal could be too entangled or underpowered to distinctly identify its origin. We hypothesize that, biologically, all instances of confounder pleiotropy could actually be vertical pleiotropy, where the mediating factor has yet to be clearly identified.

PRISM underlying model implies that direct effects result in stronger effect sizes in GWAS summary statistics. It would be a legitimate question to wonder whether direct variants are simply the most significant variants with the strongest effect sizes, while confounder and vertical variants are weaker significant variants. But although direct variants tend to have higher initial *t*-statistics from GWAS, PRISM results show much more intertwined results when looking at the original *t*-statistics of the different pleiotropies (Supplementary Fig. 22). For simple molecular traits like lipids or biomarkers, direct variants do tend to have higher *t*-statistics. However, complex traits like respiratory or metabolic traits do not follow this trend. In conclusion, we are confident that PRISM can effectively distinguish between subtle biological mechanisms.

We concluded that the gene network inferred from PRISM direct variants is more denoised and disentangled compared to the network inferred from GWAS variants. This observation raises the question of whether the reduced complexity of the PRISM-inferred network is due to PRISM limited power, potentially resulting in the detection of fewer significant variant effects and, consequently, a simpler network. However, our analysis suggests otherwise. We demonstrated that, even when reduced to the same number of nodes and edges as the PRISM network, the GWAS network still exhibited the tangled characteristics of the full GWAS network.

To apply the method, we prioritized statistical power by selecting a cohort with a large sample size, specifically the UK Biobank round 2, and could suffer from issues related to sample overlap. However, the bias from sample overlap is expected to be small when using a large sample size^28,56^. Another consequence of using UK Biobank, is that all the GWAS associations were obtained on individuals from “white British” ancestry^33^. Therefore, it would be valuable to apply PRISM to other ancestries and compare the inferred variant networks across different populations. Additionally, these results only cover common variants, as associations with rare variants are underpowered in GWAS^57^. Finally, these results are restricted to HapMap3 variants, but PRISM could be extended to more common variants.

Horizontal pleiotropy, defined as true multiple direct effects from a single genetic variant, is surprisingly rare, accounting for only 0.17% of the observed pleiotropy. In simulations, PRISM had decent power to detect horizontal variant-trait effects across a wide range of scenarios (See Supplementary Results for details). Therefore, we strongly believe that genetic variants with completely independent effects on multiple traits are extremely rare in the human genome, as we have previously shown^8^.

To conclude, at the trait level, PRISM can be used to better apprehend the genetic architecture of complex traits and the relationships between traits. At the variant level, PRISM can help understanding the specific genetic effect of a variant on multiple traits. We strongly believe that building the causal network of genetic variants is a priority to guide in vitro and in vivo follow-up studies as well as for medical applications, like genomic medicine or genome editing.

## Methods

### General principle of PRISM - Pleiotropic Relationships to Infer the SNP Model

The general objective of PRISM is to infer a network for each genetic variant from GWAS summary statistics, distinguishing between direct and pleiotropic effects. Essentially, each variant-trait association is reassessed conditional on each one of the other traits, *i.e.* in different contexts. PRISM determines whether the variant-trait association remains consistent in the different contexts, ultimately clustering the effect as direct, pleiotropic, or null. Specifically, a “confounder pleiotropy” label indicates that the variant effect operates through a shared confounder with another trait, while a “vertical pleiotropy” label signifies that the effect is mediated by a causally related trait. In contrast, a “direct effect” represents a significant, non-pleiotropic influence of the variant on the trait. After analyzing all traits, PRISM uses the resulting direct and pleiotropic labels to construct a network that links each variant to the traits it impacts.

#### PRISM models variant-trait associations conditionally to multiple other traits

Let *X* denote a trait and *β̂_k_^x^* denote the standardized effect of a genetic variant *k* on trait *X*, observed from GWAS summary statistics. PRISM aims to re-evaluate the effect of variant *k* on *X* by leveraging information from additional traits. Specifically, it seeks to determine whether the observed association between variant *k* and trait *X* arises from a direct genetic effect of *k* on *X*, or is mediated by confounders or other traits. To address this, we begin by modeling the effect of variant *k* on *X* while sequentially accounting for each one of the other traits. Let *Y* denote another trait and *β̂_k_^y^* denote the standardized effect of the genetic variant *k* on trait *Y*. Let *U* denote a latent confounder with causal effects on both *X* and *Y* (Supplementary Fig. 1). *X* and *Y* may have causal effects on each other. We model the joint distribution of *β̂_k_^X^* and *β̂_k_^Y^* as an 8-component bivariate Gaussian Mixture Model. Each component corresponds to the expected distribution of standardized effects from variants in that particular component. The eight components are: (0) No association; (1) Associated with *X*; (2) Associated with *U*; (3) Associated with *Y*; (4) Associated with *X* and *U*; (5) Associated with *X* and *Y*; (6) Associated with *U* and *Y*; (7) Associated with *X*, *Y* and *U*.

Every variant effect is hypothesized to be drawn from one of these components representing a cluster. Then we use a classical clustering strategy to assign variants to clusters based on their likelihood of belonging to each Gaussian component. Specifically, the probability that a given variant *k* belongs to Gaussian component *j* is determined by the following posterior probability:

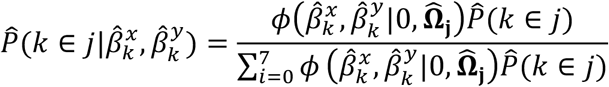

Here, **Ω̂^j^** denotes the variance-covariance matrix of Gaussian component *j*. The function *ϕ* represents the joint probability density function of the bivariate normal distribution, and *P̂*(*k* ∈ *j*) is the prior probability of variant *k* belonging to component *j*.

For each variant *k*, we compute the probability of belonging to each component. Then, we translate these probabilities into two scores corresponding to the variant having no effect, or an effect on *X*. <u>No Effect</u>: the score indicating no effect of variant *k* on neither *X* nor *Y* corresponds to the probability of belonging to component (0). <u>Effect on *X*</u>: the score indicating an effect of variant *k* on *X* corresponds to the highest probability among the components that include an effect on *X*, specifically components (1), (4),

1. and (7).

#### PRISM tests and classifies variant-trait associations

Let us consider now a set of *T* traits used to evaluate the associations of variant *k* on *X*. First, PRISM computes the previous step and models the bivariate distribution of the variant effect on *X* with each one of the *T* traits. This leads to obtain *T* paired scores (*i.e.* “no effect” and “effect on *X*“) capturing the effect of the variant *k* on *X* conditional on each one of the *T* traits representing as many pleiotropic contexts. Second, the paired scores are compared using a sign test which determines whether the score on *X* tends to be systematically greater than the null score, no matter what the context, *i.e.* for every one of the conditional *T* traits. Third, we apply a Bonferroni correction to identify a significant variant-trait effect, in addition to the usual GWAS threshold: 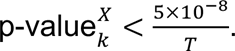

After ensuring the robustness of effect of variant *k* on trait *X* (*i.e.* significant effect according to the sign test), the variant-trait effect is then labeled with confounder, vertical or direct effects, conditional on each one of the *T* pleiotropic contexts. This classification relies on the previously estimated probabilities for the Gaussian mixture model components. Specifically:

1. Confounder pleiotropy: The effect is labeled as confounder if, in any of the pleiotropic contexts, the effect appears to be mediated exclusively by a confounder.
2. Vertical pleiotropy: The effect is labeled as vertical if, in any of the pleiotropic contexts, the effect appears to be mediated by another trait that is causal to *X*.
3. Direct effect: If the effect does not appear to be mediated by vertical or confounder pleiotropy, the effect is labeled as a direct effect.

Thus, for each variant *k*, PRISM assigns a label categorizing its effect on *X* upon significance (See Supplementary Methods for details). Since this procedure is extended for all included traits, PRISM assigns a label to the effect of each variant on each trait. Supplementary Fig. 23 provides an overview of PRISM full analysis pipeline.

Finally, for each variant, we gather all the labels inferred from PRISM into a network where nodes represent the variant and traits, and edges represent the direct, vertical, or confounder effects. PRISM provides vertical labels involving a given variant and a pair of traits, therefore this initial network is then pruned by removing vertical edges between the variant and a given trait, conditioned on other traits involved in the vertical relationships. Concretely, if the effect of the variant on a given trait can be explained by a mediated effect from another trait, we remove the edge from variant to trait. In addition, when a variant shows vertical effects on multiple traits through a common causal trait, we eliminate redundant edges.

#### Parameter estimation: PRISM uses global trait pleiotropic relationships parameters

To apply PRISM, we need to estimate **Ω**_**j**_ and *P*(*k* ∈ *j*) for each pair of traits. As described in the Supplementary Methods, **Ω**_**j**_ and *P*(*k* ∈ *j*) can be expressed using the number of genetic variants *m*, the sample size of each trait, and a set of global parameters *θ* that quantifies the bidirectional causal effects between a pair of traits *X* and *Y*, the effects of the confounder, both trait heritabilities, both trait polygenicities, as well as their respective LD score intercepts. The estimation of *θ* is performed using LHC-MR^28^, an integrative Mendelian Randomization method that accounts for a latent heritable confounder. LHC-MR takes as inputs the GWAS summary statistics for a pair of traits and outputs a set of *θ* parameters. Since LHC-MR and PRISM share a similar underlying mathematical model, the *θ* parameters estimated by LHC-MR can be used as input for PRISM.

### Simulation framework

#### Simulating GWAS summary statistics data for a complex pleiotropic network of traits

We constructed a pleiotropic network consisting of 60 simulated traits organized into four subnetworks with similar structure (See Fig. 6). Each subnetwork consisted of five groups of traits *A*, *B*, *C*, *D* and *E*. *A* connected directly to *B*_3_ and *B*_4_, while *B*_1_ and *B*_2_had mutual causal influences and *D* was causal to *A*. *B*_4_ also connected to traits from the *C* group, which made *B*_4_ the most pivotal trait in our subnetwork. Traits from group *E* had no causal relationships with any other traits. In addition a confounder was simulated for each pair of traits in the subnetwork. To link the subnetworks, a confounder was simulated between each pair of traits *B*_4_ across subnetworks. In total, the network was composed of 60 traits, and 426 confounders (105 confounders for each subnetwork with 6 cross-subnetwork confounders for traits *B*_4_). *m* = 100,000 genetic variants were simulated for each of the 60 traits generating GWAS summary statistics. We created 48 different scenarios, with various parameters for heritability (0.2, 0.6), polygenicity (0.01, 1 and 10 %), causal relationships (0.05, 0.3), and strength of the confounder, as detailed in Supplementary Table 1. To approximate a genome-wide model while maintaining computational feasibility, we opted for sets of 100,000 simulated variants. The standardized effects of all these variants on all the traits were simulated, taking into account the relationships between the traits. First, for each trait *X*, we randomly selected genetic variants to have a true direct effect on trait *X* and on the confounders of the relationships between trait *X* and the other traits. For the selected genetic variants, the true direct effects were drawn from a Gaussian distribution with parameters depending on the trait and the scenario. Then, the true effects were propagated to the other traits through vertical and confounder pleiotropy. Additionally, the effects were propagated according to the LD structure of each variant, using the real LD matrix of chromosome 1 derived from 1000 Genomes^58^. Finally, an error term was added. This produced standardized effects on all traits and LD scores for all genetic variants (See Supplementary Methods for details). The LD score of a given variant was computed as the sum of its LD with all variants (including itself), using the same LD matrix.

**Figure 6:**
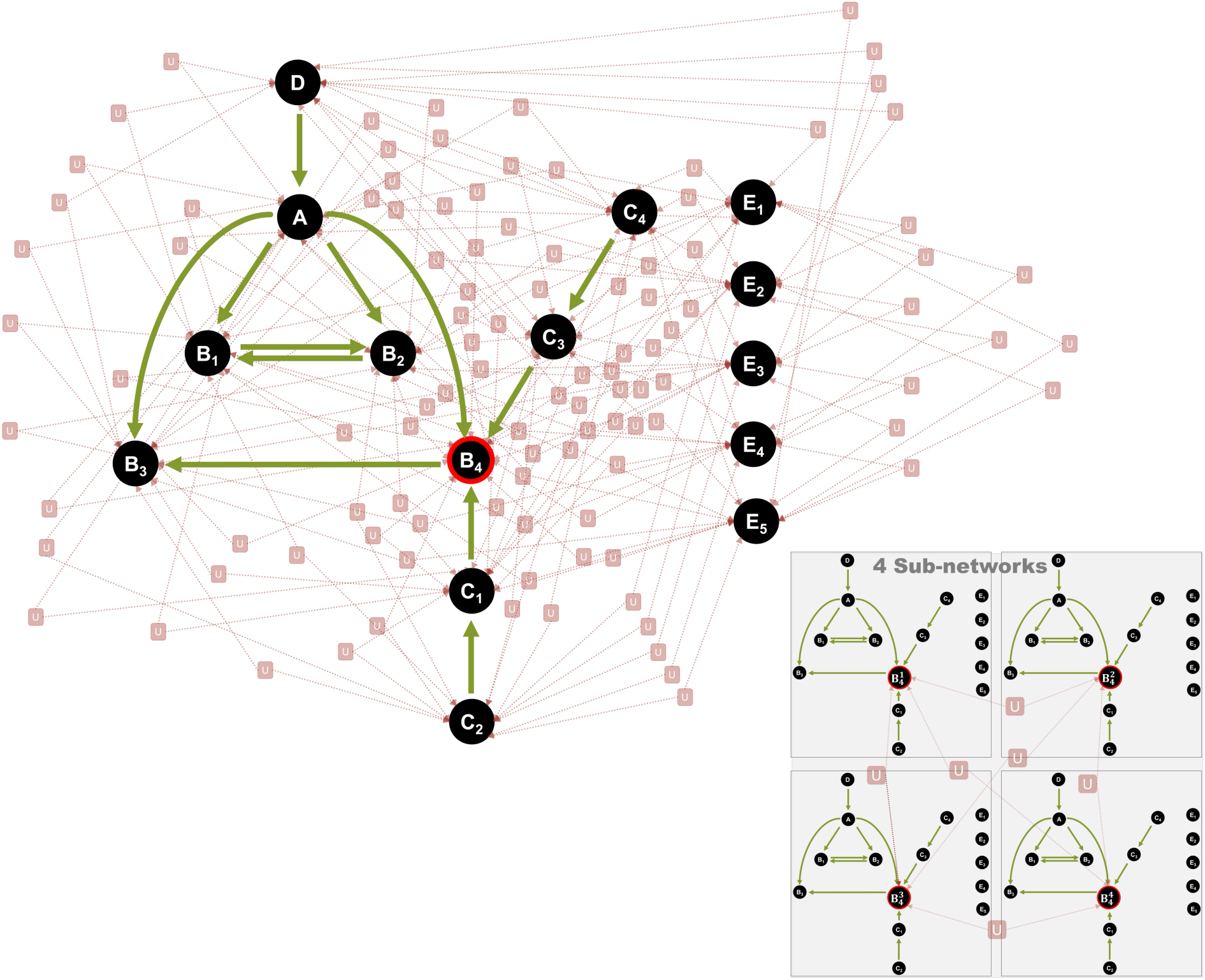
Architecture of the simulated trait network. Simulated traits are represented as circles, and green arrows indicate vertical pleiotropy between traits. Within each subnetwork, every pair of traits shares a unique confounder *U*. The global network is composed of four subnetworks, shown in the bottom-right panel. Additionally, a confounder is simulated for each pair of *B*_1_ traits across the subnetworks.

#### Calculating PRISM precision and recall

For each scenario, we processed the simulated data through PRISM to identify and label significant variant-trait effects. The objective was to compare the true labels used for simulated data generation with the labels inferred by PRISM. In our complex simulations, defining the ground truth is not straightforward, since multiple effects can propagate across traits and through LD correlations. We therefore considered two complementary approaches to compute precision and recall.

First, we used an *all effects* approach, in which a given LD block could have multiple predicted and true labels. In this approach, all predicted effects within the block were retained, and a prediction was considered correct if it matched any of the corresponding true effects. This approach is more appropriate for assessing whether PRISM predicts effects that are genuinely present in the simulated ground truth and is only used to assess the presence of false positives from a statistical point of view.

Second, we used a *prioritized effects* approach, in which a single label was assigned to each LD block. The predicted label of a given LD block was defined as the predicted label of the most significant variant within that block. The true label of each LD block was then assigned according to the following hierarchy: 1) “direct effect” if at least one variant within the block had a true direct effect; 2) “vertical effect” if at least one variant had a true vertical effect and no variant had a true direct effect; 3) “confounder effect” if at least one variant had a true confounder effect and no variant had a true direct or vertical effect; and 4) “no effect” otherwise. This approach is more informative for interpretation, but also more stringent, since prediction by PRISM could be considered incorrect when it did not correspond to the prioritized effect in the block, even if the predicted effect was present in the simulated ground truth.

In both approaches, we compared predicted labels with true labels by calculating precision and recall for each of the three effect labels. For a given label, these metrics were computed as follows: 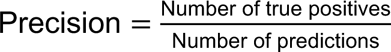 and 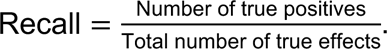 Unless otherwise stated, precision and recall were calculated globally across corresponding traits in the four subnetworks. For example, the precision and recall reported for trait *B*_4_ were obtained by aggregating variant-trait effects on *B*_4_^1^, *B*_4_^2^, *B*_4_^3^, and *B*_4_^4^. This aggregation was performed only for visualization purposes: trait specificity was preserved in the calculation, such that a predicted effect on *B*_4_^1^ was not considered correct if the corresponding true effect was on *B*_4_^2^.

#### Comparing PRISM predicted variant networks and true variant networks

A PRISM variant network consists of nodes, representing the variant and the traits, and edges, representing the variant-trait and trait-trait effects labeled with direct and pleiotropic labels. To compare predicted variant networks with true variant networks, we regrouped edges from variants within the same LD block. Then, we applied a weighted Simple Matching Coefficient (SMC), defined as follows: 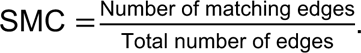 We implemented stringent criteria, requiring edges to be identical in terms of nodes, direction, and type of pleiotropy to be considered a match. Confounder pleiotropy inherently produces more edges than vertical pleiotropy, which in turn produces more edges than direct effects. Consequently, direct edges were assigned a weight of 1, vertical edges a weight of 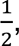 and confounder edges a weight of 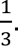

### Application to 70 traits from UK Biobank

To fully leverage PRISM potential, we selected an extensive array of complex traits and diseases from the UK Biobank^33^. However, due to computational constraints, we were unable to include all 4,178 available traits. Instead, we used the following procedure to ensure proper generalization of our results. First, we included biomarkers with a p-value for heritability lower than 0.05/31, among all 31 available biomarkers. Biomarkers were then categorized into *general biomarkers*, *blood traits*, and *lipid traits*. For the traits and diseases, we did a first manual selection of the traits to span a comprehensive ensemble of categories corresponding to anthropometric, cancer, education, global health, inflammatory, lifestyle, metabolic, psychiatric, and respiratory traits. In order to be as comprehensive as possible, we sorted through the remaining traits by excluding nonheritable traits, *i.e.* traits with a p-value associated with heritability higher than 0.05/4,147, and by excluding traits with a genetic correlation with any of the included trait higher than 0.2 in absolute value according to cross-trait LDSC regression. In addition, we excluded two traits linked to habits (*i.e.* 1150_2 - Usual side of head for mobile phone use: Right, 1150_1 - Usual side of head for mobile phone use: Left) with minor interest. Finally, we excluded the covariates, sex-specific traits, traits with a sample size below 100,000 and traits with reported issues (*e.g.* low number of cases). The final set encompassed 70 traits spanning 12 broad categories (Supplementary Table 2).

Variants from the human leukocyte antigen (HLA) region on chromosome 6 were excluded due to the high density of variants associated with autoimmune and infectious diseases, and the complex LD structure in this region^28^. Variants from sex chromosomes were excluded due to the complexity of their unique inheritance patterns and gene expression differences between sexes. Low-confidence variants and rare variants with a minor allele frequency (MAF) below 0.05 were also excluded due to their insufficient statistical power. Only variants present in the HapMap3 framework were analyzed, because LHC-MR is limited to these variants. We then processed these 70 traits through PRISM.

#### Computing per-variant heritability enrichments

To interpret PRISM results, we employed stratified LD score regression (s-LDSC)^39^. We calculated per-variant heritability enrichments across labeled variant-trait effects, separately for each trait. Specifically, for each trait, we selected GWAS-significant variants by applying a p-value threshold of p-value 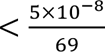 corresponding to the traditional GWAS threshold divided by the total number of traits -1. The selected variants were categorized into four groups: a GWAS-significant group and three non-overlapping PRISM-labeled groups (direct, vertical, confounder). The GWAS-significant group comprised all variants, while the PRISM-labeled groups partitioned the GWAS-significant variants based on PRISM predicted labels. Among the 70 traits, 45 had enough variants in all four groups to be processed by s-LDSC.

#### eQTL and eGenes retrieval from GTEx

We retrieved the eQTL and their corresponding eGenes from GTEx^40^ version 8. We converted the genomic coordinates from Hg38 to Hg19 using the liftOver R package. We identified significant variant-eGene pairs under the following threshold p-value < 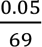 in each tissue. Variants were grouped into four categories: three categories (direct, vertical, confounder) for PRISM significant variant effects, and a category for GWAS significant associations. For each tissue, we then determined the number of unique eGenes significantly paired with variants separately in each one of the four categories. Finally, for each tissue, we calculated the enrichment in unique genes of each category as: 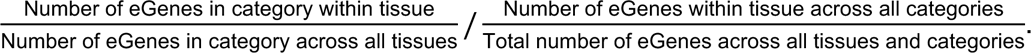 The enrichment was set to 0 if the number of eGenes in category within tissue < 5. Results for all 70 traits are available here.

#### Gene networks and pathways analysis

We configured FUMA with specific parameters to ensure accurate and reliable results. We used Ensembl version 110 as the reference, and the UK Biobank release 2b population dataset “WBrits10k” was chosen as the reference panel. A physical mapping window of 10 kb was employed to precisely map variants to nearby genes. We set the significance threshold p-value 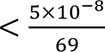 for GWAS and PRISM variants. Additionally, we incorporated eQTL mapping into our analysis using GTEx data version 6, including all available tissue types. These analyses were performed for the 70 studied traits, and we obtained mapped genes for 67 traits using FUMA for both methods (PRISM and GWAS variants). Next, using FUMA results, we conducted pathway enrichment analyses with Metascape^15^ on DisGeNET^46^ pathways for genes mapped to GWAS and PRISM direct variants. 60 traits had common genes between the two methods, and among them, 53 traits shared common pathways. Finally, on the same set of 60 traits, we constructed bipartite networks, which consist of two distinct sets of nodes (here, genes and traits) where edges only exist between nodes of different sets. Edges were added between traits and their associated genes. We performed sensitivity analyses on 10 replicates, which adjusted the GWAS network to match the number of genes and edges in PRISM, thereby allowing for an unbiased comparison of network properties. The *degree centrality* of a trait, indicating the number of associated genes, was calculated as: 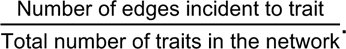 The *betweenness centrality*, measuring the extent to which a trait lies on the shortest paths between other traits, was calculated as: 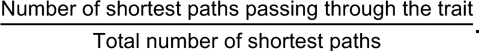 The *closeness centrality*, assessing the proximity of a trait to all other traits in the network, was calculated as 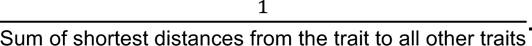

## Data availability

PRISM results are available on [Zenodo](https://doi.org/10.5281/zenodo.15074674) as well as accessible through an online user-friendly interface. Indeed, we developed a [R Shiny interface](https://verbam01.shinyapps.io/PRISM/), freely available online, to display PRISM results on our network of 70 highly heritable traits. Results can be visualized at the trait level and also to display the causal network of any genetic variant of interest.

## Code availability

PRISM is implemented in R and a user-friendly tutorial is available on [GitHub](https://github.com/martintnr/PRISM). As long as GWAS summary statistics are available and the studied variants are mapped in HapMap3, PRISM can compute any network of traits of interest to distinguish direct variant-trait effects from vertical and confounder variant-trait effects. PRISM should not be applied to fewer than 32 traits, as this is the minimum number required for the sign test to produce a p-value below the threshold 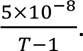

## Funding

MT, AN, and MV are supported by the French National Research Agency (ANR) (ANR-21-CE45-0023-01). MT and MFM are supported by the data intelligence institute of Paris (diiP, IdEx Université Paris Cité, ANR-18-IDEX-0001). MFM is a PhD student from the FIRE PhD program funded by the Bettencourt Schueller foundation and the EURIP graduate program (ANR-17-EURE-0012).

## Authors contributions

MT contributed to study conception, data analysis, interpretation of the results, drafting and critical revisions of the manuscript; AN contributed to data analysis, interpretation of the results and revision of the manuscript; MFM contributed to revision of the manuscript; YR contributed to study conception and revision of the manuscript; MV supervised the study and contributed to study conception, data analysis, interpretation of the results and critical revision of the manuscript.

## Acknowledgements

We wish to sincerely thank Dr Emmanuel Curis and Dr Virginie Lasserre for their helpful discussions and guidance on our work.

## Notes

### Competing Interest Statement

The authors have declared no competing interest.

### Author Declarations

The study used only openly available human data in the form of summary statistics from the UK Biobank.

### Summary of Updates

Additional simulations have been added in Main and Supplementary. Few minor text clarifications.

