## Supplementary Material for "Inferring genetic variant networks by leveraging pleiotropy shows trait relationships drive massive pleiotropy in GWAS"

#### Supplementary Figures

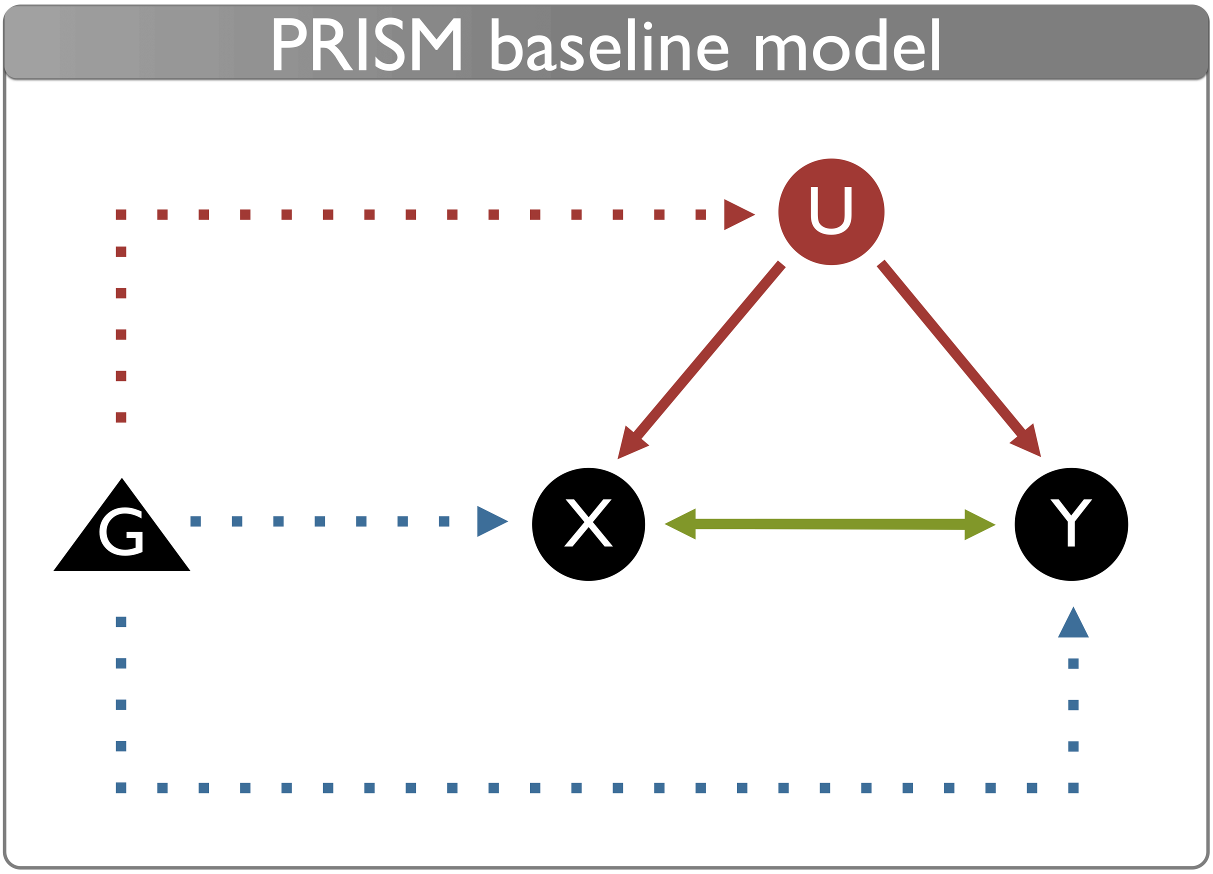


***Supplementary Figure 1.*** *$X$ and $Y$ are two complex traits, and $U$ a latent confounder with causal effects on each other. $G$ represents a genetic variant, with putative effects on $X$, $Y$ and $U$.*

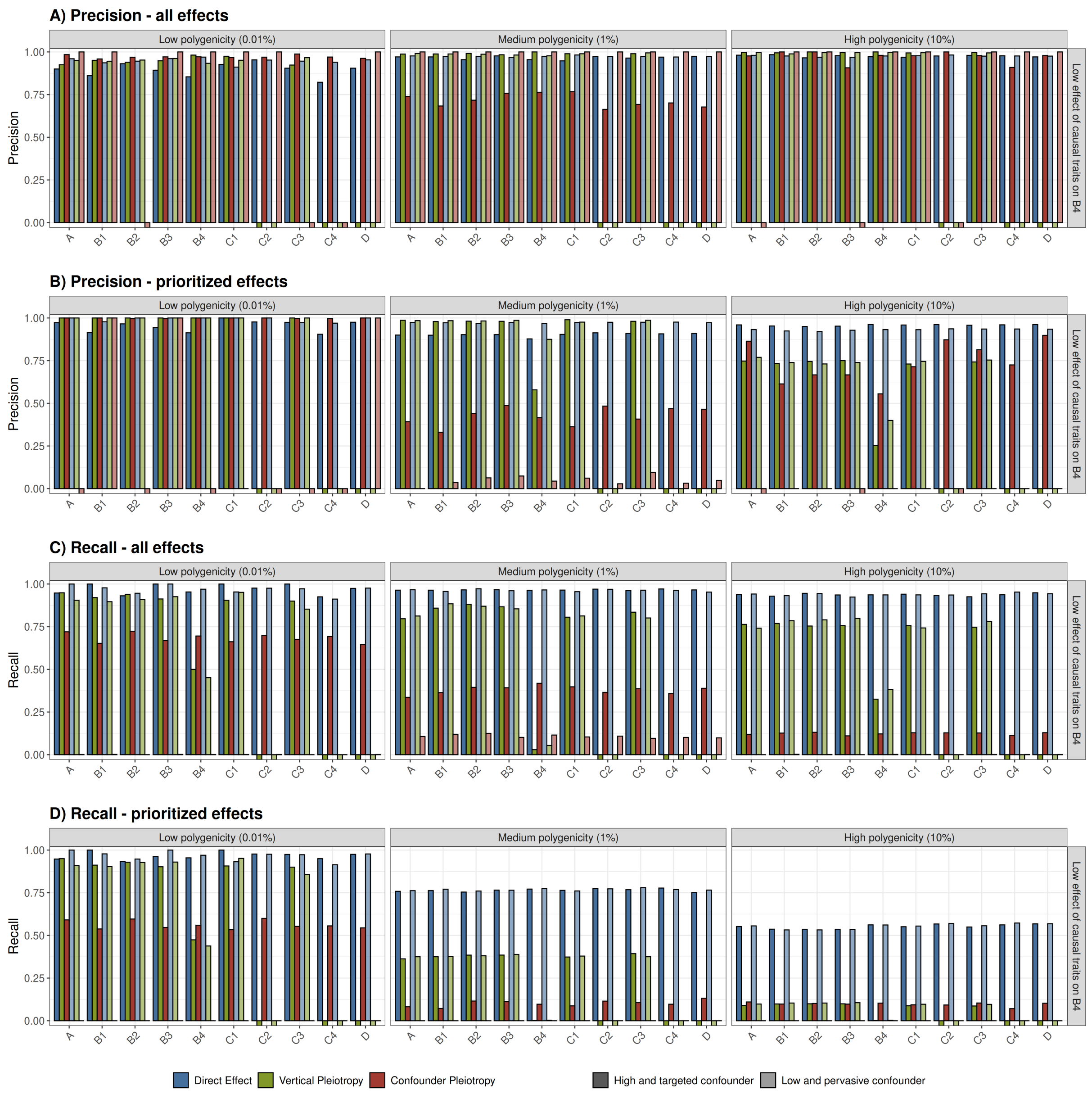


***Supplementary Figure 2. A) B)*** *Precision and* ***C) D)*** *recall of PRISM predictions for significant variant-trait effects, on simulations, with low causal effect on $B_4$.* ***A)*** *and* ***C)*** *represent the performance of PRISM when multiple predicted and true labels are allowed (i.e. all effects, see Methods).* ***B)*** *and* ***D)*** *represent the performance of PRISM when each true variant-trait effect is assigned a single prioritized label for interpretation (i.e. prioritized effects, see Methods). The x-axis represents the simulated traits, grouped across subnetworks for visualization purpose (See Fig. 6 and Methods). Significant effects are defined with $P < 5 \times 10^{-8}/59$, the recommended threshold from PRISM (See Methods). Bars are colored according to predicted labels: direct effect (blue), vertical pleiotropy (green), or confounder pleiotropy (red). Six scenarios are represented across facets, with varying parameters of polygenicity and type of confounder. Polygenicity represents the proportion of variants with a direct effect on each trait. Causal effect on $B_4$, the most pivotal trait in each subnetwork, represents the proportion of effect passed to each one of the four $B_4$ traits, for all traits with a non-zero vertical effect on $B_4$. High targeted confounder (darker shades) means that few variants (0.01%) have an effect on the confounder $U$, but with magnitude of effect rivaling direct effects. Low pervasive confounder (lighter shades) means that a large proportion (5%) of variants have an effect on the confounder, but with low magnitude. All traits are simulated with high heritability (60%). For precision, the y-axis represents the proportion of well-predicted variants among all predicted variants for a given label. For recall, the y-axis represents the proportion of well-predicted variants among all true variants for a given label. NB: In cases where no variant was clustered under a given label, it was impossible to calculate precision or recall; such instances are represented by squares below the x-axis. Results for high effect on $B_4$ can be found in Fig. 2. Results for traits $E1$-$E5$ can be found in Supplementary Fig. 3.*

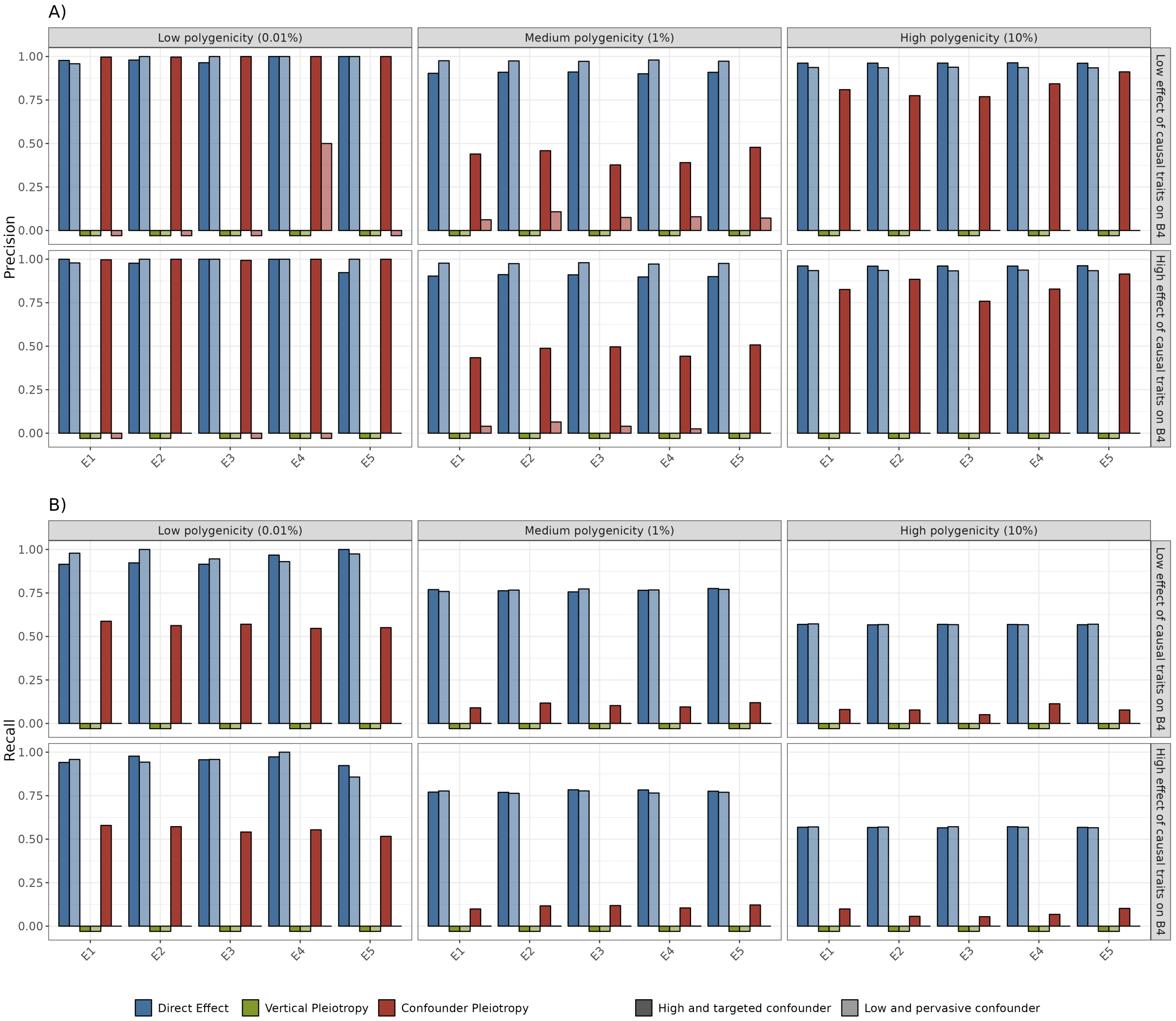


***Supplementary Figure 3. A)*** *Precision and* ***B)*** *Recall of PRISM predictions for significant variant-trait effects, on simulations, for all $E$ traits. The prioritized effects approach is used to compute precision and recall. The x-axis represents the simulated traits, grouped across subnetworks for visualization purpose (See Fig. 6 and Methods). Significant effects are defined with $P < 5 \times 10^{-8}/59$ , the recommended threshold from PRISM (See Methods). Bars are colored according to predicted labels: direct (blue), vertical pleiotropy (green) or confounder pleiotropy (red). Twelve scenarios are represented across facets, with varying parameters of polygenicity, strength of causal effect and type of confounder. Polygenicity represents the proportion of variants with a direct effect on each trait. Causal effect on $B_4$, the most pivotal trait in each subnetwork, represents the proportion of effect passed to each one of the four $B_4$ traits, for all traits with a non-zero vertical effect on $B_4$. High targeted confounder (darker shades) means that few variants (0.01%) have an effect on the confounder U, but with magnitude of effect rivaling direct effects. Low pervasive confounder (lighter shades) means that a large proportion (5%) of variants have an effect on the confounder, but with low magnitude. All traits are simulated with high heritability (60%).* ***A)*** *The y-axis represents the precision which is the proportion of well-predicted variants among all predicted variants for a given label.* ***B)*** *The y-axis represents the recall, which is the proportion of well-predicted variants among all true variants for a given label. NB: In cases where no variant was clustered under a given label, it was impossible to calculate precision or recall; such instances are represented by squares below the x-axis. Results for traits $A$-$D$ can be found in Fig. 2 and Supplementary Fig. 2.*

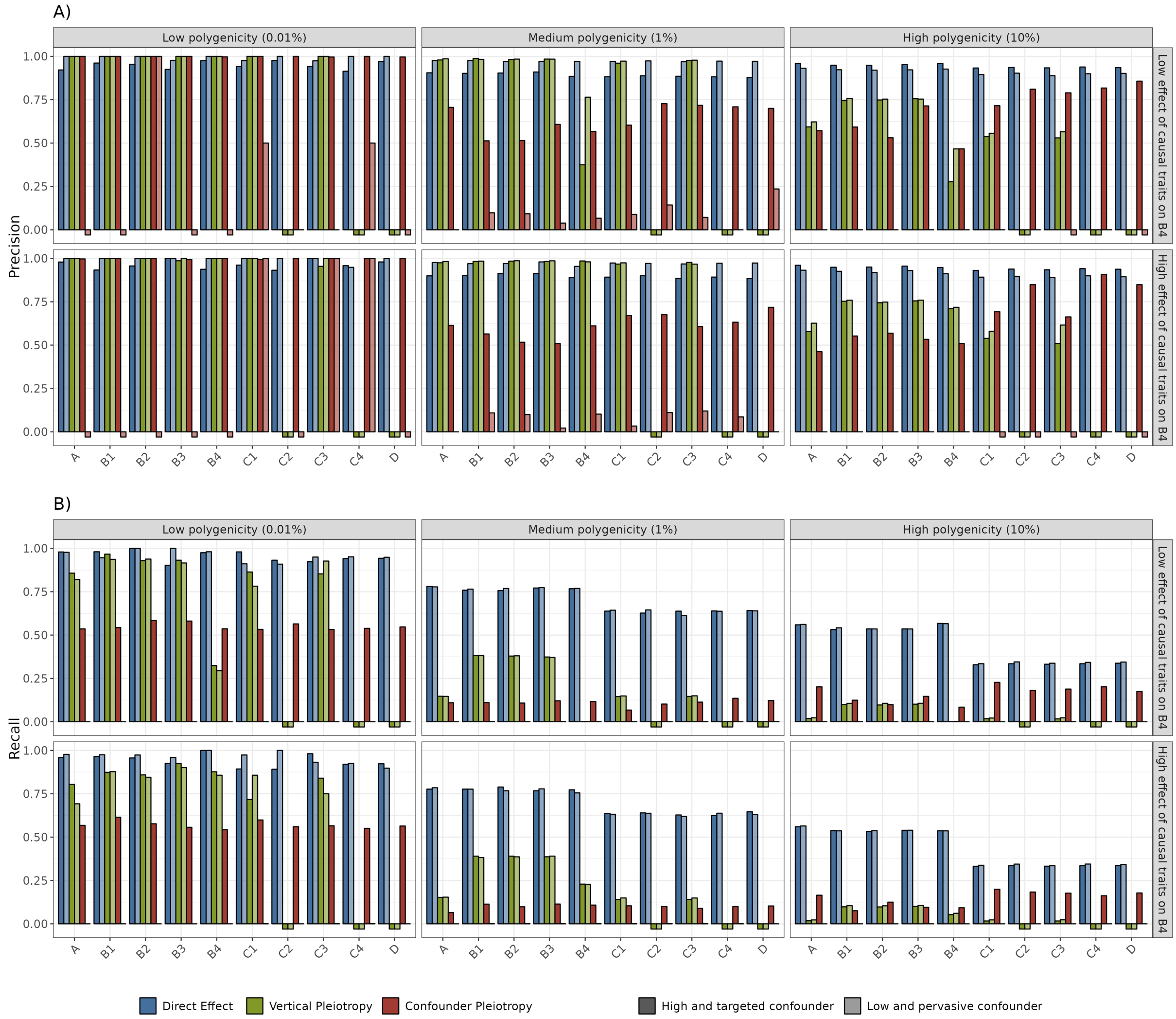


***Supplementary Figure 4. A)*** *Precision and* ***B)*** *Recall of PRISM predictions for significant variant-trait effects, on simulations, with varying trait heritability. The prioritized effects approach is used to compute precision and recall. The x-axis represents the simulated traits, grouped across subnetworks for visualization purpose (See Fig. 6 and Methods). Significant effects are defined with* $P < 5 \times 10^{-8}/59$ *, the recommended threshold from PRISM (See Methods). Bars are colored according to predicted labels: direct (blue), vertical pleiotropy (green) or confounder pleiotropy (red). Twelve scenarios are represented across facets, with varying parameters of polygenicity, strength of causal effect and type of confounder. Polygenicity represents the proportion of variants with a direct effect on each trait. Causal effect on* $B_4$*, the most pivotal trait in each subnetwork, represents the proportion of effect passed to each one of the four* $B_4$ *traits, for all traits with a non-zero vertical effect on* $B_4$*. High targeted confounder (darker shades) means that few variants (0.01%) have an effect on the confounder U, but with magnitude of effect rivaling direct effects. Low pervasive confounder (lighter shades) means that a large proportion (5%) of variants have an effect on the confounder, but with low magnitude. Traits* $A$ *and* $B$ *are simulated with high heritability (60%), while traits* $C$*,* $D$ *and* $E$ *are simulated with lower heritability (20%).* ***A)*** *The y-axis represents the precision which is the proportion of well-predicted variants among all predicted variants for a given label.* ***B)*** *The y-axis represents the recall, which is the proportion of well-predicted variants among all true variants for a given label. NB: In cases where no variant was clustered under a given label, it was impossible to calculate precision or recall; such instances are represented by squares below the x-axis.*

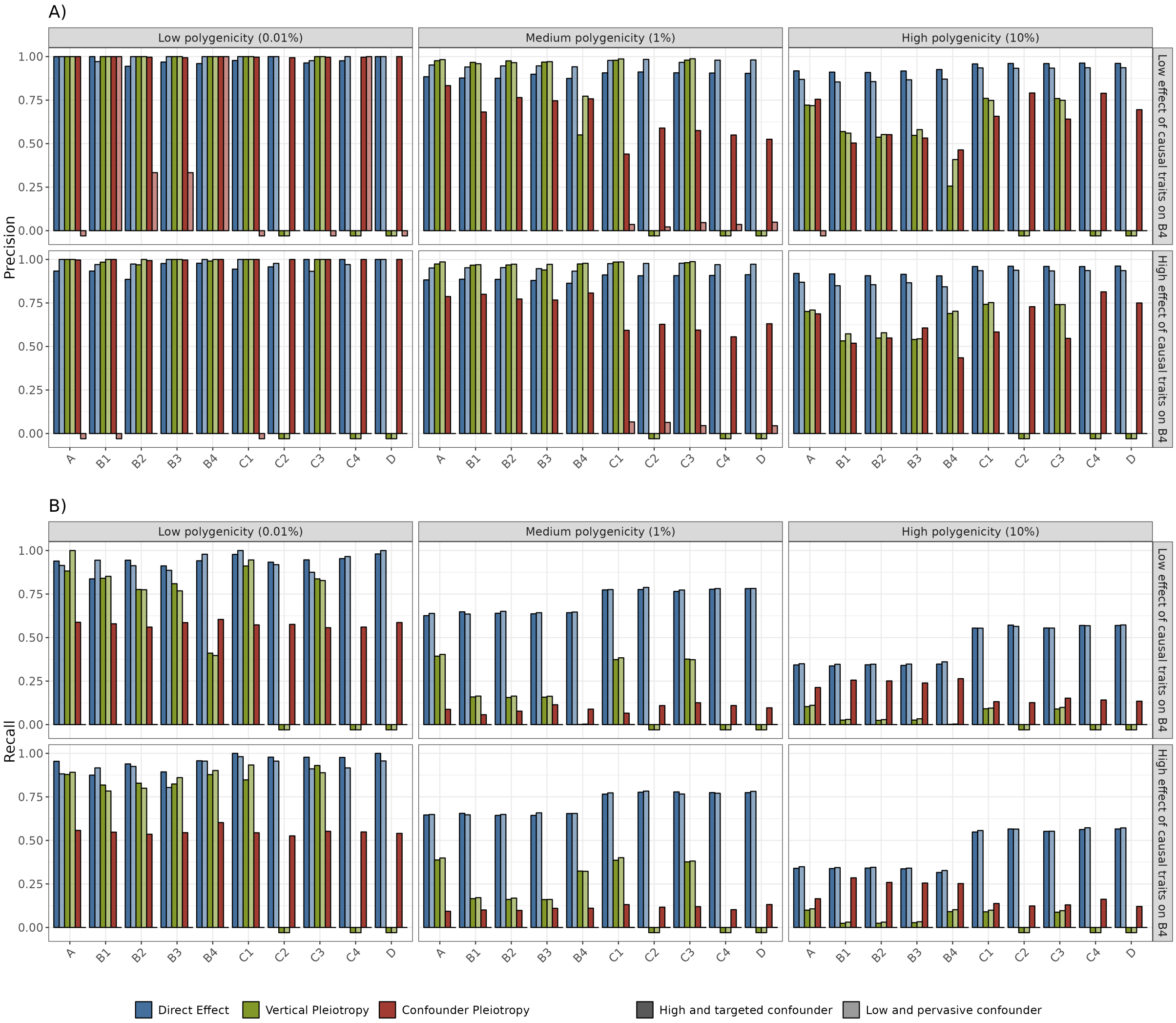


***Supplementary Figure 5. A)*** *Precision and **B)** Recall of PRISM predictions for significant variant-trait effects, on simulations, with varying trait heritability. The prioritized effects approach is used to compute precision and recall. The x-axis represents the simulated traits, grouped across subnetworks for visualization purpose (See Fig. 6 and Methods). Significant effects are defined with $P < 5 \times 10^{-8}/59$ , the recommended threshold from PRISM (See Methods). Bars are colored according to predicted labels: direct (blue), vertical pleiotropy (green) or confounder pleiotropy (red). Twelve scenarios are represented across facets, with varying parameters of polygenicity, strength of causal effect and type of confounder. Polygenicity represents the proportion of variants with a direct effect on each trait. Causal effect on $B_4$, the most pivotal trait in each subnetwork, represents the proportion of effect passed to each one of the four $B_4$ traits, for all traits with a non-zero vertical effect on $B_4$. High targeted confounder (darker shades) means that few variants (0.01%) have an effect on the confounder U, but with magnitude of effect rivaling direct effects. Low pervasive confounder (lighter shades) means that a large proportion (5%) of variants have an effect on the confounder, but with low magnitude. Traits $C$, $D$ and $E$ are simulated with high heritability (60%), while traits $A$ and $B$ are simulated with lower heritability (20%). **A)** The y-axis represents the precision which is the proportion of well-predicted variants among all predicted variants for a given label. **B)** The y-axis represents the recall, which is the proportion of well-predicted variants among all true variants for a given label. NB: In cases where no variant was clustered under a given label, it was impossible to calculate precision or recall; such instances are represented by squares below the x-axis.*

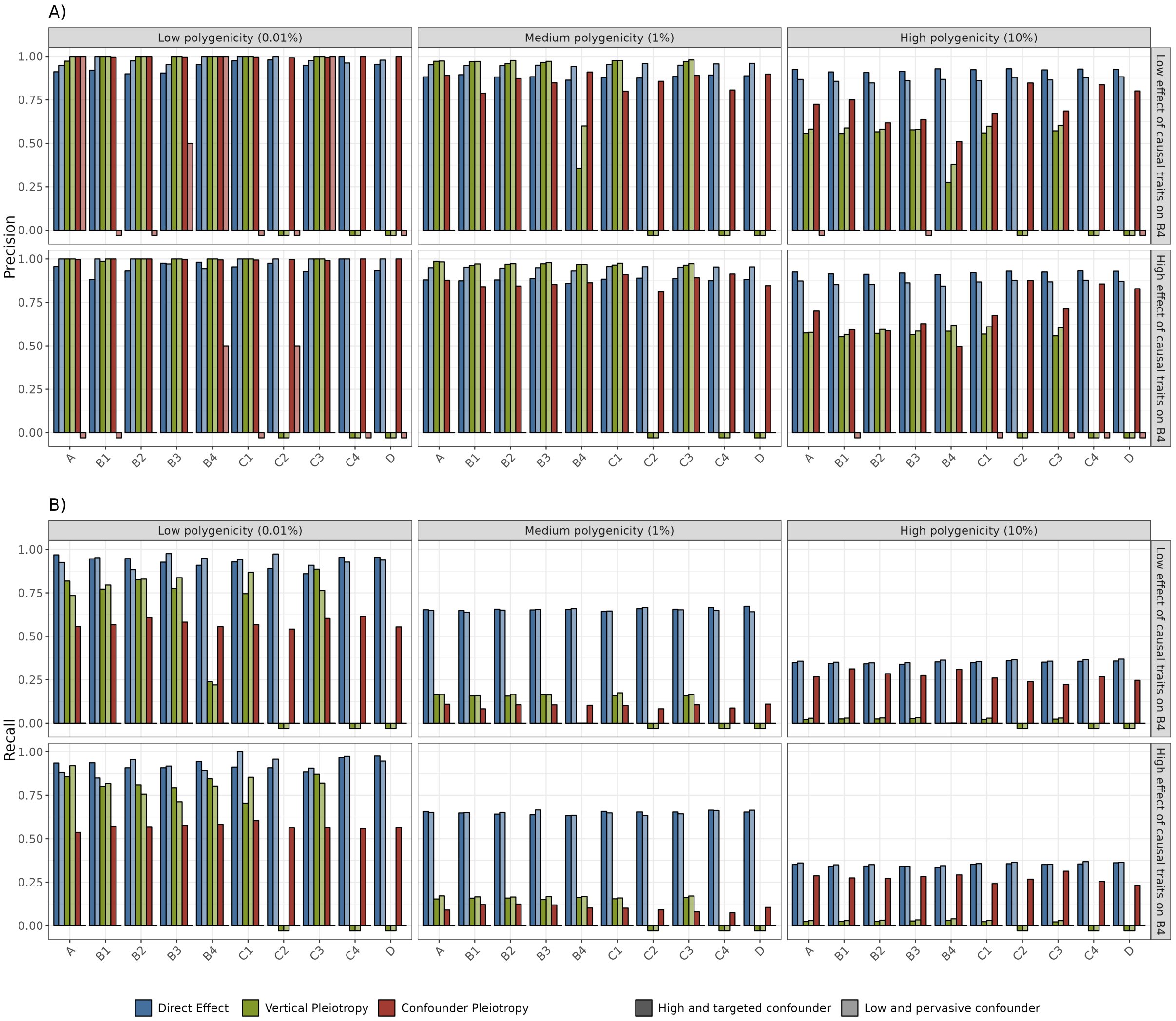


***Supplementary Figure 6. A)*** *Precision and* ***B)*** *Recall of PRISM predictions for significant variant-trait effects, on simulations, with lower trait heritability. The prioritized effects approach is used to compute precision and recall. The x-axis represents the simulated traits, grouped across subnetworks for visualization purpose (See Fig. 6 and Methods). Significant effects are defined with $P < 5 \times 10^{-8}/59$ , the recommended threshold from PRISM (See Methods). Bars are colored according to predicted labels: direct (blue), vertical pleiotropy (green) or confounder pleiotropy (red). Twelve scenarios are represented across facets, with varying parameters of polygenicity, strength of causal effect and type of confounder. Polygenicity represents the proportion of variants with a direct effect on each trait. Causal effect on $B_4$, the most pivotal trait in each subnetwork, represents the proportion of effect passed to each one of the four $B_4$ traits, for all traits with a non-zero vertical effect on $B_4$. High targeted confounder (darker shades) means that few variants (0.01%) have an effect on the confounder U, but with magnitude of effect rivaling direct effects. Low pervasive confounder (lighter shades) means that a large proportion (5%) of variants have an effect on the confounder, but with low magnitude. All traits are simulated with lower heritability (20%).* ***A)*** *The y-axis represents the precision which is the proportion of well-predicted variants among all predicted variants for a given label.* ***B)*** *The y-axis represents the recall, which is the proportion of well-predicted variants among all true variants for a given label. NB: In cases where no variant was clustered under a given label, it was impossible to calculate precision or recall; such instances are represented by squares below the x-axis.*

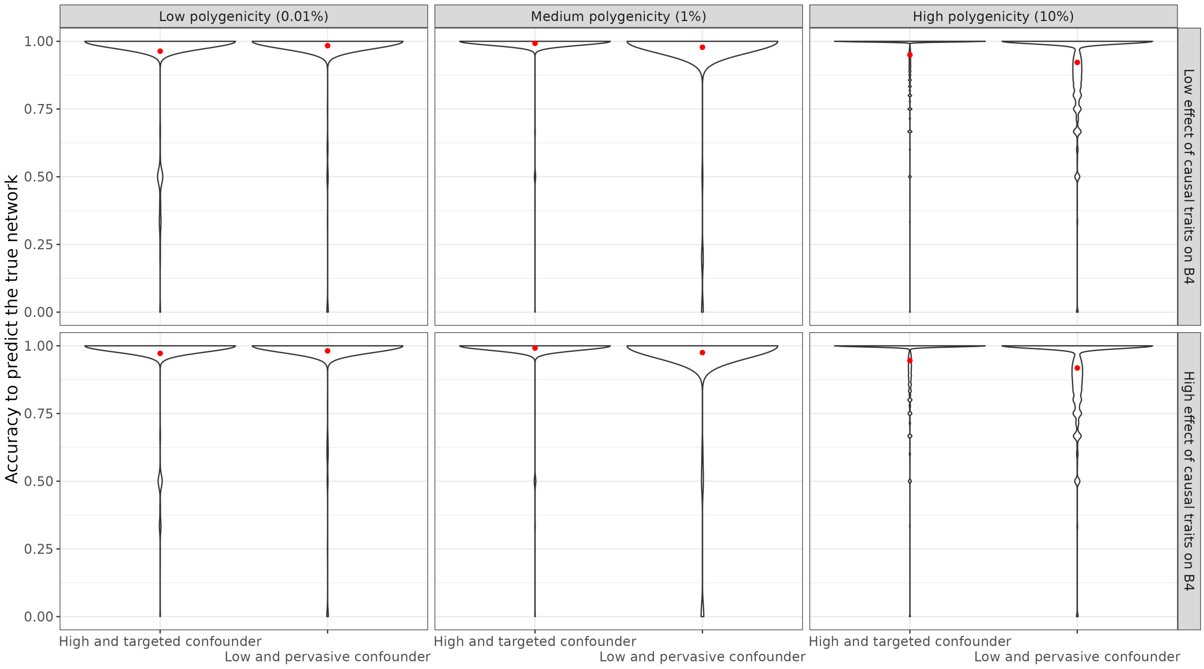


***Supplementary Figure 7.*** *Accuracy to predict the true network in simulations. Violin plot of the similarity between PRISM-predicted variant networks and true simulated networks, evaluated using the Simple Matching Coefficient (SMC), represented on the y-axis. The SMC measures the accuracy of the predicted networks by matching their edges with those of the true networks. A SMC of 1 indicates that all predicted edges are true edges, while a SMC of 0 indicates that none of the predicted edges correspond to the true edges. The x-axis displays the twelve simulated scenarios involving 60 traits with high heritability (60%), as detailed in Supplementary Table 1. The red dot represents the average SMC.*

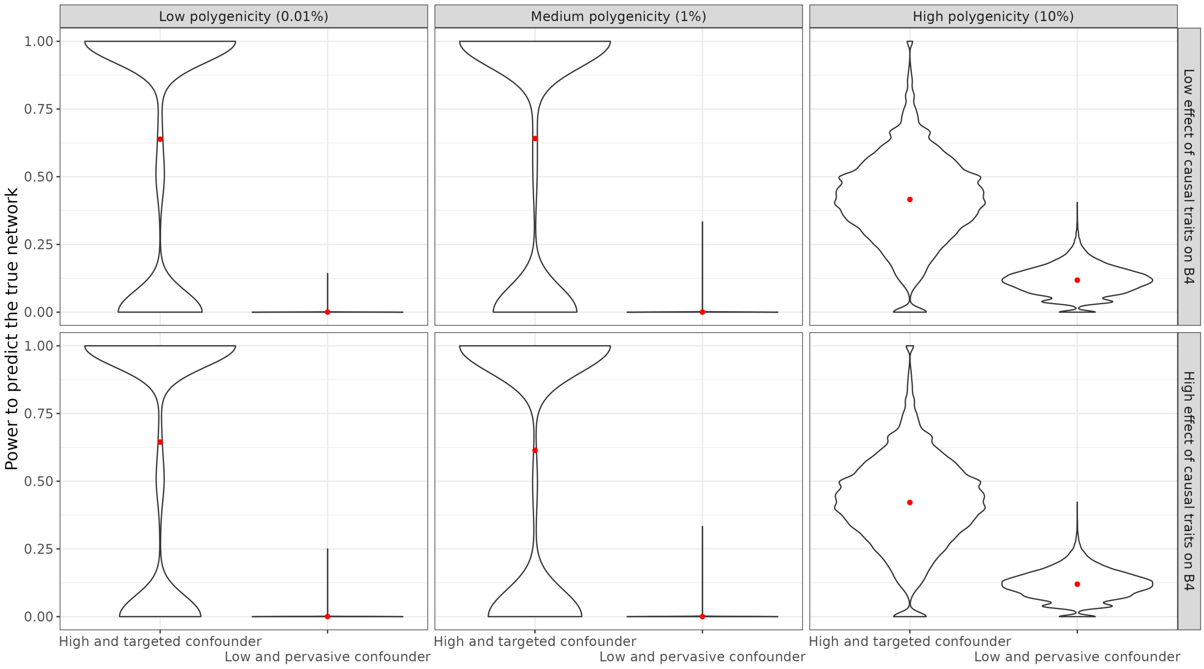


***Supplementary Figure 8.*** *Power to predict the true network in simulations. Violin plot of the similarity between true simulated networks and PRISM-predicted variant networks, evaluated using the Simple Matching Coefficient (SMC), represented on the y-axis. The SMC measures the power of detecting true networks by matching their edges with those of the predicted networks. A SMC of 1 indicates that all true edges are predicted, while a SMC of 0 indicates that none of the true edges are predicted. The x-axis displays the twelve simulated scenarios involving 60 traits with high heritability (60%), as detailed in Supplementary Table 1. The red dot represents the average SMC.*

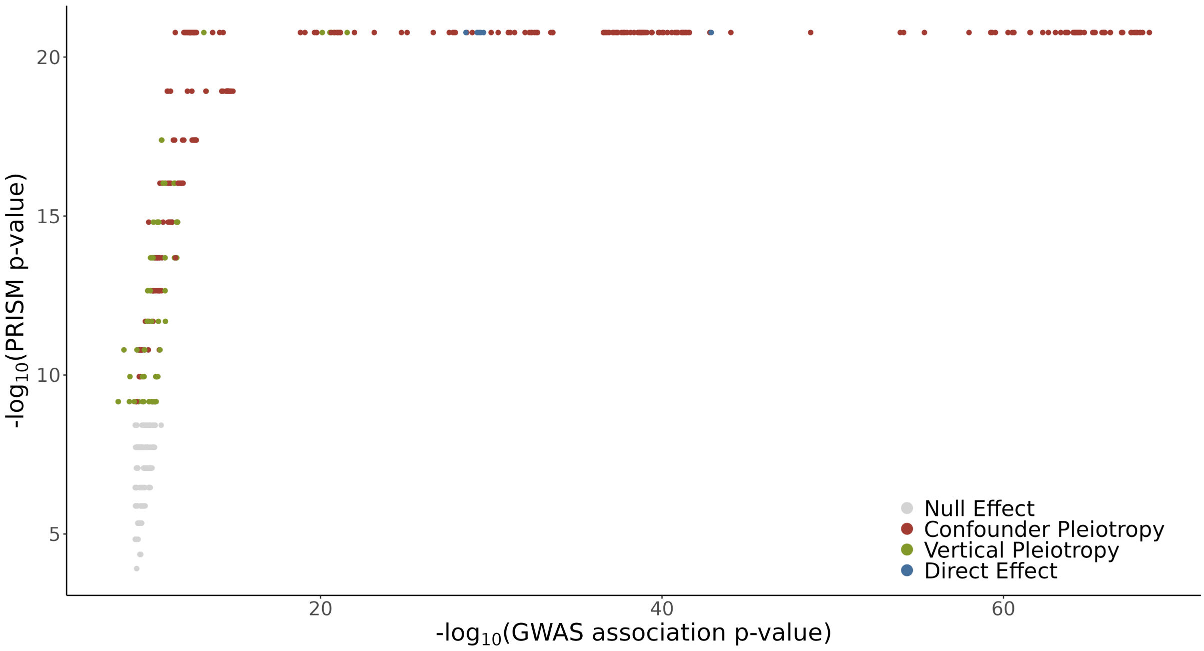


***Supplementary Figure 9.*** *P-values ($-log_{10}$) from the UK Biobank GWAS on coronary heart disease (CHD) (x-axis) and PRISM sign test (y-axis) for 910 genetic variants. Each dot represents a significant variant according to GWAS or PRISM, colored according to PRISM predicted labels: direct (blue), vertical (green) and confounder (red).*

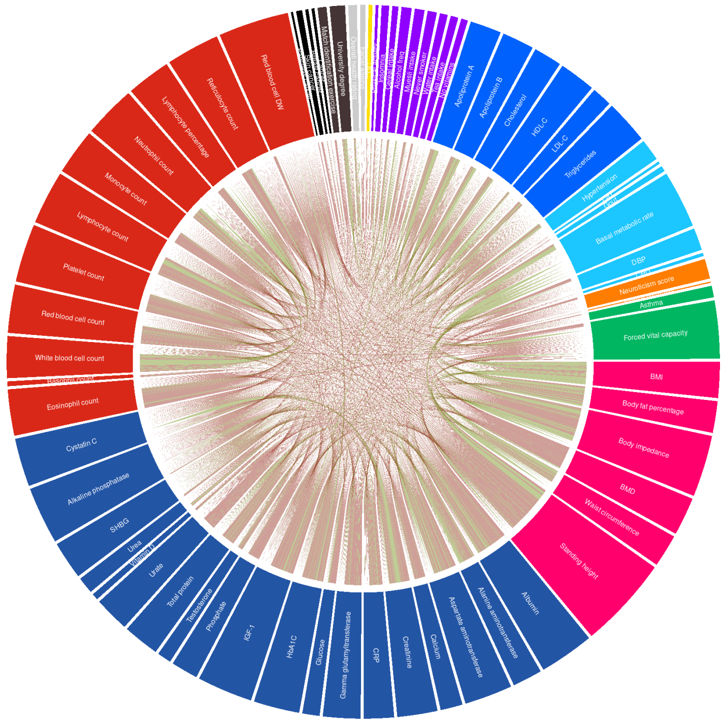


***Supplementary Figure 10.*** *Circos plot of shared variants, for all traits processed by PRISM. This represents genetic pleiotropy obtained from PRISM, with each line representing a significant variant with direct horizontal or pleiotropic effect on two traits according to PRISM. Blue lines represent horizontal pleiotropy, green lines represent vertical pleiotropy, red lines represent confounder pleiotropy.*

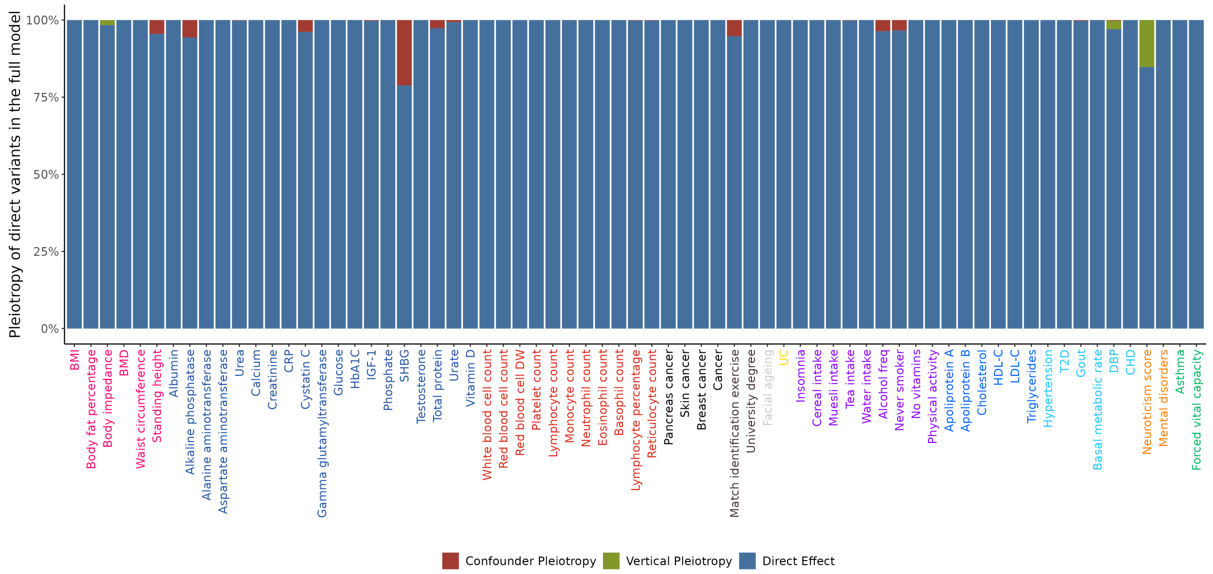


***Supplementary Figure 11.*** *PRISM predictions for all direct variant effects on Overall health rating identified in the full model, after removing each trait one by one. The y-axis shows how effects predicted as direct in the full model were classified in these incomplete models. The x-axis shows the trait removed. Significant effects are defined with* $P < 5 \times 10^{-8}/69$*, the recommended threshold from PRISM (See Methods). Bar colors indicate the classification group of the direct effects in the incomplete models.*

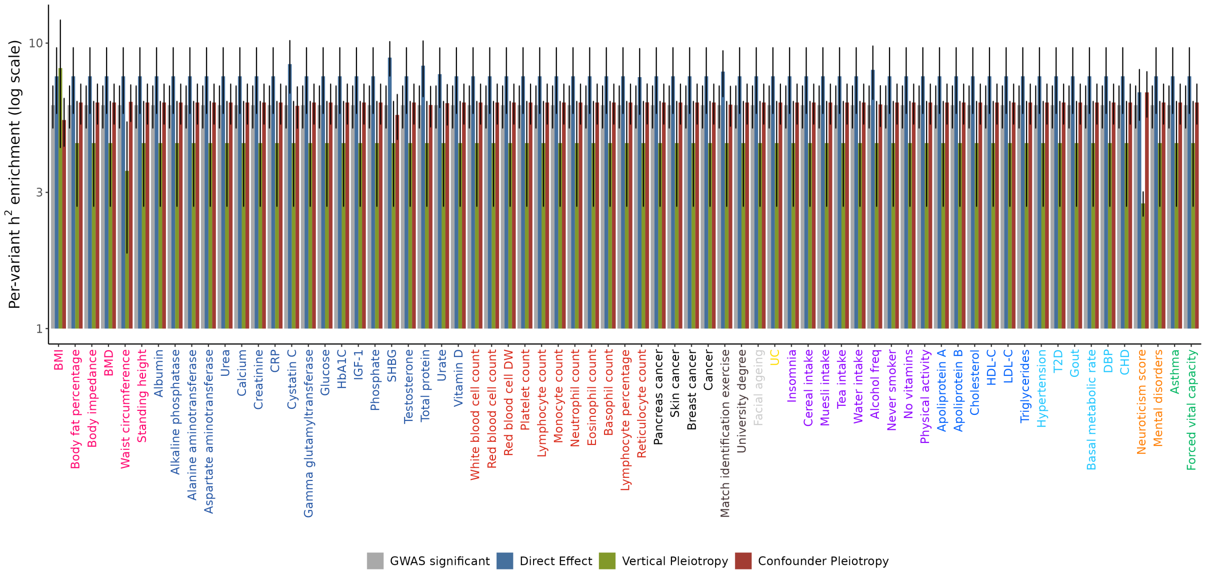


***Supplementary Figure 12.*** *Enrichment in per-variant heritability for variant effects on Overall health rationg, after removing each trait one by one. The x-axis shows the removed traits, color-coded by categories. The y-axis, presented on a logarithmic scale, indicates enrichment in per-variant heritability as estimated by stratified LD score regression, repeated 59 times (once per removed trait).*

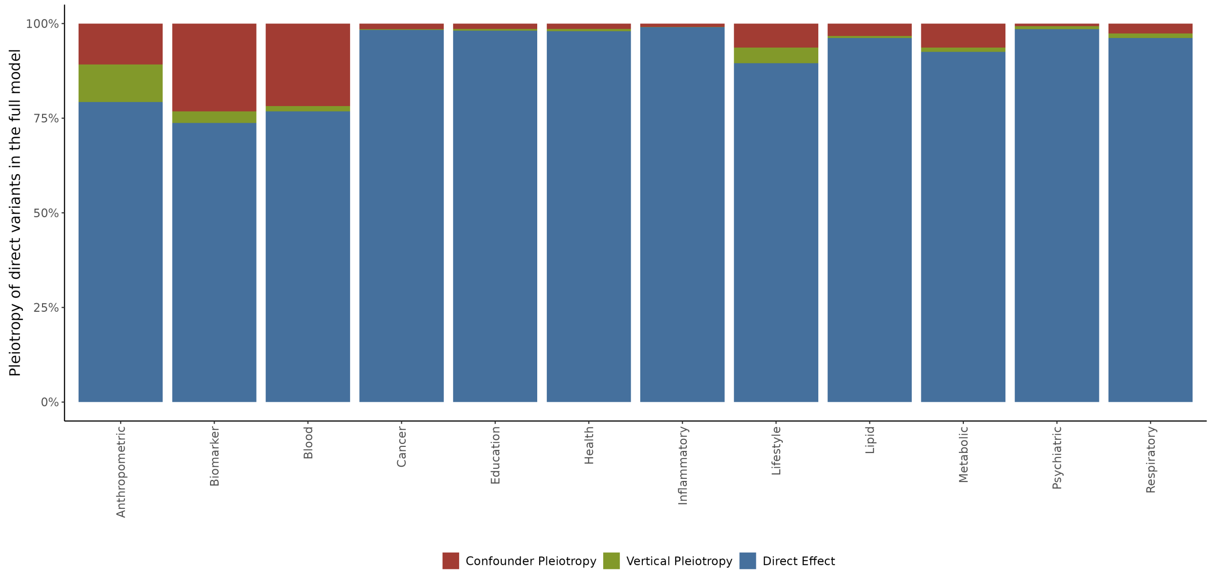


***Supplementary Figure 13.*** *PRISM predictions for all direct variant–trait effects identified in the full model, after removing all traits in each indicated category. The y-axis shows how effects predicted as direct in the full model were classified in these incomplete models. The x-axis shows the category of traits removed. Significant effects are defined with* $P < 5 \times 10^{-8}/69$*, the recommended threshold from PRISM (See Methods). Bar colors indicate the classification group of the direct effects in the incomplete models.*

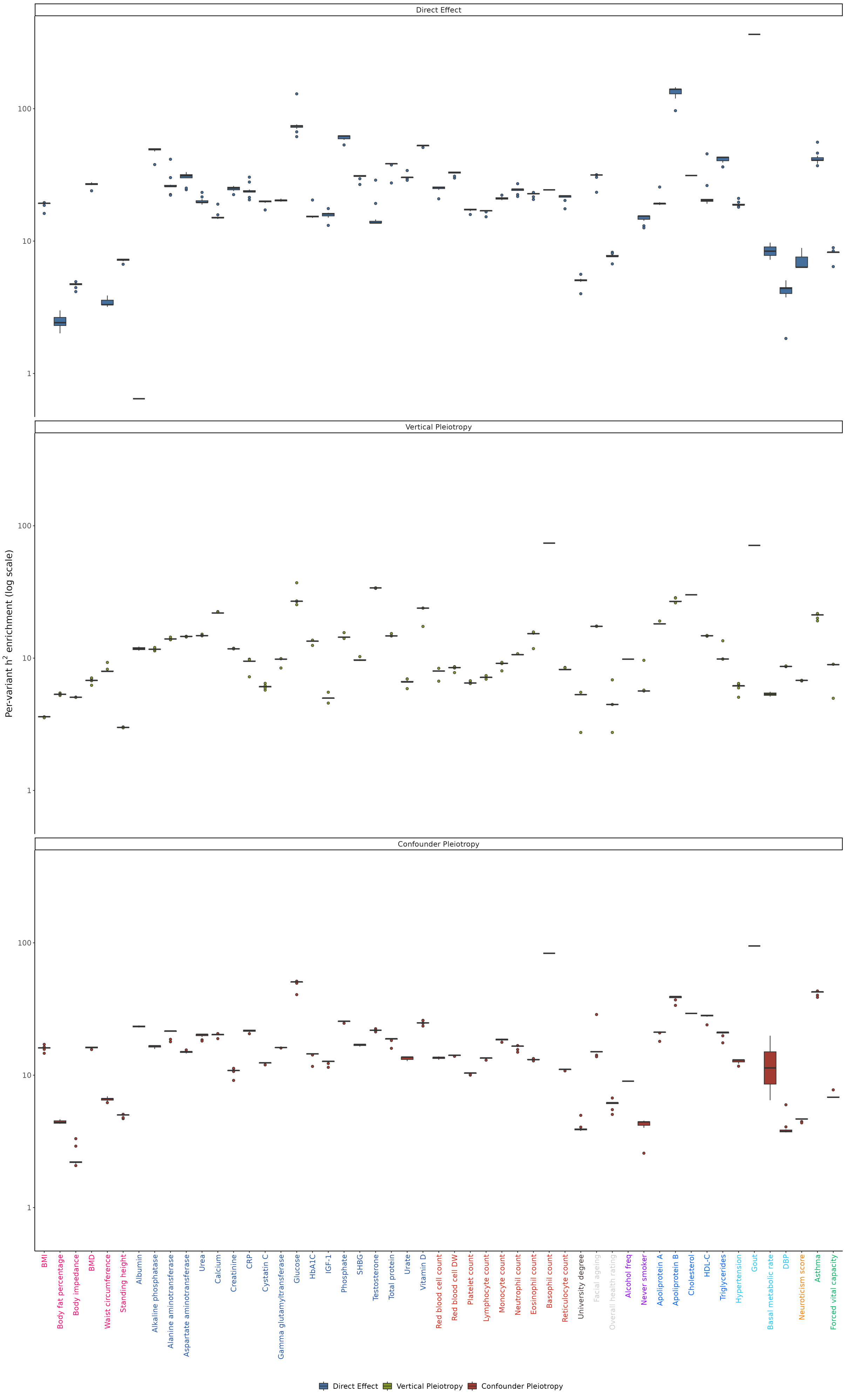

Direct Effect
Vertical Pleiotropy
Confounder Pleiotropy
Per-variant h² enrichment (log scale)
100
10
1
BMI
Body fat percentage
Body impedance
BMD
Waist circumference
Standing height
Albumin
Alkaline phosphatase
Alanine aminotransferase
Aspartate aminotransferase
Urea
Calcium
Creatinine
CRP
Cystatin C
Gamma glutamyltransferase
Glucose
HbA1C
IGF-1
Phosphate
SHBG
Testosterone
Total protein
Urate
Vitamin D
Red blood cell count
Red blood cell DW
Platelet count
Lymphocyte count
Monocyte count
Neutrophil count
Eosinophil count
Basophil count
Reticulocyte count
University degree
Facial ageing
Overall health rating
Alcohol freq
Never smoker
Apoliprotein A
Apoliprotein B
Cholesterol
HDL-C
Triglycerides
Hypertension
Gout
Basal metabolic rate
DBP
Neuroticism score
Asthma
Forced vital capacity
Direct Effect
Vertical Pleiotropy
Confounder Pleiotropy

***Supplementary Figure 14.*** *Enrichment in per-variant heritability for genetic variants from PRISM categories after removing all traits in each category. The x-axis shows the studied traits, color-coded by categories. The y-axis, presented on a logarithmic scale, indicates enrichment in per-variant heritability as estimated by stratified LD score regression, repeated 11 times (once per removed category). The three panels depict enrichment for direct, vertical, and confounder pleiotropy, respectively.*

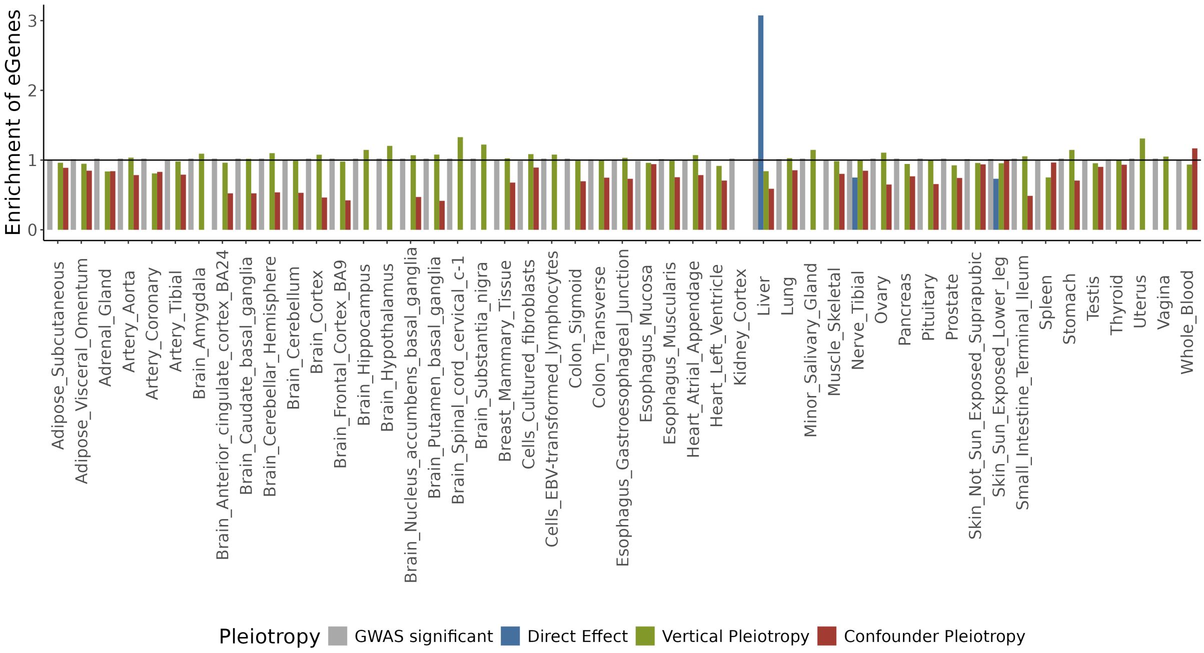


***Supplementary Figure 15.*** *Enrichment of eGenes, from different tissues, mapped to genetic variants, according to variants labels. The x-axis represents all tissues. The y-axis represents the enrichment of eGenes. The studied trait is 30710_irnt from UK Biobank, C-reactive protein. The eGenes were retrieved from GTEx v8. Enrichment peaks calculated from less than 5 genes were set to 0.*

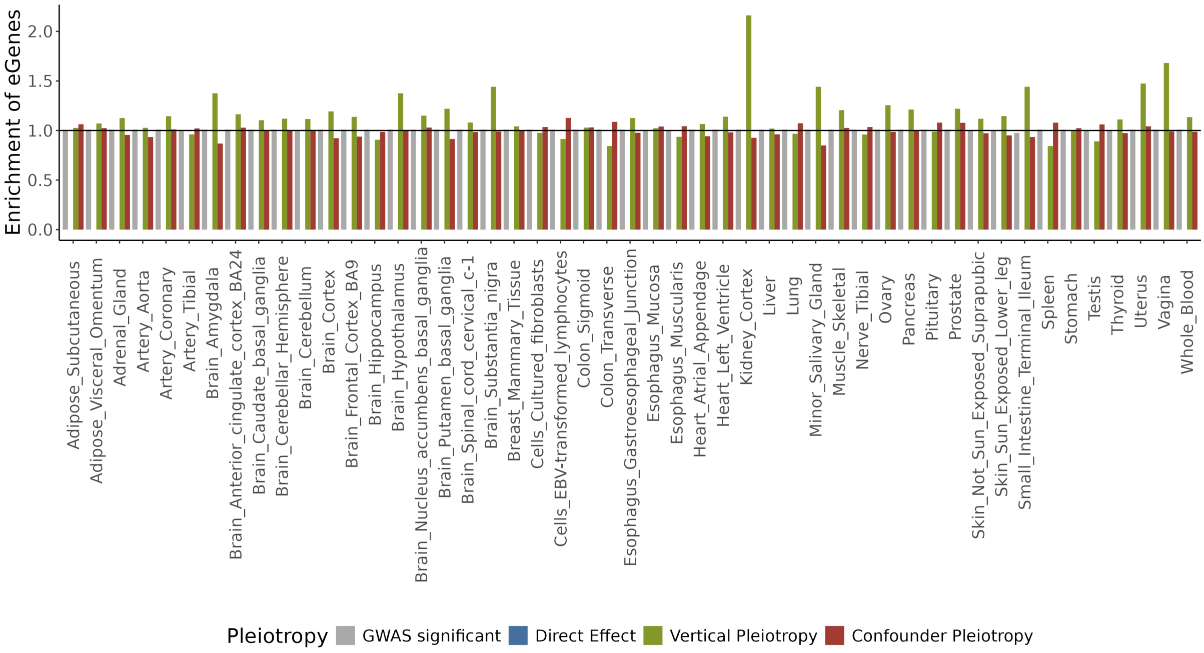


***Supplementary Figure 16.*** *Enrichment of eGenes, from different tissues, mapped to genetic variants, according to variants labels. The x-axis represents all tissues. The y-axis represents the enrichment of eGenes. The studied trait is 30600_irnt from UK Biobank, albumin. The eGenes were retrieved from GTEx v8. Enrichment peaks calculated from less than 5 genes were set to 0.*

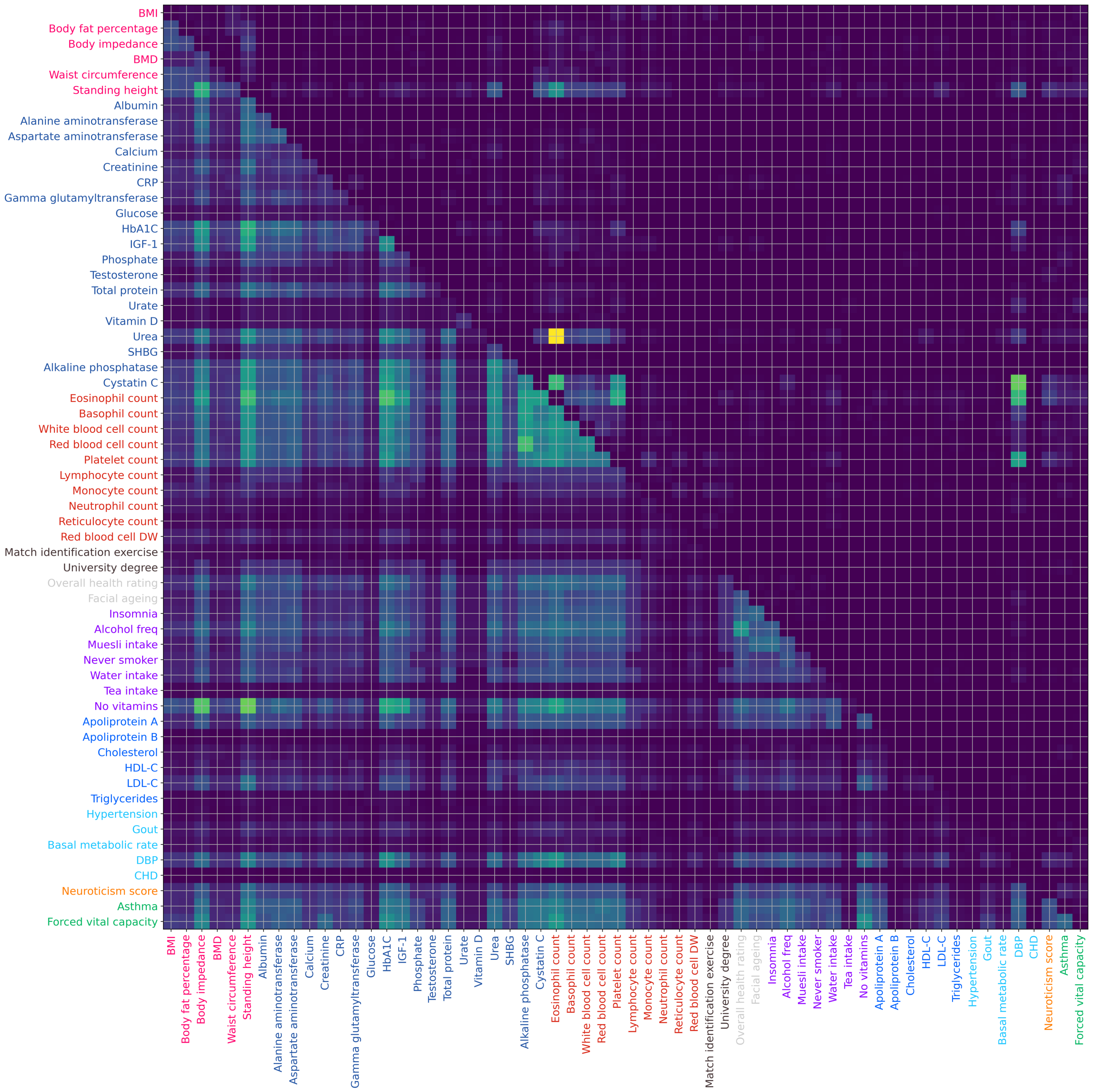


***Supplementary Figure 17.*** *Heatmap of shared genes between 60 traits with at least one direct variant. The bottom-left triangle represents common genes mapped from GWAS variants. The top-right triangle represents common genes mapped from PRISM direct-effect variants. Each tile is the intersection between two traits, and the color of the tile represents the number of common genes between those two traits divided by the total number of genes (PRISM or GWAS).*

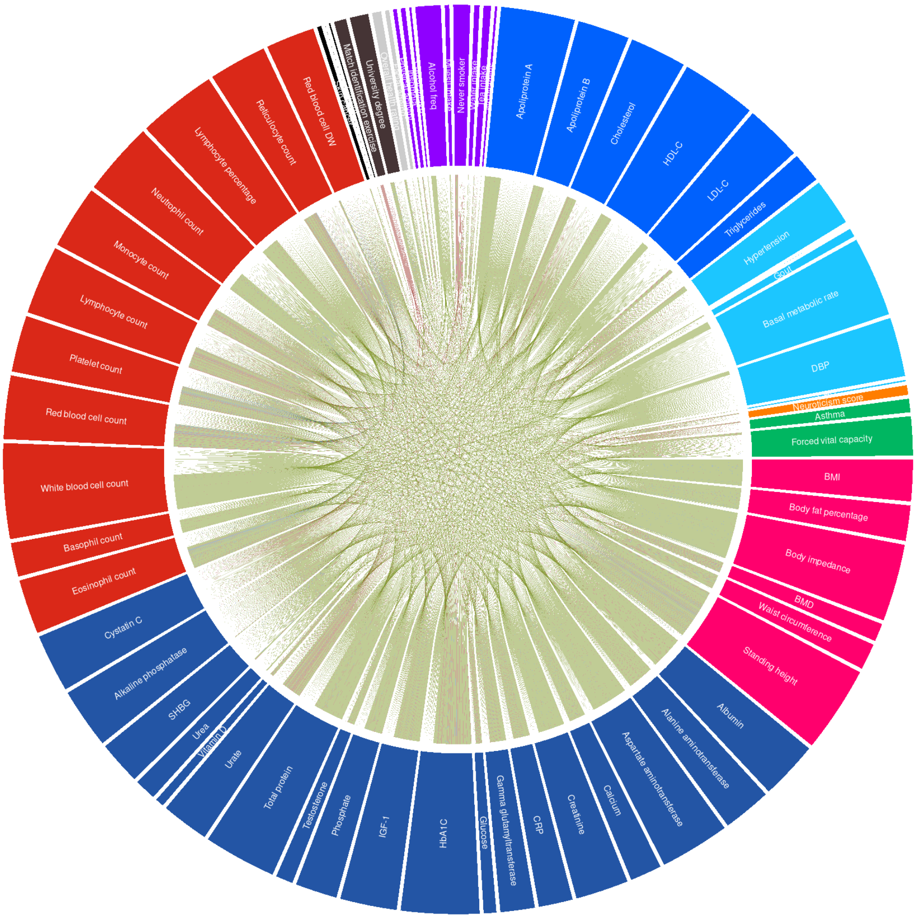


Supplementary Figure 18. *Circos plot of genes mapped to shared variants, for 67 traits from the UK Biobank. This represents genetic pleiotropy obtained from PRISM, with each line representing a gene mapped to a significant variant with direct horizontal or pleiotropic effect on two traits according to PRISM. Blue lines represent horizontal pleiotropy, green lines represent vertical pleiotropy, red lines represent confounder pleiotropy. To map a gene to a genetic variant, we prioritize first horizontal pleiotropy, then vertical pleiotropy, then confounder pleiotropy.*

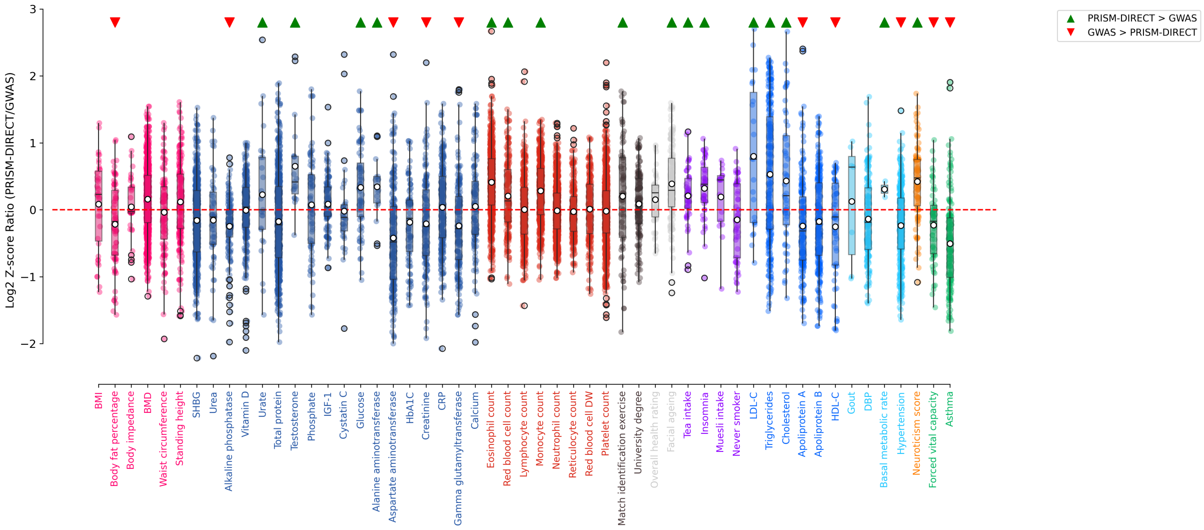


Supplementary Figure 19. *Z-score ratios for shared pathways enrichment between PRISM direct mapped genes and GWAS mapped genes. The x-axis represents all traits, colored by category. The y-axis represents a boxplot of Z-score ratios between PRISM direct and GWAS, from all pathways, in a log2 scale. A green triangle above a boxplot means that the log2 of the mean ratio is above* 0.2. *A red triangle above a boxplot means that the log2 of the mean ratio is below* 0.2.

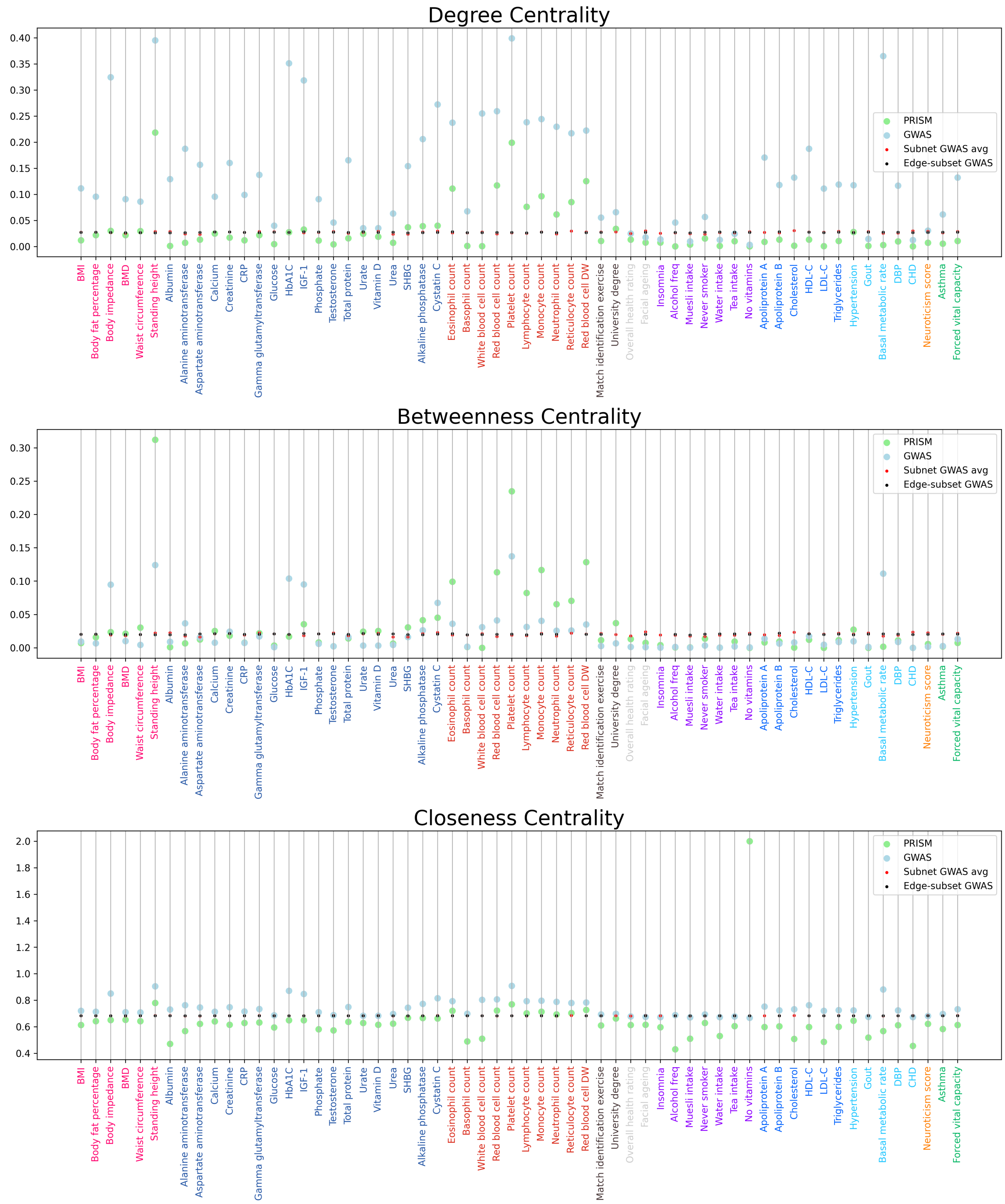


***Supplementary Figure 20.*** *Centrality measures of PRISM direct bipartite gene-trait network and GWAS bipartite gene-trait network. The three sub-plots show respectively the degree, the betweeness, and the closeness metrics, represented on the y-axis. The x-axis represents the 60 traits with at least one direct variant, colored by category, as the metrics are specific to a trait in the network. Green dots correspond to the PRISM direct network. Grey dots correspond to the GWAS network. Blue dots correspond to the average of multiple networks with randomly removed genes, to have the same number of genes as PRISM. Red dots correspond to the average of multiple networks with randomly removed genes and edges, to have the same number of genes and edges as PRISM.*

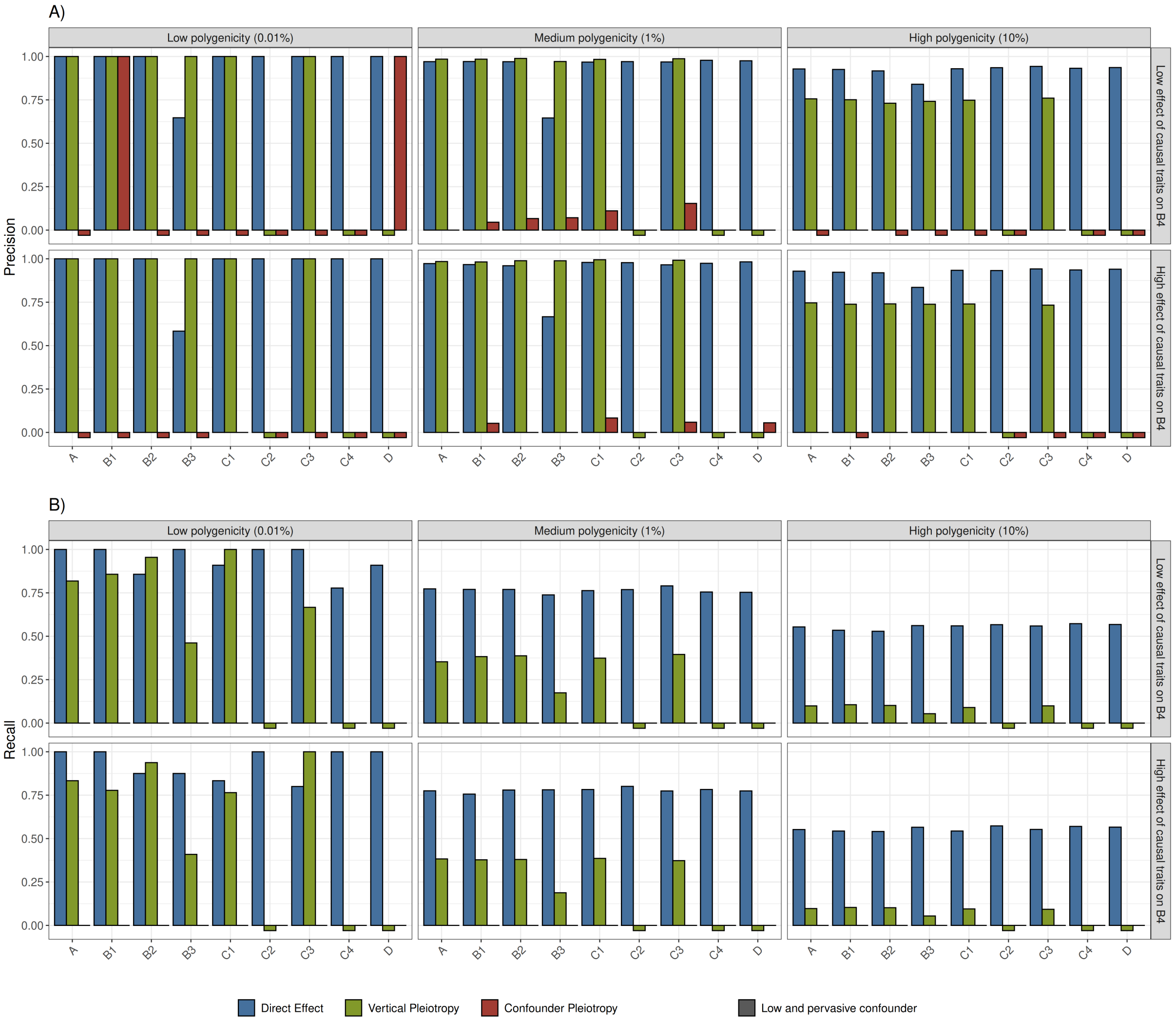


***Supplementary Figure 21. A)*** *Precision and* ***B)*** *Recall of PRISM predictions for significant variant-trait effects, on simulations, without $B_4^1$. The prioritized effects approach is used to compute precision and recall. The x-axis represents the simulated traits of the first subnetwork only (See Fig. 6 and Methods). Trait $B_4^1$ was used to simulate the network of traits, but was not included in PRISM. Significant effects are defined with $P < 5 \times 10^{-8}/59$ , the recommended threshold from PRISM (See Methods). Bars are colored according to predicted labels: direct (blue), vertical pleiotropy (green) or confounder pleiotropy (red). Six scenarios are represented across facets, with varying parameters of polygenicity and strength of causal effect. Polygenicity represents the proportion of variants with a direct effect on each trait. Causal effect on $B_4$, the most pivotal trait in each subnetwork, represents the proportion of effect passed to each one of the four $B_4$ traits, for all traits with a non-zero vertical effect on $B_4$. Low pervasive confounder means that a large proportion (5%) of variants have an effect on the confounder, but with low magnitude. All traits are simulated with high heritability (60%).* ***A)*** *The y-axis represents the precision which is the proportion of well-predicted variants among all predicted variants for a given label.* ***B)*** *The y-axis represents the recall, which is the proportion of well-predicted variants among all true variants for a given label. NB: In cases where no variant was clustered under a given label, it was impossible to calculate precision or recall; such instances are represented by squares below the x-axis.*

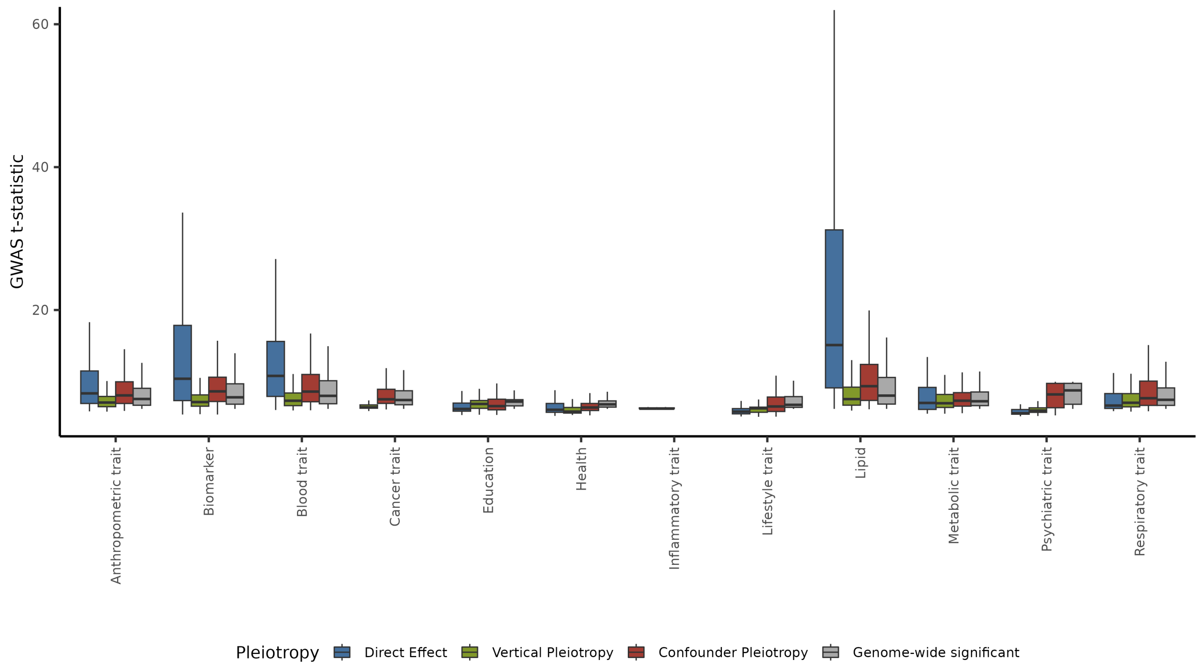


***Supplementary Figure 22.*** *Initial GWAS t-statistics of labeled genetic variants, grouped by labels and trait categories. The y-axis represents the initial t-statistics in GWAS summary statistics from UK Biobank. The x-axis represents traits grouped by broad categories. Boxplots are colored according to PRISM predicted labels, and GWAS significance at threshold* $P < 5 \times 10^{-8}/69$.

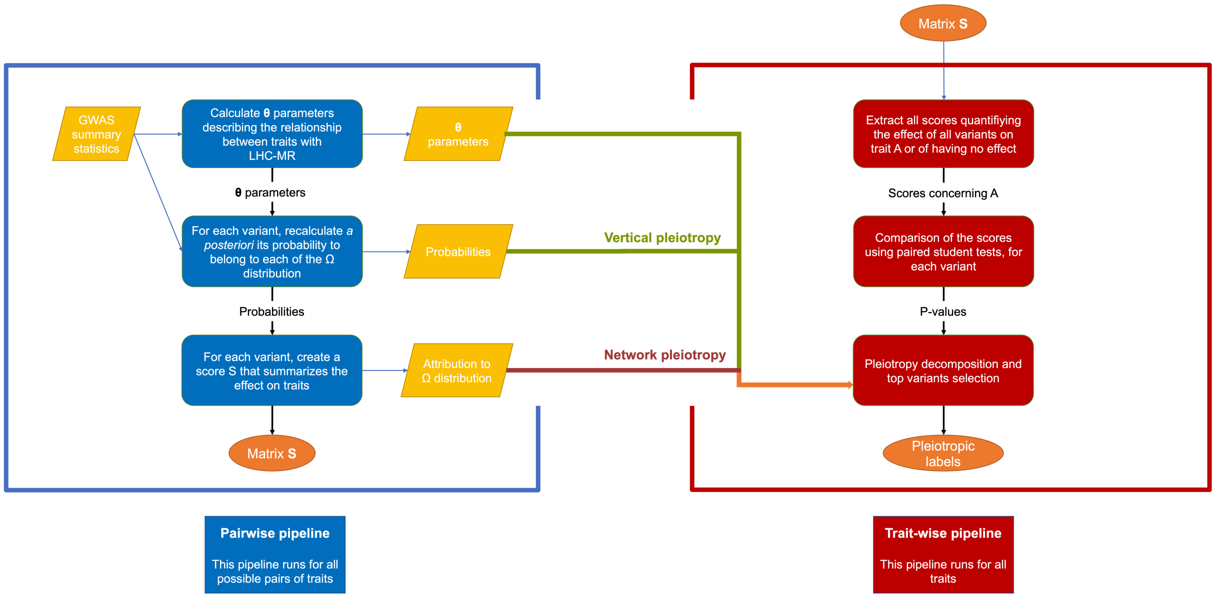


***Supplementary Figure 23.*** *The PRISM pipeline is divided in two main steps, pairwise and traitwise pipeline. The left side represents the pairwise pipeline, whereas the right side represents the traitwise pipeline.*

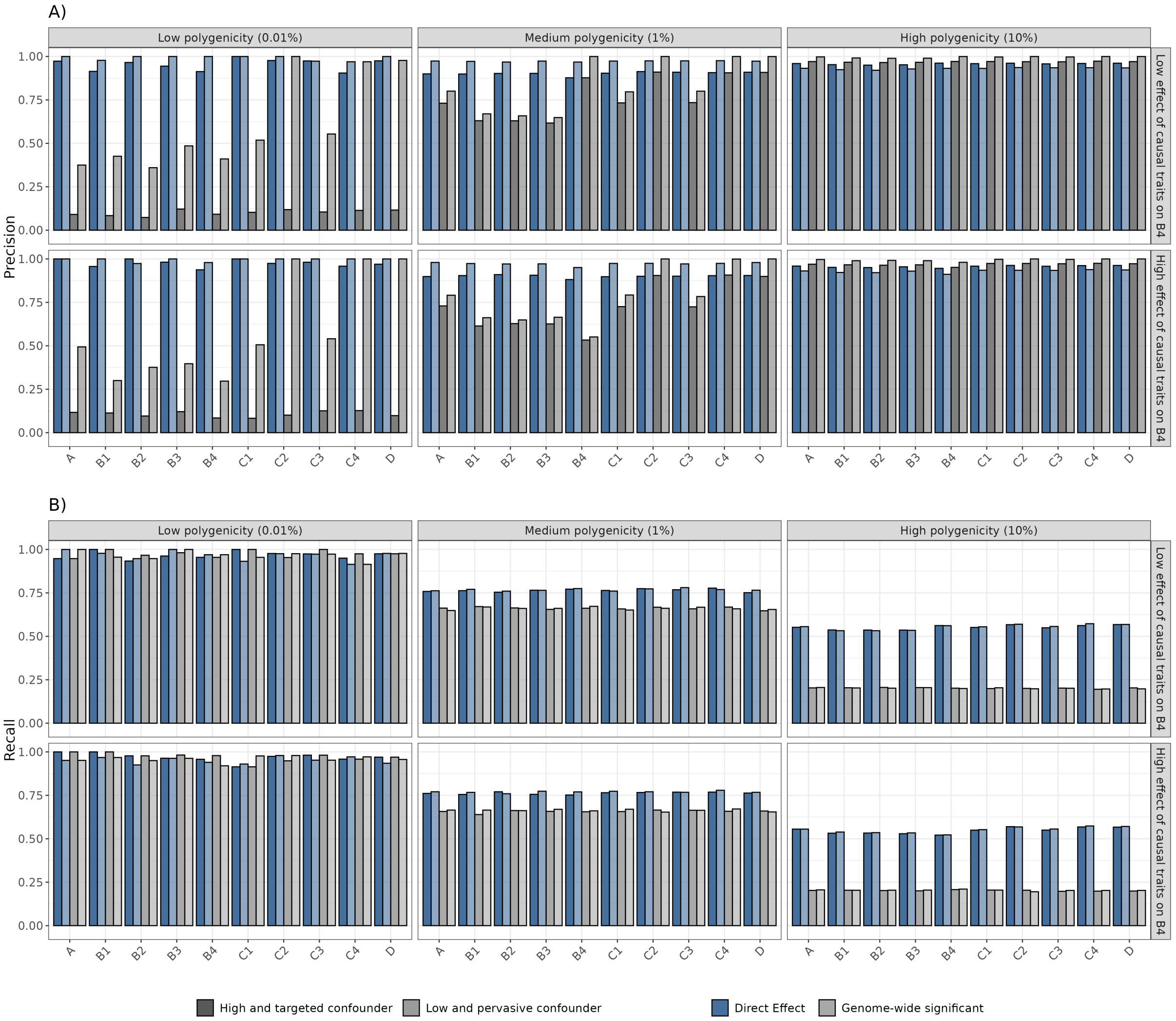


***Supplementary Figure 24. A)*** *Precision and* ***B)*** *Recall of PRISM and GWAS predictions for significant variant-trait effects, on simulations. The prioritized effects approach is used to compute precision and recall. The x-axis represents the simulated traits, grouped across subnetworks for visualization purpose (See Fig. 6 and Methods). Significant effects are defined with* $P < 5 \times 10^{-8}/59$ *, the recommended threshold from PRISM (See Methods). Bars are colored according to predicted labels: direct according to PRISM (blue), or direct according to GWAS-significance (grey) . Twelve scenarios are represented across facets, with varying parameters of polygenicity, strength of causal effect and type of confounder. Polygenicity represents the proportion of variants with a direct effect on each trait. Causal effect on* $B_4$*, the most pivotal trait in each subnetwork, represents the proportion of effect passed to each one of the four* $B_4$ *traits, for all traits with a non-zero vertical effect on* $B_4$*. High targeted confounder (darker shades) means that few variants (0.01%) have an effect on the confounder U, but with magnitude of effect rivaling direct effects. Low pervasive confounder (lighter shades) means that a large proportion (5%) of variants have an effect on the confounder, but with low magnitude. All traits are simulated with high heritability (60%).* ***A)*** *The y-axis represents the precision which is the proportion of well-predicted variants among all predicted variants for a given label.* ***B)*** *The y-axis represents the recall, which is the proportion of well-predicted variants among all true variants for a given label.*

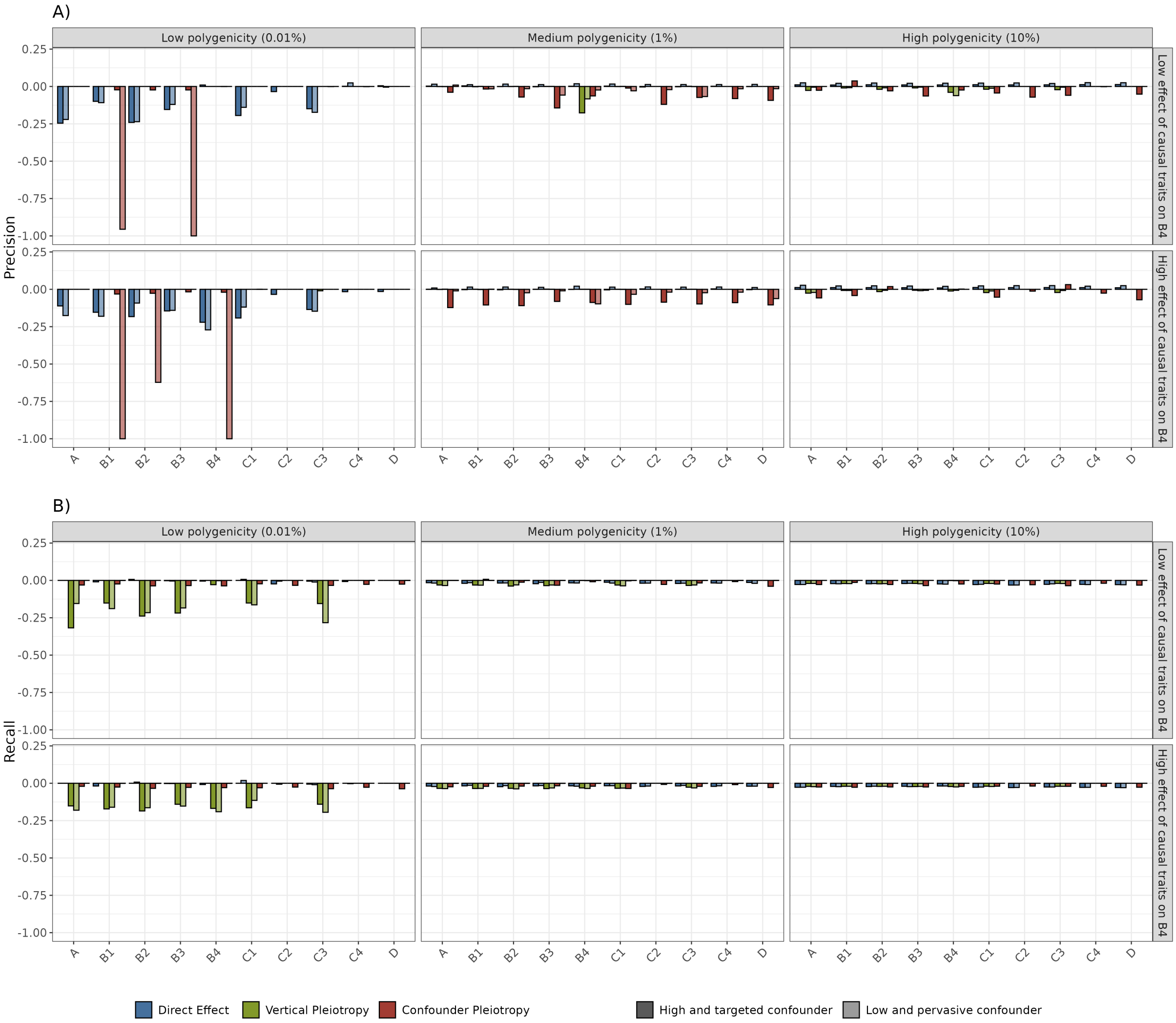


***Supplementary Figure 25.*** *Difference in* ***A)*** *Precision and* ***B)*** *Recall between PRISM predictions on simulations with or without noised global parameters. The prioritized effects approach is used to compute precision and recall. The x-axis represents the simulated traits, grouped across subnetworks and 10 replicates for visualization purpose (See Fig. 6, Methods and Supplementary Results). Significant effects are defined with* $P < 5 \times 10^{-8}/59$ *, the recommended threshold from PRISM (See Methods). Bars are colored according to predicted labels: direct (blue), vertical pleiotropy (green) or confounder pleiotropy (red). Twelve scenarios are represented across facets, with varying parameters of polygenicity, strength of causal effect and type of confounder. Polygenicity represents the proportion of variants with a direct effect on each trait. Causal effect on* $B_4$*, the most pivotal trait in each subnetwork, represents the proportion of effect passed to each one of the four* $B_4$ *traits, for all traits with a non-zero vertical effect on* $B_4$*. High targeted confounder (darker shades) means that few variants (0.01%) have an effect on the confounder U, but with magnitude of effect rivaling direct effects. Low pervasive confounder (lighter shades) means that a large proportion (5%) of variants have an effect on the confounder, but with low magnitude. All traits are simulated with high heritability (60%).* ***A)*** *The y-axis represents the precision which is the proportion of well-predicted variants among all predicted variants for a given label.* ***B)*** *The y-axis represents the recall, which is the proportion of well-predicted variants among all true variants for a given label.*

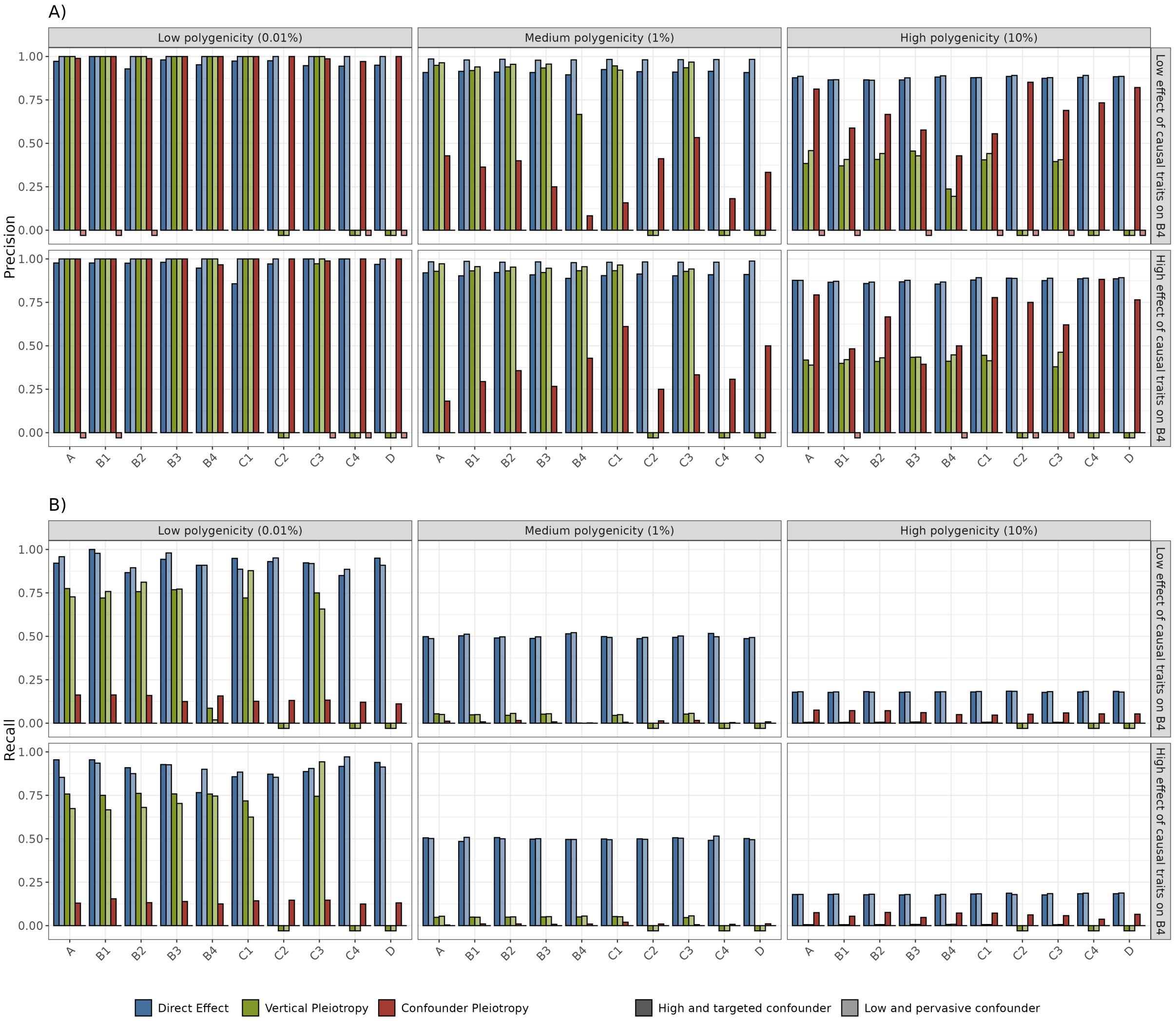


***Supplementary Figure 26. A)*** *Precision and* ***B)*** *Recall of PRISM predictions for significant variant-trait effects on simulations, with lower sample size. The prioritized effects approach is used to compute precision and recall. The x-axis represents the simulated traits, grouped across subnetworks for visualization purpose (See Fig. 6 and Methods). Significant effects are defined with $P < 5 \times 10^{-8}/59$ , the recommended threshold from PRISM (See Methods). Bars are colored according to predicted labels: direct (blue), vertical pleiotropy (green) or confounder pleiotropy (red). Twelve scenarios are represented across facets, with varying parameters of polygenicity, strength of causal effect and type of confounder. Polygenicity represents the proportion of variants with a direct effect on each trait. Causal effect on $B_4$, the most pivotal trait in each subnetwork, represents the proportion of effect passed to each one of the four $B_4$ traits, for all traits with a non-zero vertical effect on $B_4$. High targeted confounder (darker shades) means that few variants (0.01%) have an effect on the confounder U, but with magnitude of effect rivaling direct effects. Low pervasive confounder (lighter shades) means that a large proportion (5%) of variants have an effect on the confounder, but with low magnitude. All traits are simulated with high heritability (60%) and a sample size of 50,000.* ***A)*** *The y-axis represents the precision which is the proportion of well-predicted variants among all predicted variants for a given label.* ***B)*** *The y-axis represents the recall, which is the proportion of well-predicted variants among all true variants for a given label. NB: In cases where no variant was clustered under a given label, it was impossible to calculate precision or recall; such instances are represented by squares below the x-axis.*

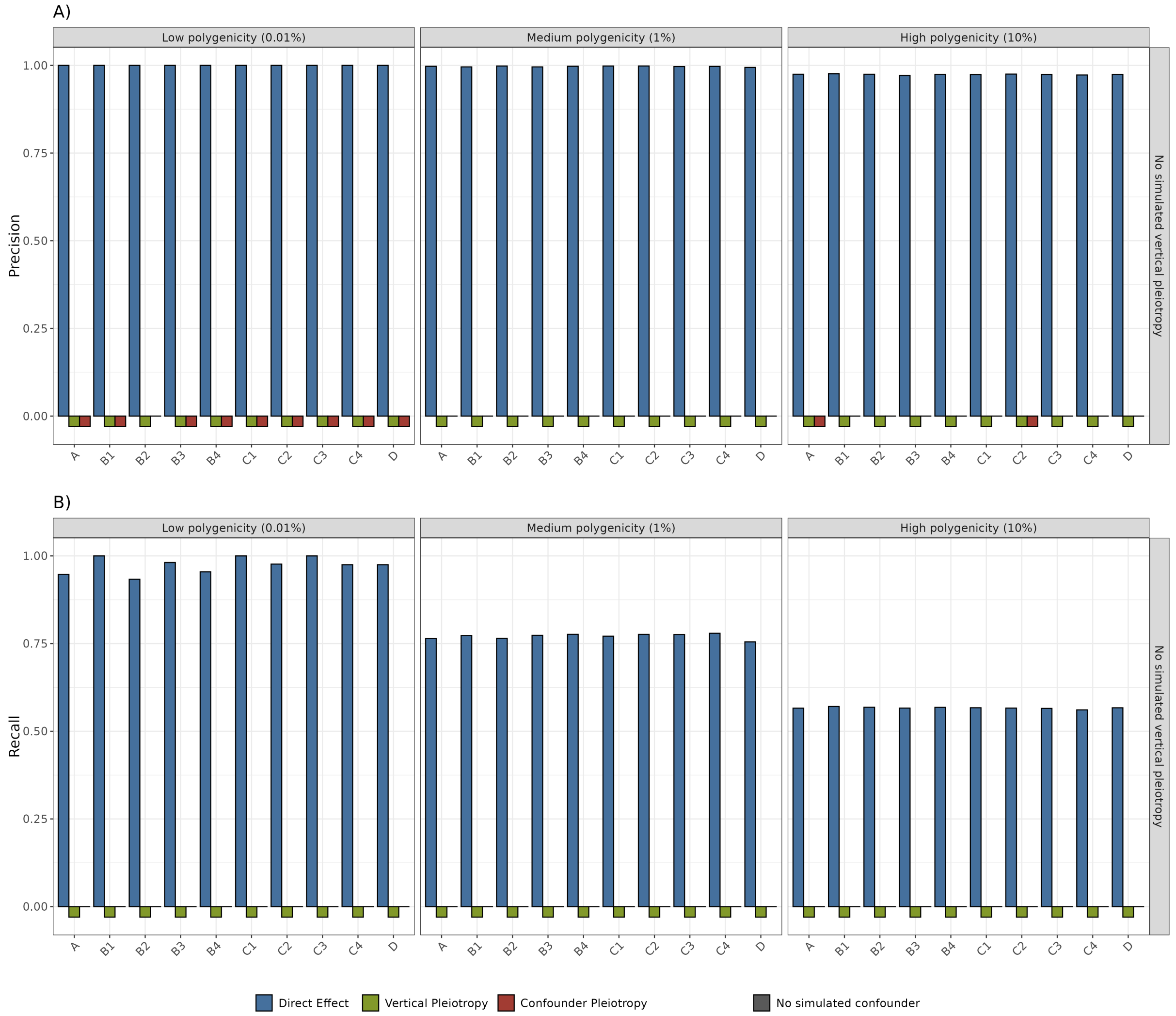


***Supplementary Figure 27. A)*** *Precision and* ***B)*** *Recall of PRISM predictions for significant variant-trait effects simulated without any vertical or confounder pleiotropy. The prioritized effects approach is used to compute precision and recall. The x-axis represents the simulated traits, grouped across subnetworks for visualization purpose (See Fig. 6 and Methods). Significant effects are defined with* $P < 5 \times 10^{-8}/59$ *, the recommended threshold from PRISM (See Methods). Bars are colored according to predicted labels: direct (blue), vertical pleiotropy (green) or confounder pleiotropy (red). Three scenarios are represented across facets, with varying parameters of polygenicity. Polygenicity represents the proportion of variants with a direct effect on each trait. All traits are simulated with high heritability (60%).* ***A)*** *The y-axis represents the precision which is the proportion of well-predicted variants among all predicted variants for a given label.* ***B)*** *The y-axis represents the recall, which is the proportion of well-predicted variants among all true variants for a given label. NB: In cases where no variant was clustered under a given label, it was impossible to calculate precision or recall; such instances are represented by squares below the x-axis.*

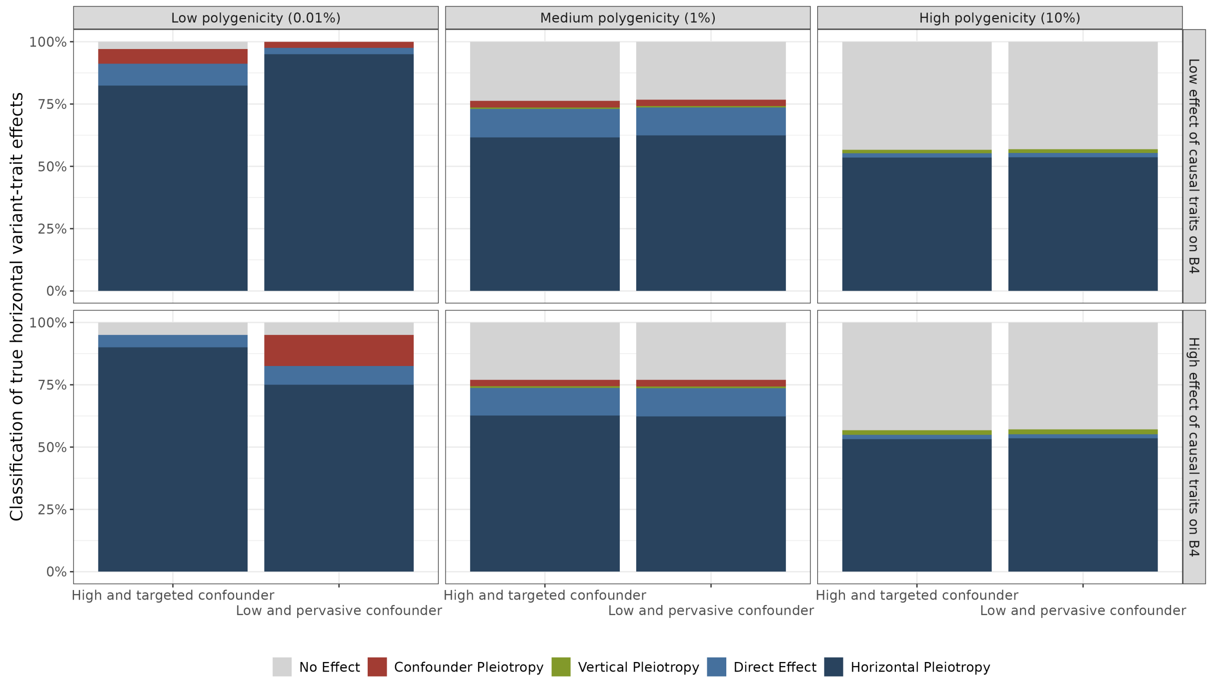


***Supplementary Figure 28.*** *PRISM predictions for true horizontal variant-trait effects, on simulations. The y-axis represents the proportion of prediction in each group, in different conditions. The x-axis represents the confounder effects. Each bar represents a scenario, with variant-trait predictions merged for each trait. Significant effects are defined with* $P < 5 \times 10^{-8}/59$*, the recommended threshold from PRISM (See Methods). Bars are colored according to the group the horizontal effect was predicted as. Twelve scenarios are represented across facets, with varying parameters. Polygenicity represents the proportion of variants with a direct effect on each trait. Effect on* $B_4$*, the most pivotal trait, represents the proportion of effect passed to* $B_4$*, for all traits with a non-zero vertical effect on* $B_4$*. High targeted confounder means that few variants (0.01%) have an effect on the confounder U, but with magnitude of effect rivaling direct effects. Low pervasive confounder means that a large proportion (5%) of variants have an effect on the confounder, but with low magnitude.*

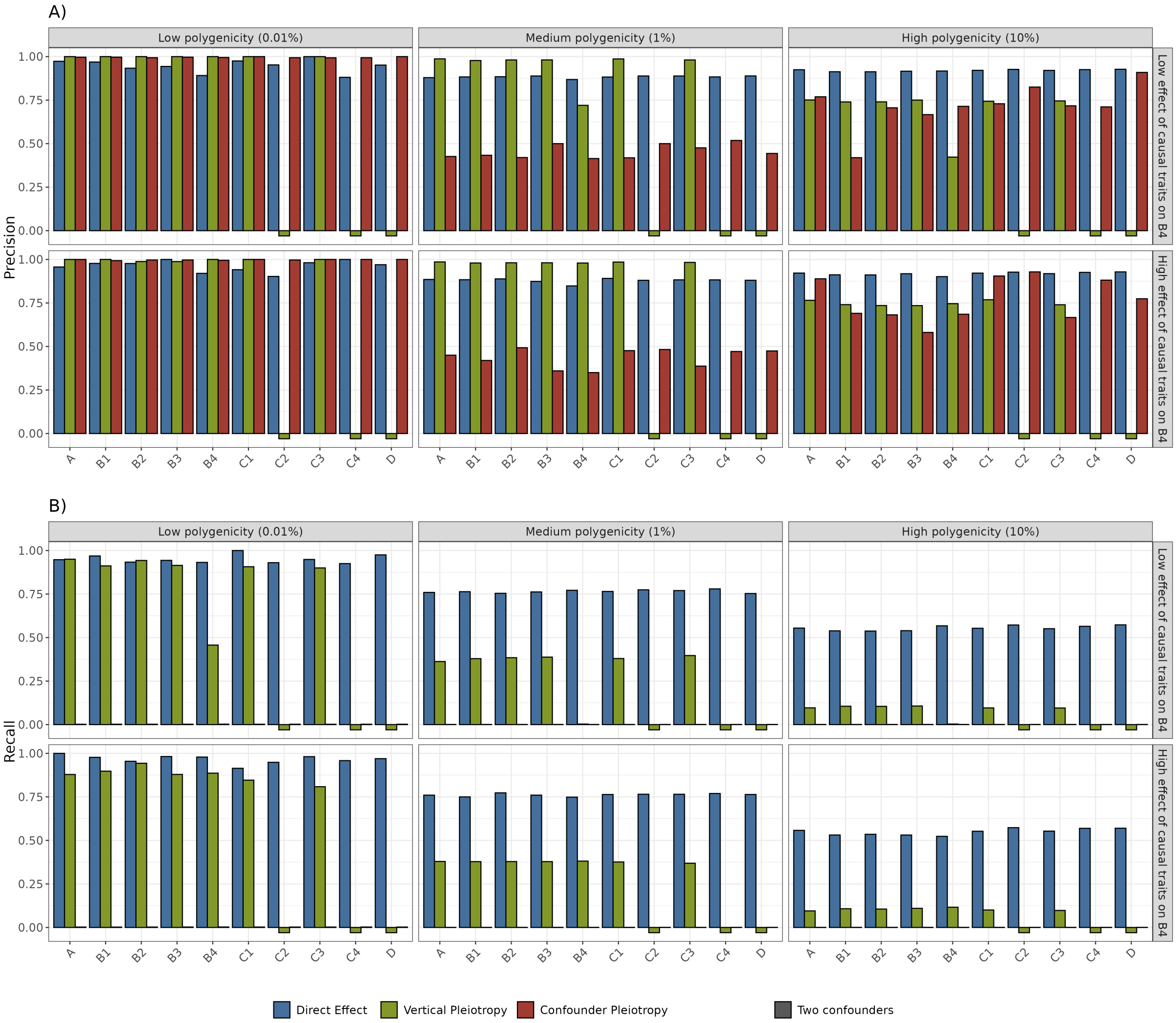


***Supplementary Figure 29. A)*** *Precision and* ***B)*** *Recall of PRISM predictions for significant variant-trait effects on simulations, with two simulated confounders. The prioritized effects approach is used to compute precision and recall. The x-axis represents the simulated traits, grouped across subnetworks for visualization purpose (See Fig. 6 and Methods). Significant effects are defined with* $P < 5 \times 10^{-8}/59$ *, the recommended threshold from PRISM (See Methods). Bars are colored according to predicted labels: direct (blue), vertical pleiotropy (green) or confounder pleiotropy (red). Six scenarios are represented across facets, with varying parameters of polygenicity and strength of causal effect. Polygenicity represents the proportion of variants with a direct effect on each trait. Causal effect on* $B_4$*, the most pivotal trait in each subnetwork, represents the proportion of effect passed to each one of the four* $B_4$ *traits, for all traits with a non-zero vertical effect on* $B_4$*. Two confounders means that few variants (0.01%) have an effect on the confounder U with magnitude of effect rivaling direct effects, and simultaneously that a large proportion (5%) of variants have an effect on the confounder with low magnitude. All traits are simulated with high heritability (60%).* ***A)*** *The y-axis represents the precision which is the proportion of well-predicted variants among all predicted variants for a given label.* ***B)*** *The y-axis represents the recall, which is the proportion of well-predicted variants among all true variants for a given label. NB: In cases where no variant was clustered under a given label, it was impossible to calculate precision or recall; such instances are represented by squares below the x-axis.*

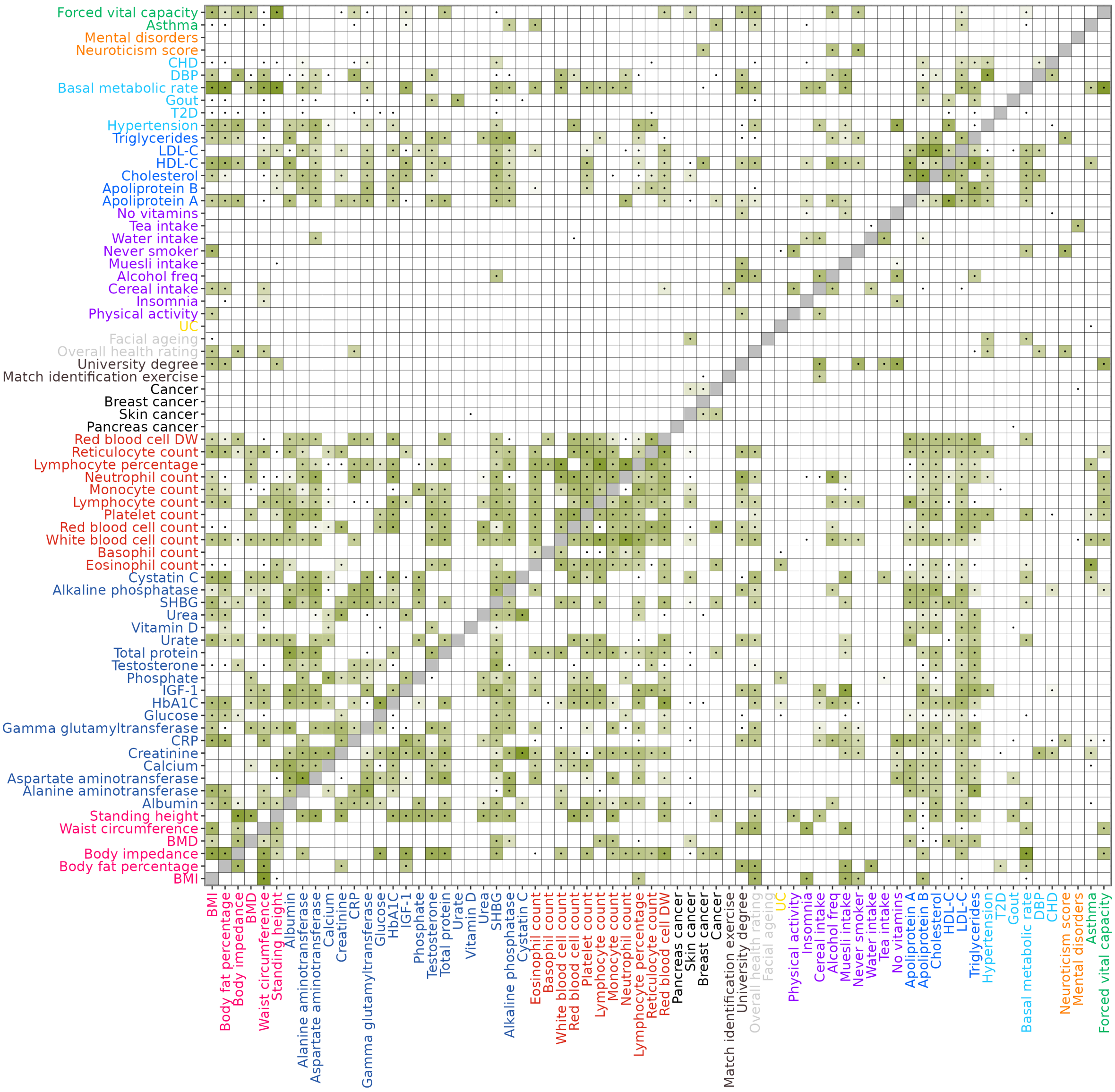


***Supplementary Figure 30.*** *Heatmap of trait relationships, measured by PRISM and LHC-MR. Each cell corresponds to the effect of the trait of the x-axis (exposure) on the trait of the y-axis (outcome). The green color gradient indicates the vertical effect, scaled according to the number of PRISM predicted vertical variant-trait effects. Dots mark significant effects identified by LHC-MR ($P < 0.05/69$). Diagonal cells are shaded grey because the effect of a trait on itself is not displayed.*

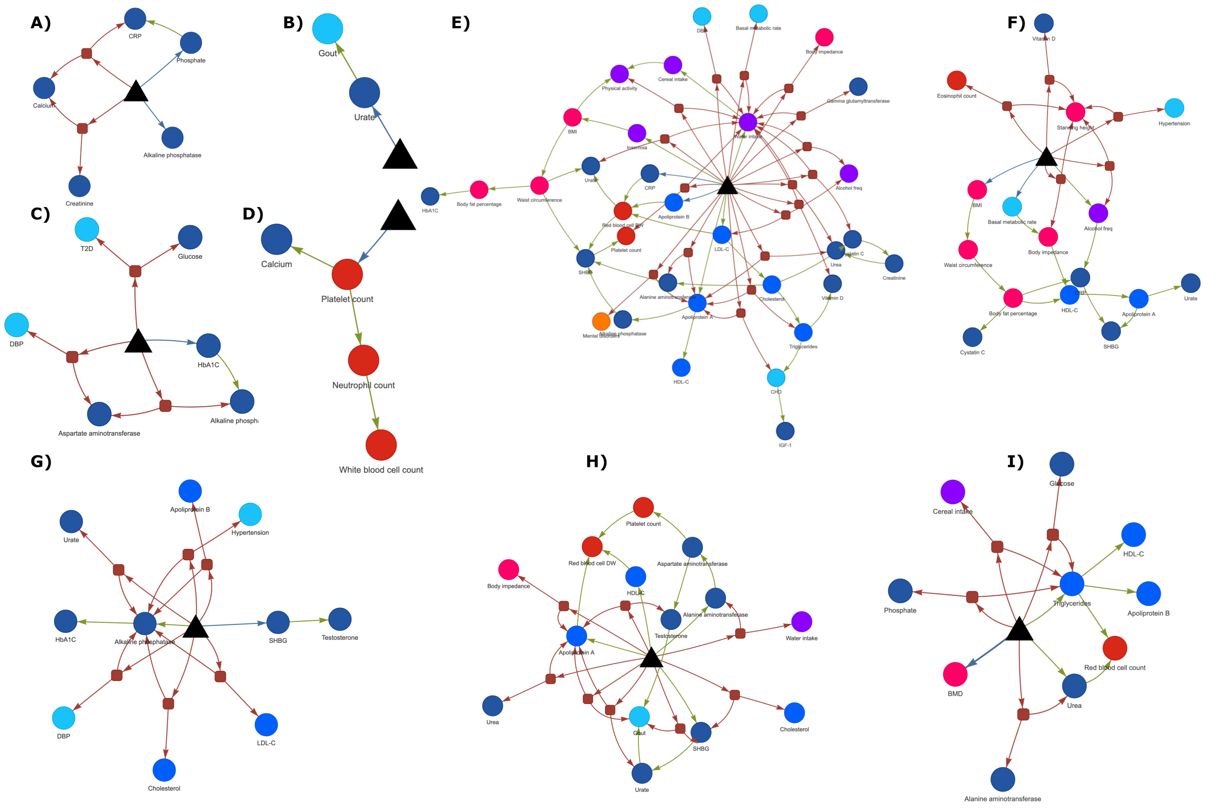


***Supplementary Figure 31.*** *PRISM inferred network for a panel of validated genetic variants. Genetic variants are represented as black triangles. Red arrows are effects of the variant through a confounder meaning confounder pleiotropy. Green arrows are effects of variant through a causal trait meaning vertical pleiotropy. Blue arrows are direct causal effects from the variant to traits. Confounders are represented as red squares and traits are represented as circles colored according to trait categories.*

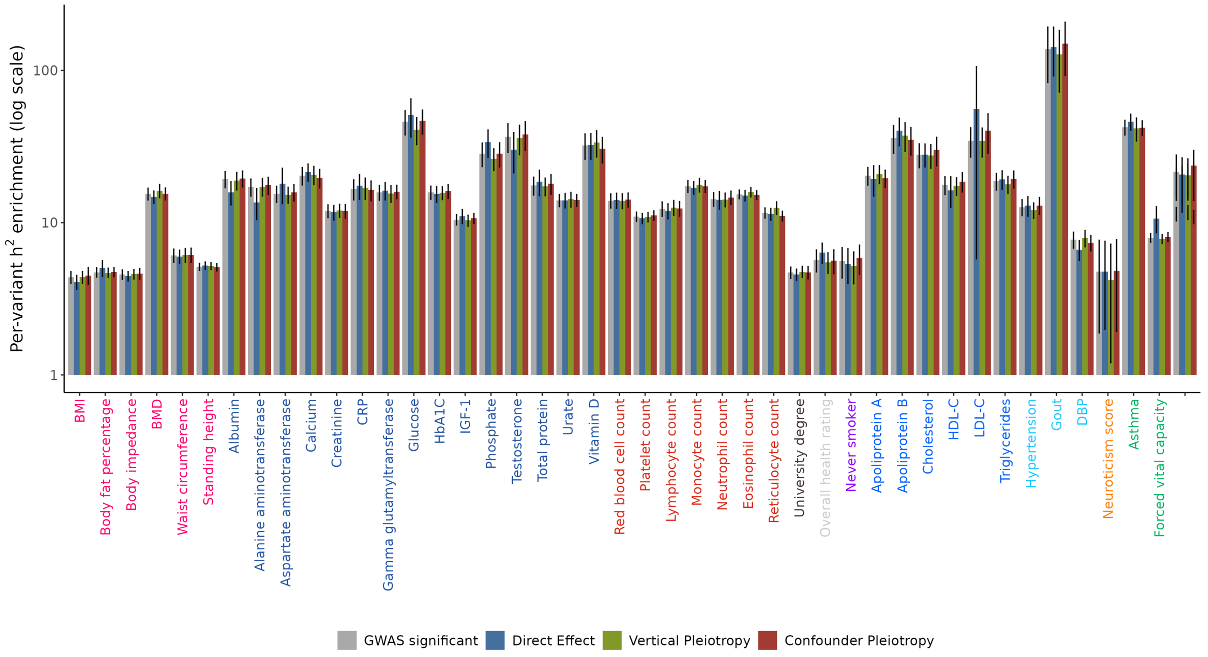


***Supplementary Figure 32.*** *Enrichment in per-variant heritability of genetic variants, according to randomized PRISM categories (direct, vertical, confounder) and GWAS. The x-axis represents the traits colored by broad categories. The y-axis represents the enrichment in per-variant heritability, estimated using stratified LD score regression and plotted on a logarithmic scale. Error bars represent the standard error of each enrichment estimate, also derived from stratified LD score regression. PRISM labels were randomly shuffled while preserving the number of variants in each category.*

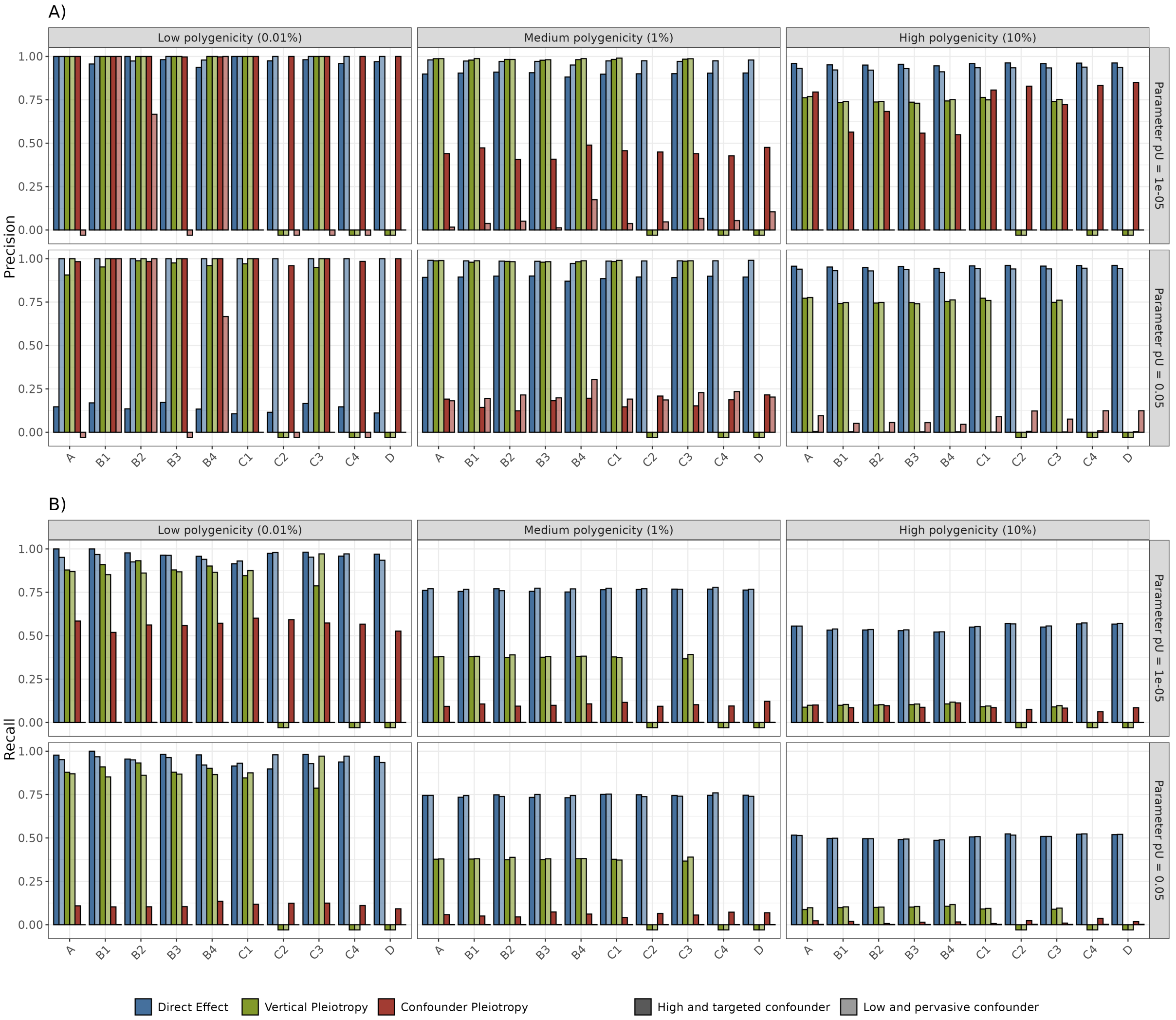


***Supplementary Figure 33. A)*** *Precision and* ***B)*** *Recall of PRISM predictions for significant variant-trait effects, on simulations, with varying value of pU parameter. The prioritized effects approach is used to compute precision and recall. The x-axis represents the simulated traits, grouped across subnetworks for visualization purpose (See Fig. 6 and Methods). For example, trait $A$ represents variant-trait effects on all traits $A^1$, $A^2$, $A^3$ and $A^4$. Significant effects are defined with $P < 5 \times 10^{-8}/59$, the recommended threshold from PRISM (See Methods). Bars are colored according to predicted labels: direct (blue), vertical pleiotropy (green) or confounder pleiotropy (red). Twelve scenarios are represented across facets, with varying parameters of polygenicity, strength of causal effect and type of confounder. Polygenicity represents the proportion of variants with a direct effect on each trait. Causal effect (the proportion of effect passed from one trait in vertical pleiotropy to the other trait) is set to 30%. Parameter pU represents PRISM chosen value for latent parameter $\pi_u$. High targeted confounder (darker shades) means that few variants (0.01%) have an effect on the confounder U, but with magnitude of effect rivaling direct effects. Low pervasive confounder (lighter shades) means that a large proportion (5%) of variants have an effect on the confounder, but with low magnitude. All traits are simulated with high heritability (60%).* ***A)*** *The y-axis represents the precision which is the proportion of well-predicted variants among all predicted variants for a given label.* ***B)*** *The y-axis represents the recall, which is the proportion of well-predicted variants among all true variants for a given label. NB: In cases where no variant was clustered under a given label, it was impossible to calculate precision or recall; such instances are represented by squares below the x-axis.*

### Supplementary Results

#### PRISM vastly improves the interpretation of variant-trait associations detected in GWAS.

Genome-wide association studies (GWASs) aim to identify significant associations between genetic variants and a given trait, whereas PRISM re-examines these associations through the prism of other traits. We aimed to assess the added value of PRISM compared to traditional GWAS results. To do so, in our simulations, we calculated the precision and recall to detect direct effects using either genome-wide significant association from GWAS, or PRISM significant direct variants (Supplementary Fig. 24). First, in scenarios with low polygenicity, GWAS exhibited high recall but low precision to predict direct effects, as expected. Precision was particularly reduced in cases of strong pleiotropy, *i.e.* high confounder effects, and traits affected by vertical pleiotropy (*i.e.* traits $A$, $B_1$-$B_4$, $C_1$, $C_3$). In contrast, PRISM demonstrated high recall and high precision across all traits, regardless of the pleiotropic complexity. Second, in the most realistic scenarios with medium polygenicity, PRISM consistently outperformed GWAS in both precision and recall. Third, in scenarios with high polygenicity, since individual genetic variant effect sizes were small, GWAS exhibited high precision but low recall. GWAS only detected the strongest effect sizes, *i.e.* predominantly direct effects. On the other hand, PRISM largely outperformed GWAS in recall while maintaining comparable precision. We conclude that PRISM, using GWAS results, can vastly improve the biological interpretation of significant associations detected in GWAS.

#### PRISM labels can be influenced by the set of processed traits.

In our simulations, we applied PRISM to the simulated complex network of traits (See Fig. 6), but purposefully omitted one of the most pivotal traits of the first subnetwork ($B_4^1$, See Methods) from input data. To keep the total number of traits processed by PRISM constant, we included an additional trait $E_0$ with neither any causal interaction nor any confounder with any other trait. As shown in Supplementary Fig. 21, the precision for predicting direct effects remained largely comparable to that obtained in the complete simulation, with the exception of a specific case where the sole causal link to a trait, namely $B_4^1$ for $B_3^1$, was ommitted. In this setting, we observed reduced precision for predicting direct effects on $B_3^1$ across all scenarios, as the vertical pleiotropy from $B_4^1$ to $B_3^1$ was not included in the analysis. In this case, PRISM was unable to establish the mediation link with the omitted pivotal trait $B_4^1$. Thus, for the real data analysis, we selected a broad and representative set of complex traits and diseases from the UK Biobank (see Methods and Discussion) to mitigate this potential limitation.

#### PRISM is robust to reasonable parameter estimation errors.

PRISM relies on global parameters from LHC-MR to estimate the relationships between traits. Therefore, we tested the robustness of PRISM to inaccurate estimations of the global parameters in the prediction of variant-trait effects. Specifically, we replicated the analysis presented in Fig. 2, introducing random noise to the global parameters inputed in PRISM. Every parameter was sampled from a uniform distribution ranging from -20% to +20% of its true value. This process was repeated across 10 replicates, exclusively within the PRISM pipeline (effect sizes were not resimulated). We then calculated PRISM precision and recall in predicting variant-trait effects globally across replicates and compared these metrics to those obtained from the original parameters. The results, presented in Supplementary Fig 25, demonstrated that PRISM maintained robust precision and recall despite reasonable estimation errors in the global parameters, especially for medium and high polygenicity which are more realistic. It is worth mentioning that the apparent drop in confounder precision observed at low polygenicity only stemmed from the extremely small number of predicted confounder effects in these scenarios. In other words, predicting one wrong confounder effect could lead to a drop of 100% in precision in this particular case.

#### PRISM precision is robust to lower sample size

PRISM takes as input standardized effect sizes that are influenced by sample size, and sample size serves as a parameter within the PRISM model. Therefore, to assess PRISM robustness to reduced sample size, we replicated the simulations presented in Fig. 2 using a sample size of $N = 50{,}000$, compared to the original $N = 361{,}194$. This reduction in sample size increased the error term in the simulated effect sizes (see Supplementary Methods). The reduced sample size parameter was incorporated into PRISM, resulting in larger diagonal terms in matrix $\Sigma_0$, which integrates sample size into the model (see Supplementary Methods). As shown in Supplementary Fig. 26, PRISM demonstrated a minimal decrease in precision but exhibited a more pronounced decrease in recall, as expected. Importantly, in PRISM we deliberately favor precision over recall.

#### PRISM accurately predicts direct variant-trait effects in the absence of pleiotropy.

In our simulations, we applied PRISM to the network of 60 traits (See Fig. 6), but purposefully removed pleiotropy. Specifically, we set all causal effects between traits to zero (no vertical pleiotropy), set all confounder effects to zero (no confounder pleiotropy), and prevented variants to exert direct effects on more than one trait (no horizontal pleiotropy). As shown in Supplementary Fig. 27, PRISM exhibited excellent precision and high recall to detect direct effects in these scenarios. In the highly polygenic scenario, a recall of approximately 50% may appear underwhelming. However, given the fixed total heritability and high polygenicity, there were thousands of direct variants, many of which had extremely small effect sizes that PRISM could not reliably distinguish from non-causal variants. We sought to determine the smallest effect sizes that PRISM could reliably detect. For a fixed level of total heritability, the scenario with high polygenicity involved many variants with small effects, while low polygenicity involved fewer variants with larger effects. Analyzing the distribution of effect sizes, we found that the smallest detectable effects had median magnitudes of ~1.4, ~1.8, and ~1.7 standard deviations for high, medium, and low polygenicity respectively.

#### PRISM power to detect direct effects is mostly independent of the number of input traits.

A variant–trait association is considered PRISM-significant if the sign test yields a p-value below $\frac{5\times10^{-8}}{T-1}$, for $T$ traits included in the analysis. The sign test is expected to produce smaller p-values for true direct effects as more traits are included, as the probability of affecting $X$ should always be greater than the probability of a null effect. In simulations, in a scenario with medium polygenicity, low pervasive confounder, and strong causal effects (Scenario 26, Supplementary Table 1), we tested whether increasing the number of traits would enhance the recall to detect true direct effects.

Specifically, we ran PRISM on 15 (one subnetwork), 30 (two subnetworks), 45 (three subnetworks), and 60 (four subnetworks) traits. For 15 and 30 traits, the recall was zero, indicating that too few traits were included for the sign test to reach the significance threshold. However, when the number of traits increased to 45 or 60, the global direct recall across all traits remained essentially the same (~79%), suggesting that once the number of 31 traits is exceeded, adding more traits does not substantially increase power. Furthermore, precision was similar (~95%) under both 45 and 60 traits. We conclude that with the sign test, once the 31 traits bar is passed, additional traits do not affect the power to detect direct effects.

##### PRISM reliably detects horizontal pleiotropy in simulations.

In real data, horizontal pleiotropy was surprisingly rare, accounting for only 0.17% of the observed pleiotropy in GWAS. In simulations, we investigated whether this scarcity could result from a bias in PRISM against horizontal pleiotropy. A horizontal variant-trait effect is defined as a direct effect from a variant with at least one other direct effect. Given the low probability of randomly generating horizontal pleiotropy in scenarios with low polygenicity, we introduced four true horizontal variants (See Supplementary Methods for details). In our simulations, we assessed PRISM ability to predict all true horizontal variant-trait effects (Supplementary Fig. 28). Our findings indicate that PRISM is adequately capable of detecting horizontal pleiotropy and does not exhibit bias against it.

##### PRISM is mostly robust to multiple confounders.

Both PRISM and LHC-MR assume the presence of a single confounder influencing each pair of traits. The confounder is notably modeled using fixed priors for polygenicity and heritability, which are neither specified by the user nor derived from the input data. Therefore, we tested the robustness of PRISM under conditions involving multiple confounders with distinct characteristics. Specifically, we conducted simulations replicating the analysis from Fig. 2, but introduced two simultaneous, independent confounders for each trait pair: one confounder exhibiting high targeted effects (0.01% polygenicity), and another with pervasive but low-magnitude effects (5% polygenicity). The results, presented in Supplementary Fig. 29, demonstrated that PRISM maintained robust precision and recall for direct and vertical effects in the presence of two confounders. Precision in detecting confounder effects remains similar to high targeted confounder scenarios. However, the recall for confounder effects was very low, reflecting PRISM limited power to detect the numerous weak-effect variants arising from the pervasive confounders.

##### PRISM provides additional insights on trait relationships.

Mendelian Randomization, specifically LHC-MR in this study, predicts vertical effects among traits. We compared the LHC-MR estimates of these trait relationships with the combined vertical variant–trait effects identified by PRISM in the deconvoluted networks. In Supplementary Fig. 30, we observed that for most vertical relationships, PRISM was able to pinpoint significant vertical variant-trait effects corresponding to these trait relationships. Interestingly, certain causal relationships identified by LHC-MR did not correspond to vertical effects predicted by PRISM. For instance, LHC-MR detected causal associations between several anthropometric traits (BMI, body fat percentage, waist circumference, and standing height) and coronary heart disease (CHD), yet PRISM did not predict any corresponding vertical variants. We can hypothesize that these causal relationships identified by LHC-MR might be mediated by biomarkers, lipids, or lower-level traits rather than by anthropometric traits.

Supporting this hypothesis, PRISM notably predicted vertical variant effects from apolipoprotein B and diastolic blood pressure. Additionally, multiple traits, including anthropometrics, biomarkers, and lipids, were identified by LHC-MR as having causal effects on Type 2 diabetes. However, PRISM predicted no vertical variants for these traits, highlighting the complexity of the Type 2 diabetes pathogenesis.

#### PRISM direct variants show no distinctive features in large-scale genetic studies.

One of our key results is the enriched per-variant heritability observed in PRISM direct variants. Consequently, we investigated whether direct variants exhibit distinctive characteristics in other large-scale genetic studies. First, we compared CADD scores[1], that assess the deleteriousness of genetic variants, between direct, pleiotropic and GWAS variants. Second, we compared minor allele frequencies from GNOMAD[2] v4.0 between direct, pleiotropic and GWAS variants, across African, American, and European ancestries. Third, we compared predicted pathogenicity scores of variants from AlphaMissense[3] between direct, pleiotropic and GWAS variants. In all three analyses, we found no significant differences between direct, pleiotropic, and GWAS variants. We concluded that direct variants were typical genetic variants that are neither less common, nor significantly more pathogenic or deleterious. The distinction between direct and pleiotropic variants lies in what they affect rather than the nature of their impact.

#### PRISM produces coherent results on a panel of literature-validated variants.

We attempted to systematically compare PRISM results to a set of gold-standard variants from Open Targets[4]. However, establishing a systematic criterion for comparison was very challenging and the high redundancy in the traits with validated variants made it less compelling. Therefore, we chose to examine a panel of variants from Open Targets with a variety of traits. Here are some additional examples where PRISM inferred a variant network aligned with the literature.

Variant rs1697421 (Supplementary Fig. 31A) mapping to gene ALPL which encodes for Alkaline Phosphatase has been validated for its association with phosphorus levels. Additionally, ALPL has been studied as a potential biomarker for C-reactive protein (CRP)[5]. These findings highlighted the significance of ALPL in metabolic and inflammatory processes.

The genetic variant rs1471633 (Supplementary Fig. 31B) is located in the PDZK1 gene, which encodes a protein involved in the regulation of urate transport[6]. Additionally, variant rs1967017, which is in complete linkage disequilibrium with rs1471633, has been validated as impacting PDZK1 expression and altering the excretion of urate[7]. This is consistent with PRISM predictions, which indicated that rs1471633 directly affected urate levels, subsequently inducing a vertical effect on gout disease.

Variant rs560887 (Supplementary Fig. 31C) is located in the G6PC2 gene, coding for a protein involved in the gluconeogenic and glycogenolytic pathways[8]. Polymorphisms in G6PC2 have been validated in mice to alter fasting blood glucose levels, which are associated with type 2 diabetes (T2D)[9]. Interestingly, PRISM predicted a direct effect of this variant on HbA1c, a marker of blood glucose levels. It has been shown that variant rs560887 is linked to higher HbA1c in individuals with glucokinase mutations in MODY (Maturity Onset Diabetes of the Young) patients. Specifically, individuals who are GG homozygotes are more likely to meet the diagnostic criteria for diabetes based on their HbA1c levels[10]. Unfortunately, PRISM was not able to establish a vertical link between HbA1c and T2D, but did identify a confounder effect on glucose levels and T2D.

Variant rs6993770 (Supplementary Fig. 31D) maps to the ZFPM2 gene, which encodes a transcription factor of the FOG family, known for regulating hematopoiesis. This variant has been validated as causal for plateletcrit, which is the proportion of blood volume occupied by platelets. Although associated with multiple blood traits in GWAS, PRISM only found a sole direct effect on platelet count and only vertical effects on the additional blood traits.

Variant rs429358 (Supplementary Fig. 31E) make allele APOE-$\varepsilon 4$ of apolipoprotein E. The allele is well known for its involvement in cognitive impairment, specifically Alzheimer's disease. But the allele has also been linked to the metabolic syndrome[11,12] which is a group of conditions that occur simultaneously, increasing the risk of heart disease, stroke, and type 2 diabetes, and that includes high blood pressure, high blood sugar, excess body fat around the waist, and abnormal cholesterol or triglyceride levels in full agreement with PRISM findings.

Variant rs12970134 (Supplementary Fig. 31F) is located in the MC4R (Melanocortin 4 Receptor) gene, coding for the melanocortin 4 receptor protein, a key regulator of energy homeostasis. This variant has been significantly associated with body mass index (BMI) in a case-control study[13]. Interestingly, the MC4R gene is heavily studied due to its critical role in obesity[14–18]. Notably, a recent article[19] has highlighted that loss-of-function variants in the MC4R gene have substantial impacts on BMI, weight, fat mass, and lean mass. These findings align with PRISM, which identifies a direct effect of this variant on BMI and basal metabolic rate. Consequently, a myriad of pleiotropic ripple effects was predicted, for example vertical effects on body impedance, waist circumference, body fat percentage, HDL-cholesterol, and Apolipoprotein A. Pleiotropic effects on inflammation markers, urate, and hypertension further linked this variant to obesity-related complications.

Variant rs12150660 (Supplementary Fig. 31G) is located in the SHBG gene, coding for the Sex Hormone-Binding Globulin protein (SHBG). This protein binds and regulates androgens and estrogens, exhibiting a particularly high affinity for testosterone[20]. PRISM did predict that this variant had a direct impact on SHBG, and a vertical effect on testosterone levels through SHBG. Additionally, PRISM forecasted potential confounder effects on various traits, such as diastolic blood pressure, inflammation markers, and body impedance. A review of the scientific literature suggests that these confounder effects might be linked to metabolic syndrome, as testosterone deficiency is implicated in its development[21].

Variant rs738408 (Supplementary Fig. 31H) maps to the PNPLA3 gene, coding for a triacylglycerol lipase involved in the hydrolysis of triacylglycerol in adipocytes. This variant is associated with NAFLD[22] (non-alcoholic fatty liver disease), and is in tight LD with rs738409, which accounts for a large fraction of liver disease heritability[23]. However, the precise role of PNPLA3 in liver lipid metabolism remains unclear[24]. In any case, the link between these variants and liver diseases accounts for the numerous confounder effects on inflammation markers, body impedance, and even gout[25] predicted by PRISM. Furthermore, variant rs738409 is linked to variations in alanine aminotransferase levels[26], aligning with PRISM vertical effect, though further details are lacking.

Genetic variant rs7741021 (Supplementary Fig. 31I) is located in the RSPO3 gene, coding for the R-spondin-3 protein. This variant has been significantly associated with bone mass[27]. Additionally, rs7741021 has been shown to affect circulating levels of R-spondin-3 protein, where elevated levels are linked to increased bone mineral density (BMD) and a substantially reduced risk of distal forearm fractures[28]. These results aligned with the direct effect of this variant on BMD as predicted by PRISM. Furthermore, PRISM predicted pleiotropic effects on lipid traits, including ApoB, triglycerides, and HDL-C levels. This can be explained by the role of RSPO3 in influencing peripheral adipose tissue storage capacity[29], providing insight into these additional effects.

#### Randomly shuffling PRISM labels eliminates the direct variants heritability enrichment.

One of our key results is the enriched per-variant heritability observed in PRISM direct variants. To assess whether this enrichment could be explained solely by the smaller number of variants or other spurious factors, we randomly reassigned the pleiotropic labels while preserving the total number of variants in each category, and then re-ran s-LDSC. As shown in Supplementary Fig. 32, this shuffling completely eliminated the enrichment signal, indicating that the observed enrichment is a genuine signal rather than an artifact.

### Supplementary Tables

*Table 1: Parameters of all 32 scenarios, used to simulate GWAS summary statistics.*

| Scenario | h2_AB | h2_CDE | Trait Polygenicity | Confounder Polygenicity | Causal effect (except B4) | Causal effect (to B4) |
|---|---|---|---|---|---|---|
| 1 | 0.2 | 0.2 | 0.0001 | 0.05 | 0.3 | 0.3 |
| 2 | 0.2 | 0.6 | 0.0001 | 0.05 | 0.3 | 0.3 |
| 3 | 0.6 | 0.2 | 0.0001 | 0.05 | 0.3 | 0.3 |
| 4 | 0.6 | 0.6 | 0.0001 | 0.05 | 0.3 | 0.3 |
| 5 | 0.2 | 0.2 | 0.0001 | 0.0001 | 0.3 | 0.3 |
| 6 | 0.2 | 0.6 | 0.0001 | 0.0001 | 0.3 | 0.3 |
| 7 | 0.6 | 0.2 | 0.0001 | 0.0001 | 0.3 | 0.3 |
| 8 | 0.6 | 0.6 | 0.0001 | 0.0001 | 0.3 | 0.3 |
| 9 | 0.2 | 0.2 | 0.0001 | 0.05 | 0.3 | 0.05 |
| 10 | 0.2 | 0.6 | 0.0001 | 0.05 | 0.3 | 0.05 |
| 11 | 0.6 | 0.2 | 0.0001 | 0.05 | 0.3 | 0.05 |
| 12 | 0.6 | 0.6 | 0.0001 | 0.05 | 0.3 | 0.05 |
| 13 | 0.2 | 0.2 | 0.0001 | 0.0001 | 0.3 | 0.05 |
| 14 | 0.2 | 0.6 | 0.0001 | 0.0001 | 0.3 | 0.05 |
| 15 | 0.6 | 0.2 | 0.0001 | 0.0001 | 0.3 | 0.05 |
| 16 | 0.6 | 0.6 | 0.0001 | 0.0001 | 0.3 | 0.05 |
| 17 | 0.2 | 0.2 | 0.1 | 0.05 | 0.3 | 0.3 |
| 18 | 0.2 | 0.6 | 0.1 | 0.05 | 0.3 | 0.3 |
| 19 | 0.6 | 0.2 | 0.1 | 0.05 | 0.3 | 0.3 |
| 20 | 0.6 | 0.6 | 0.1 | 0.05 | 0.3 | 0.3 |
| 21 | 0.2 | 0.2 | 0.1 | 0.0001 | 0.3 | 0.3 |
| 22 | 0.2 | 0.6 | 0.1 | 0.0001 | 0.3 | 0.3 |
| 23 | 0.6 | 0.2 | 0.1 | 0.0001 | 0.3 | 0.3 |
| 24 | 0.6 | 0.6 | 0.1 | 0.0001 | 0.3 | 0.3 |
| 25 | 0.2 | 0.2 | 0.1 | 0.05 | 0.3 | 0.05 |
| 26 | 0.2 | 0.6 | 0.1 | 0.05 | 0.3 | 0.05 |

| Scenario | h2_AB | h2_CDE | Trait Polygenicity | Confounder Polygenicity | Causal effect (except B4) | Causal effect (to B4) |
|---|---|---|---|---|---|---|
| 27 | 0.6 | 0.2 | 0.1 | 0.05 | 0.3 | 0.05 |
| 28 | 0.6 | 0.6 | 0.1 | 0.05 | 0.3 | 0.05 |
| 29 | 0.2 | 0.2 | 0.1 | 0.0001 | 0.3 | 0.05 |
| 30 | 0.2 | 0.6 | 0.1 | 0.0001 | 0.3 | 0.05 |
| 31 | 0.6 | 0.2 | 0.1 | 0.0001 | 0.3 | 0.05 |
| 32 | 0.6 | 0.6 | 0.1 | 0.0001 | 0.3 | 0.05 |
| 33 | 0.2 | 0.2 | 0.01 | 0.05 | 0.3 | 0.3 |
| 34 | 0.2 | 0.6 | 0.01 | 0.05 | 0.3 | 0.3 |
| 35 | 0.6 | 0.2 | 0.01 | 0.05 | 0.3 | 0.3 |
| 36 | 0.6 | 0.6 | 0.01 | 0.05 | 0.3 | 0.3 |
| 37 | 0.2 | 0.2 | 0.01 | 0.0001 | 0.3 | 0.3 |
| 38 | 0.2 | 0.6 | 0.01 | 0.0001 | 0.3 | 0.3 |
| 39 | 0.6 | 0.2 | 0.01 | 0.0001 | 0.3 | 0.3 |
| 40 | 0.6 | 0.6 | 0.01 | 0.0001 | 0.3 | 0.3 |
| 41 | 0.2 | 0.2 | 0.01 | 0.05 | 0.3 | 0.05 |
| 42 | 0.2 | 0.6 | 0.01 | 0.05 | 0.3 | 0.05 |
| 43 | 0.6 | 0.2 | 0.01 | 0.05 | 0.3 | 0.05 |
| 44 | 0.6 | 0.6 | 0.01 | 0.05 | 0.3 | 0.05 |
| 45 | 0.2 | 0.2 | 0.01 | 0.0001 | 0.3 | 0.05 |
| 46 | 0.2 | 0.6 | 0.01 | 0.0001 | 0.3 | 0.05 |
| 47 | 0.6 | 0.2 | 0.01 | 0.0001 | 0.3 | 0.05 |
| 48 | 0.6 | 0.6 | 0.01 | 0.0001 | 0.3 | 0.05 |

*Table 2: Description of the 70 traits included from the UK Biobank. The first column specifies the trait category, the following three columns correspond to the Trait code in UK Biobank, the description of the trait in UK Biobank and the trait name we used throughout the paper. The final colunm specifies the sample size.*

| Category | Trait Code in UK Biobank | Description of the trait in UK Biobank | Trait name | Sample size |
|---|---|---|---|---|
| Anthropometric trait | 3148_irnt | Heel bone mineral density (BMD) | BMD | 206,496 |

| Category | Trait Code in UK Biobank | Description of the trait in UK Biobank | Trait name | Sample size |
|---|---|---|---|---|
| Anthropometric trait | 23099_irnt | Body fat percentage | Body fat percentage | 354,628 |
| Anthropometric trait | 23106_irnt | Impedance of whole body | Body impedance | 354,795 |
| Anthropometric trait | 21001_irnt | Body mass index (BMI) | BMI | 359,983 |
| Anthropometric trait | 50_irnt | Standing height | Standing height | 360,388 |
| Anthropometric trait | 48_irnt | Waist circumference | Waist circumference | 360,564 |
| Biomarker | 30850_irnt | Testosterone (nmol/L) | Testosterone | 312,102 |
| Biomarker | 30830_irnt | SHBG (nmol/L) | SHBG | 312,215 |
| Biomarker | 30810_irnt | Phosphate (mmol/L) | Phosphate | 314,658 |
| Biomarker | 30740_irnt | Glucose (mmol/L) | Glucose | 314,916 |
| Biomarker | 30860_irnt | Total protein (g/L) | Total protein | 314,921 |
| Biomarker | 30680_irnt | Calcium (mmol/L) | Calcium | 315,153 |
| Biomarker | 30600_irnt | Albumin (g/L) | Albumin | 315,268 |
| Biomarker | 30890_irnt | Vitamin D (nmol/L) | Vitamin D | 329,247 |
| Biomarker | 30770_irnt | IGF-1 (nmol/L) | IGF-1 | 342,439 |
| Biomarker | 30650_irnt | Aspartate aminotransferase (U/L) | Aspartate aminotransferase | 342,990 |
| Biomarker | 30710_irnt | C-reactive protein (mg/L) | CRP | 343,524 |
| Biomarker | 30880_irnt | Urate (umol/L) | Urate | 343,836 |
| Biomarker | 30670_irnt | Urea (mmol/L) | Urea | 344,052 |

| Category | Trait Code in UK Biobank | Description of the trait in UK Biobank | Trait name | Sample size |
|---|---|---|---|---|
| Biomarker | 30700_irnt | Creatinine (umol/L) | Creatinine | 344,104 |
| Biomarker | 30730_irnt | Gamma glutamyltransferase (U/L) | Gamma glutamyltransferase | 344,104 |
| Biomarker | 30620_irnt | Alanine aminotransferase (U/L) | Alanine aminotransferase | 344,136 |
| Biomarker | 30750_irnt | Glycated haemoglobin (mmol/mol) | HbA1C | 344,182 |
| Biomarker | 30720_irnt | Cystatin C (mg/L) | Cystatin C | 344,264 |
| Biomarker | 30610_irnt | Alkaline phosphatase (U/L) | Alkaline phosphatase | 344,292 |
| Blood trait | 30250_irnt | Reticulocyte count | Reticulocyte count | 344,729 |
| Blood trait | 30150 | Eosinophill count | Eosinophil count | 349,856 |
| Blood trait | 30160 | Basophill count | Basophil count | 349,856 |
| Blood trait | 30120_irnt | Lymphocyte count | Lymphocyte count | 349,856 |
| Blood trait | 30130_irnt | Monocyte count | Monocyte count | 349,856 |
| Blood trait | 30140_irnt | Neutrophill count | Neutrophil count | 349,856 |
| Blood trait | 30180_irnt | Lymphocyte percentage | Lymphocyte percentage | 349,861 |
| Blood trait | 30000_irnt | White blood cell (leukocyte) count | White blood cell count | 350,470 |
| Blood trait | 30070_irnt | Red blood cell (erythrocyte) distribution width | Red blood cell DW | 350,473 |
| Blood trait | 30080_irnt | Platelet count | Platelet count | 350,474 |
| Blood trait | 30010_irnt | Red blood cell (erythrocyte) count | Red blood cell count | 350,475 |

| Category | Trait Code in UK Biobank | Description of the trait in UK Biobank | Trait name | Sample size |
|---|---|---|---|---|
| Cancer trait | C_PANCREAS | Malignant neoplasm of pancreas | Pancreas cancer | 361,194 |
| Cancer trait | C3_SKIN | Malignant neoplasm of skin | Skin cancer | 361,194 |
| Cancer trait | C50 | Diagnoses - main ICD10: C50 Malignant neoplasm of breast | Breast cancer | 361,194 |
| Cancer trait | II_NEOPLASM | Neoplasms | Cancer | 361,194 |
| Education | 6138_1 | Qualifications: College or University degree | University degree | 357,549 |
| Education | 20023_irnt | Mean time to correctly identify matches | Match identification exercise | 358,695 |
| Health | 1757 | Facial ageing | Facial ageing | 330,409 |
| Health | 2178 | Overall health rating | Overall health rating | 359,681 |
| Inflammatory trait | K51 | Diagnoses - main ICD10: K51 Ulcerative colitis | UC | 361,194 |
| Lifestyle trait | 1468_4 | Cereal type: Muesli | Muesli intake | 299,898 |
| Lifestyle trait | 1528 | Water intake | Water intake | 333,363 |
| Lifestyle trait | 884 | Number of days/week of moderate physical activity 10+ minutes | Physical activity | 343,943 |
| Lifestyle trait | 1458 | Cereal intake | Cereal intake | 345,019 |
| Lifestyle trait | 1488_irnt | Tea intake | Tea intake | 349,376 |
| Lifestyle trait | 6155_100 | Vitamin and mineral | No vitamins | 359,245 |

| Category | Trait Code in UK Biobank | Description of the trait in UK Biobank | Trait name | Sample size |
|---|---|---|---|---|
| | | supplements: None of the above | | |
| Lifestyle trait | 20116_0 | Smoking status: Never | Never smoker | 359,706 |
| Lifestyle trait | 1558 | Alcohol intake frequency. | Alcohol freq | 360,726 |
| Lifestyle trait | 1200 | Sleeplessness / insomnia | Insomnia | 360,738 |
| Lipid | 30630_irnt | Apoliprotein A (g/L) | Apoliprotein A | 313,387 |
| Lipid | 30760_irnt | HDL cholesterol (mmol/L) | HDL-C | 315,133 |
| Lipid | 30640_irnt | Apoliprotein B (g/L) | Apoliprotein B | 342,590 |
| Lipid | 30780_irnt | LDL direct (mmol/L) | LDL-C | 343,621 |
| Lipid | 30870_irnt | Triglycerides (mmol/L) | Triglycerides | 343,992 |
| Lipid | 30690_irnt | Cholesterol (mmol/L) | Cholesterol | 344,278 |
| Metabolic trait | 4079_irnt | Diastolic blood pressure, automated reading | DBP | 340,162 |
| Metabolic trait | 23105_irnt | Basal metabolic rate | Basal metabolic rate | 354,825 |
| Metabolic trait | 20002_1065 | Non-cancer illness code, self-reported: hypertension | Hypertension | 361,141 |
| Metabolic trait | 20002_1223 | Non-cancer illness code, self-reported: type 2 diabetes | T2D | 361,141 |
| Metabolic trait | 20002_1466 | Non-cancer illness code, self-reported: gout | Gout | 361,141 |
| Metabolic trait | I25 | Diagnoses - main ICD10: I25 Chronic ischaemic heart disease | CHD | 361,194 |

| Category | Trait Code in UK Biobank | Description of the trait in UK Biobank | Trait name | Sample size |
|---|---|---|---|---|
| Psychiatric trait | 20127_irnt | Neuroticism score | Neuroticism score | 293,006 |
| Psychiatric trait | V_MENTAL_BEHAV | Mental and behavioural disorders | Mental disorders | 361,194 |
| Respiratory trait | 3062_irnt | Forced vital capacity (FVC) | Forced vital capacity | 329,404 |
| Respiratory trait | 20002_1111 | Non-cancer illness code, self-reported: asthma | Asthma | 361,141 |

### Supplementary Methods

#### LHC-MR

The PRISM method relies on the Mendelian Randomization (MR) model from LHC-MR[30,31]. LHC-MR is an integrative MR method that aims to infer the causal relationship of a pair of traits from GWAS summary statistics, while taking into account a latent confounder. The principle of PRISM is to reroute the trait-level MR model from LHC-MR to infer information at the level of genetic variants. Thus, to introduce our PRISM model, we recapitulate some elements of the model from LHC-MR, specifically part 1, 2 and 3.

**Part 1: Basic structural Equation Model**

The structural equation model is defined by the following equations:

$$X = q_x \cdot U + \alpha_{y\to x}Y + \mathbf{G} \cdot \vec{\gamma}_x + \epsilon_X \qquad (A)$$

$$Y = q_y \cdot U + \alpha_{x\to y}Y + \mathbf{G} \cdot \vec{\gamma}_y + \epsilon_Y \qquad (B)$$

$$U = \mathbf{G} \cdot \vec{\gamma}_u + \epsilon_U \qquad (C)$$

In this model (See Supplementary Fig. 1):

- $X$ and $Y$ are continuous random variables representing two complex traits.
- $U$ is a continuous random variable representing a latent heritable confounder, with causal effects $q_x$ and $q_y$ on $X$ and $Y$, respectively.
- To simplify the notations, we assume that $E(X) = E(Y) = E(U) = 0$ and $Var(X) = Var(Y) = Var(U) = 1$.
- $X$ and $Y$ have causal effects on each other, denoted $\alpha_{x\to y}$ and $\alpha_{y\to x}$.
- The genome-wide genotype data for $m$ genetic variants and $n$ individuals are denoted by $\mathbf{G} \in \mathbb{R}^{n\times m}$. The vectors $\vec{\gamma}_x, \vec{\gamma}_y, \vec{\gamma}_u \in \mathbb{R}^m$ represent the true multivariate direct effects of all $m$ genetic variants on $X$, $Y$ and $U$, respectively.
- $\epsilon_X \sim \mathcal{N}(0, \sigma_x^2)$, $\epsilon_Y \sim \mathcal{N}(0, \sigma_y^2)$, and $\epsilon_U \sim \mathcal{N}(0, \sigma_u^2)$ are normally distributed and mutually independent error terms.

We assume that only a proportion $0 \le \lambda_x, \lambda_y, \lambda_u \le 1$ of the genome has a direct effect on $X$, $Y$ and $U$, respectively. The vectors $\vec{\gamma}_x, \vec{\gamma}_y, \vec{\gamma}_u$ are modeled using a spike-and-slab distribution:

$$\vec{\gamma}_a = \vec{\xi}_a \odot \vec{\kappa}_a \qquad (D)$$

where $\xi_a \sim \mathcal{B}(m, \lambda_a)$ and $\kappa_a \sim \mathcal{N}(0, \frac{h_a^2}{m\lambda_a})$ for $a = x, y, u$.

$\vec{\xi}_a, \vec{\kappa}_a \in \mathbb{R}^m$ are vectors of dimension $m$. Let $A$ denote either trait $X$, $Y$, or $U$. The vector $\vec{\xi}_a$ consists of binary elements (0 or 1) indicating whether the genetic variant has an effect on $A$. The effect $\vec{\kappa}_a$ follows a Gaussian distribution. $h_x^2, h_y^2, h_u^2$ represent the heritability of $X$, $Y$ et $U$, respectively. The symbol $\odot$ denotes the element-wise product. We assume zero covariance between the direct effects of a genetic variant on $X$, $Y$ and $U$, thus $\mathbf{cov}(\vec{\gamma}_x, \vec{\gamma}_y) = \mathbf{cov}(\vec{\gamma}_x, \vec{\gamma}_y) = \mathbf{cov}(\vec{\gamma}_x, \vec{\gamma}_y) = 0$.

**Part 2: Reparametrization of the summary statistics distributions.**

We observe the univariate association summary statistics on traits $X$ and $Y$ from finite samples $N_x$ and $N_y$, of size $n_x$ and $n_y$ respectively. The realizations of $X$, $Y$ and $U$ are denoted by $\vec{x}, \vec{y}, \vec{u} \in \mathbb{R}^{n_x}$. The genome-wide genetic data for $n_x$ individuals with $m$ variants are represented by $\mathbf{G}_x \in \mathbb{R}^{n_x \times m}$. The genetic data for a variant $k$ which is tested for association across $n_x$ individuals are denoted $\vec{g}_k \in \mathbb{R}^{n_x}$. All variant genotypes are standardized to a standard normal distribution.

$$\mathbf{G} = \begin{pmatrix} g_1^1 & g_2^1 & \cdots & g_k^1 & \cdots & g_m^1 \\ \vdots & \vdots & & \vdots & & \vdots \\ g_1^i & g_2^i & \cdots & g_k^i & \cdots & g_m^i \\ \vdots & \vdots & & \vdots & & \vdots \\ g_1^{n_x} & g_2^{n_x} & \cdots & g_k^{n_x} & \cdots & g_m^{n_x} \end{pmatrix} \in \mathbb{R}^{n_x \times m}$$

The association summary statistics $\beta_k^x$ for variant $k$ on trait $X$ can be written:

$$\beta_k^x = \frac{\langle \vec{g}_k, \vec{x} \rangle}{n_x}$$

Substituting $\vec{x}$ with its expression from equation $(A)$, we obtain:

$$\beta_k^x = \frac{q_x \cdot \langle \vec{g}_k, \vec{u} \rangle + \alpha_{y \to x} \cdot \langle \vec{g}_k, \vec{y} \rangle + \langle \vec{g}_k, \mathbf{G}_x \cdot \vec{\gamma}_x \rangle + g_k \cdot \epsilon_x}{n_x}$$

Further substituting $\vec{y}$ and $\vec{u}$ with their expression from equations $(B)$ and $(C)$, we obtain:

$$\beta_k^x = \frac{q_x \cdot \langle \vec{g}_k, \mathbf{G}_x \cdot \vec{\gamma}_u + \epsilon_u \rangle}{n_x} + \frac{\alpha_{y \to x} \cdot \langle \vec{g}_k, (q_y \cdot \mathbf{G}_x \cdot \vec{\gamma}_u + \epsilon_u) + \alpha_{x \to y} \vec{x} + (\mathbf{G}_x \cdot \vec{\gamma}_y + \epsilon_y) \rangle}{n_x}$$
$$+ \frac{\langle \vec{g}_k, \mathbf{G}_x \cdot \vec{\gamma}_x \rangle + g_k \cdot \epsilon_x}{n_x}$$

$$\beta_k^x = \frac{q_x \cdot \langle \vec{g}_k, \mathbf{G}_x \cdot \vec{\gamma}_u \rangle}{n_x} + q_x \cdot \epsilon_k^u + \frac{\alpha_{y \to x} q_y \cdot \langle \vec{g}_k, \mathbf{G}_x \cdot \vec{\gamma}_u \rangle}{n_x} + \alpha_{y \to x} q_y \cdot \epsilon_k^u + \alpha_{y \to x} \alpha_{x \to y} \beta_k^x$$
$$+ \frac{\alpha_{y \to x} \langle \vec{g}_k, \mathbf{G}_x \cdot \vec{\gamma}_y \rangle}{n_x} + \alpha_{y \to x} \epsilon_k^y + \frac{\langle \vec{g}_k, \mathbf{G}_x \cdot \vec{\gamma}_x \rangle}{n_x} + \epsilon_k^x$$

because $\beta_k^x = \frac{\langle \vec{g}_k, \vec{x} \rangle}{n_x}$ and with: $\epsilon_k^x = \frac{\langle \vec{g}_k \cdot \epsilon_x \rangle}{n_x} \sim \mathcal{N}(0, \frac{\sigma_x^2}{n_x})$, $\epsilon_k^u = \frac{\langle \vec{g}_k \cdot \epsilon_u \rangle}{n_x} \sim \mathcal{N}(0, \frac{\sigma_u^2}{n_x})$, $\epsilon_k^y = \frac{\langle \vec{g}_k \cdot \epsilon_y \rangle}{n_x} \sim \mathcal{N}(0, \frac{\sigma_y^2}{n_x})$

To simplify, we denote $\vec{\rho}_k = \frac{\mathbf{G}_x^\top \cdot \vec{g}_k}{n_x}$, as the Linkage Disequilibrium (LD) coefficients between variant $k$ and all markers in the genome. So:

$$\beta_k^x = q_x \cdot \langle \vec{\rho}_k, \vec{\gamma}_u \rangle + q_x \cdot \epsilon_k^u + \alpha_{y \to x} q_y \cdot \langle \vec{\rho}_k, \vec{\gamma}_u \rangle + \alpha_{y \to x} q_y \cdot \epsilon_k^u + \alpha_{y \to x} \alpha_{x \to y} \beta_k^x$$
$$+ \alpha_{y \to x} \langle \vec{\rho}_k, \vec{\gamma}_y \rangle + \alpha_{y \to x} \epsilon_k^y + \langle \vec{\rho}_k, \vec{\gamma}_x \rangle + \epsilon_k^x$$

We denote $\eta_k^x = (q_x + \alpha_{y \to x} \cdot q_y) \cdot \epsilon_u^k + \alpha_{y \to x} \epsilon_y^k + \epsilon_x^k \sim \mathcal{N}(0, \frac{i_x}{n_x})$, with $i_x = (q_x + \alpha_{y \to x} \cdot q_y)^2 \sigma_u^2 + \alpha_{y \to x} \sigma_y^2 + \sigma_x^2$. $i_x$ is equivalent to the LD score regression intercept[32]. So:

$$\beta_k^x = (\alpha_{y\to x} q_y + q_x) \cdot \langle \vec{\rho}_k, \vec{\gamma}_u \rangle + \alpha_{y\to x} \langle \vec{\rho}_k, \vec{\gamma}_y \rangle + \langle \vec{\rho}_k, \vec{\gamma}_x \rangle + \alpha_{y\to x} \alpha_{x\to y} \beta_k^x + \eta_k^x$$

We hypothesize that $\alpha_{y\to x}\alpha_{x\to y} \approx 0$ because in real traits, either one of the two causal effects is zero, or the product of the bidirectional causal effects is very small. In addition, as mentioned in the LHC-MR paper, in case of high bidirectional effects, the interpretation of all parameters becomes delicate. So, regrouping all $\beta_k^x$ on the left and factorizing:

$$\beta_k^x \approx (1 - \alpha_{y\to x}\alpha_{x\to y})\beta_k^x = (\alpha_{y\to x} q_y + q_x) \cdot \langle \vec{\rho}_k, \vec{\gamma}_u \rangle + \alpha_{y\to x} \langle \vec{\rho}_k, \vec{\gamma}_y \rangle + \langle \vec{\rho}_k, \vec{\gamma}_x \rangle + \eta_k^x$$

Then, we substitute equation $(D)$:

$$\beta_k^x = (\alpha_{y\to x} q_y + q_x) \cdot \langle \vec{\rho}_k, \vec{\xi}_u \odot \vec{\kappa}_u \rangle + \alpha_{y\to x} \langle \vec{\rho}_k, \vec{\xi}_x \odot \vec{\kappa}_x \rangle + \langle \vec{\rho}_k, \vec{\xi}_y \odot \vec{\kappa}_y \rangle + \eta_k^x$$

$$\beta_k^x = (\alpha_{y\to x} q_y + q_x) \cdot \langle \vec{\rho}_k \odot \vec{\xi}_u, \vec{\kappa}_u \rangle + \alpha_{y\to x} \langle \vec{\rho}_k \odot \vec{\xi}_y, \vec{\kappa}_y \rangle + \langle \vec{\rho}_k \odot \vec{\xi}_x, \vec{\kappa}_x \rangle + \eta_k^x$$

Assuming similar LD structures ($\vec{\rho}_k$), we apply the same reasoning for $\beta_k^y$:

$$\beta_k^y = (\alpha_{x\to y} q_x + q_y) \cdot \langle \vec{\rho}_k \odot \vec{\xi}_u, \vec{\kappa}_u \rangle + \alpha_{x\to y} \langle \vec{\rho}_k \odot \vec{\xi}_x, \vec{\kappa}_x \rangle + \langle \vec{\rho}_k \odot \vec{\xi}_y, \vec{\kappa}_y \rangle + \eta_k^y$$

with $\eta_k^x \sim \mathcal{N}(0, \frac{i_y}{n_y})$ and $i_y = (q_y + \alpha_{x\to y} \cdot q_x)^2 \sigma_u^2 + \alpha_{x\to y}\sigma_x^2 + \sigma_y^2$

Therefore, the joint distribution of can be written as:

$$\begin{pmatrix} \beta_k^x \\ \beta_k^y \end{pmatrix} = \begin{pmatrix} 1 \\ \alpha_{x\to y} \end{pmatrix} \cdot \langle \vec{\rho}_k \odot \vec{\xi}_x, \vec{\kappa}_x \rangle + \begin{pmatrix} \alpha_{y\to x} \\ 1 \end{pmatrix} \cdot \langle \vec{\rho}_k \odot \vec{\xi}_y, \vec{\kappa}_y \rangle + \begin{pmatrix} \alpha_{y\to x} \cdot t_y + t_x \\ \alpha_{x\to y} \cdot t_x + t_y \end{pmatrix} \cdot \langle \vec{\rho}_k \odot \vec{\xi}_u, \vec{\kappa}_u \rangle + \begin{pmatrix} \eta_x^k \\ \eta_y^k \end{pmatrix}$$

Following the same reasoning as the cross-trait LD-score regression[32], the noise term distribution is[30]:

$$\begin{pmatrix} \eta_x^k \\ \eta_y^k \end{pmatrix} \sim \mathcal{N}\left( \begin{pmatrix} 0 \\ 0 \end{pmatrix}, \begin{pmatrix} i_x/n_x & \dfrac{corr(X,Y) \cdot n_{x\cap y}}{n_x \cdot n_y} \\ \dfrac{corr(X,Y) \cdot n_{x\cap y}}{n_x \cdot n_y} & i_y/n_y \end{pmatrix} \right)$$

Here, $n_{x\cap y}$ represents the sample overlap between the two studies used to calculate $\hat{\beta}_k^x$ and $\hat{\beta}_k^y$, and $corr(X,Y)$ is the phenotypic correlation between variables $X$ and $Y$. $\frac{corr(X,Y)\cdot n_{x\cap y}}{\sqrt{n_x \cdot n_y}}$ is akin to the cross-trait LD Score regression intercept[33]. We set $corr(X,Y) = 0$. Our objective with PRISM is to identify pleiotropic variant-trait effects. We consider a variant-trait effect induced by the correlation between two traits not as noise, but as vertical pleiotropy. Furthermore, in the case of large sample size, the bias caused by sample overlap is expected to be small[34].

**Part 3: Derivation of the likelihood function.**

Let $\pi_x, \pi_u, \pi_y$ denote the fractions of variants for which $\vec{\rho}_k \odot \vec{\xi}_x$, $\vec{\rho}_k \odot \vec{\xi}_u$, $\vec{\rho}_k \odot \vec{\xi}_y$ are non-zero respectively. These are the fractions of observed variants that are in non-zero LD with at least one causal variant for $X$, $U$, $Y$ respectively. We assume that causal markers are sufficiently tagged by GWAS variants. Then $\pi_x > \lambda_x, \pi_u > \lambda_u, \pi_y > \lambda_y$, since LD propagates association signals beyond strictly causal markers. Next, we calculate the variance of the building blocks. For $a = x, u, y$:

$$Var(\langle \vec{\rho}_k \odot \vec{\xi}_a, \vec{\kappa}_a \rangle) = E[\langle \vec{\rho}_k \odot \vec{\xi}_a, \vec{\kappa}_a \rangle^2] - E[\langle \vec{\rho}_k \odot \vec{\xi}_a, \vec{\kappa}_a \rangle]^2$$

Given that $\rho_k, \xi_a, \kappa_a$ are independent:

$$Var(\langle \vec{\rho}_k \odot \vec{\xi}_a, \vec{\kappa}_a \rangle) = E[(\vec{\rho}_k \odot \vec{\xi}_a)^2] \cdot E[\vec{\kappa}_a^2] - E[\vec{\rho}_k \odot \vec{\xi}_a]^2 \cdot E[\vec{\kappa}_a]^2$$

Since $E[\vec{\kappa}_a] = 0$:

$$Var(\langle \vec{\rho}_k \odot \vec{\xi}_a, \vec{\kappa}_a \rangle) = E[(\vec{\rho}_k \odot \vec{\xi}_a)^2] \cdot E[\vec{\kappa}_a^2] = E[\vec{\rho}_k^2] \cdot E[\vec{\xi}_a^2] \cdot E[\vec{\kappa}_a^2] = l_k \cdot \lambda_a \cdot \frac{h_x^2}{\lambda_a m}$$

$$= l_k \cdot \frac{h_x^2}{m}$$

where $l_k = \langle \vec{\rho}_k, \vec{\rho}_k \rangle$ corresponds to the LD score.

Since $\langle \vec{\rho}_k \odot \vec{\xi}_a, \vec{\kappa}_a \rangle$ is modeled as a spike-and-slab Gaussian mixture model, with mixing proportion $\pi_a$, the non-zero component variance is the total variance divided by the mixing proportion. Thus, the distribution of the component can be expressed as:

$$\langle \vec{\rho}_k \odot \vec{\xi}_a, \vec{\kappa}_a \rangle \sim \pi_a \cdot \mathcal{N}(0, l_k \cdot \frac{h_a^2}{m \cdot \pi_a}) + (1 - \pi_a) \cdot \mathcal{N}(0,0)$$

Here, $\pi_a$ represents the proportion of genetic variants with a true causal effect on $A$ or in non-zero LD with such variants, and $(1 - \pi_a)$ represents the variants with no effect and in zero LD with causal variants.

- Next, we partition the genomic variants into eight disjoint components based on their associations with $X$, $Y$ and $U$:
  - (0) No association
  - (1) Associated with $X$
  - (2) Associated with $U$
  - (3) Associated with $Y$
  - (4) Associated with $X$ and $U$
  - (5) Associated with $X$ and $Y$
  - (6) Associated with $U$ and $Y$
  - (7) Associated with $X$, $Y$ and $U$
- We assume that the components are independent, so the proportion of variants in each component can be expressed as:
  - (0) $\omega_0 = (1 - \pi_u)(1 - \pi_x)(1 - \pi_y)$
  - (1) $\omega_1 = \pi_x\ (1 - \pi_u)(1 - \pi_y)$
  - (2) $\omega_2 = \pi_u\ (1 - \pi_x)(1 - \pi_y)$
  - (3) $\omega_3 = \pi_y\ (1 - \pi_u)(1 - \pi_x)$
  - (4) $\omega_4 = \pi_x\ \pi_u\ (1 - \pi_y)$

(5) $\omega_5 = \pi_x\, \pi_y\, (1 - \pi_u)$

(6) $\omega_6 = \pi_y\, \pi_u\, (1 - \pi_x)$

(7) $\omega_7 = \pi_x\, \pi_y\, \pi_u$

The joint distribution of $\begin{pmatrix}\beta_k^x \\ \beta_k^y\end{pmatrix}$ depends on the component from which variant $k$ is drawn. For example, if variant $k$ is drawn from component (1):

$$\begin{pmatrix}\beta_k^x \\ \beta_k^y\end{pmatrix} = \begin{pmatrix}1 \\ \alpha_{x\to y}\end{pmatrix} \cdot \langle \vec{\rho}_k \odot \vec{\xi}_x, \vec{\kappa}_x \rangle + \begin{pmatrix}\eta_x^k \\ \eta_y^k\end{pmatrix}$$

because $\vec{\xi}_y = \vec{0}$ and $\vec{\xi}_u = \vec{0}$.

$$\begin{pmatrix}\beta_k^x \\ \beta_k^y\end{pmatrix} \sim \begin{pmatrix}1 \\ \alpha_{x\to y}\end{pmatrix} \cdot \pi_x \cdot \mathcal{N}(0, l_k \cdot \frac{h_x^2}{m \cdot \pi_x}) + \begin{pmatrix}\eta_x^k \\ \eta_y^k\end{pmatrix}$$

$$= \mathcal{N}\left(\begin{pmatrix}0 \\ 0\end{pmatrix}, l_k \cdot \frac{h_x^2}{m \cdot \pi_x}\begin{bmatrix}1 & \alpha_{x\to y} \\ \alpha_{x\to y} & {\alpha_{x\to y}}^2\end{bmatrix}\right) + \begin{pmatrix}\eta_x^k \\ \eta_y^k\end{pmatrix}$$

Following the same reasoning for all 8 components, and summing them according to their respective proportion $\omega_j$:

$$\begin{pmatrix}\beta_k^x \\ \beta_k^y\end{pmatrix} \sim \sum_{i=0}^{7} \omega_j \cdot \mathcal{N}\left(\begin{pmatrix}0 \\ 0\end{pmatrix}, \mathbf{\Omega}_{ki}\right)$$

- where:
  - (0) $\mathbf{\Omega}_{k0} = \mathbf{\Sigma}_0$
  - (1) $\mathbf{\Omega}_{k1} = l_k \cdot \mathbf{\Sigma}_x + \mathbf{\Omega}_0$
  - (2) $\mathbf{\Omega}_{k2} = l_k \cdot \mathbf{\Sigma}_u + \mathbf{\Omega}_0$
  - (3) $\mathbf{\Omega}_{k3} = l_k \cdot \mathbf{\Sigma}_y + \mathbf{\Omega}_0$
  - (4) $\mathbf{\Omega}_{k4} = l_k \cdot (\mathbf{\Sigma}_x + \mathbf{\Sigma}_u) + \mathbf{\Omega}_0$
  - (5) $\mathbf{\Omega}_{k5} = l_k \cdot (\mathbf{\Sigma}_x + \mathbf{\Sigma}_y) + \mathbf{\Omega}_0$
  - (6) $\mathbf{\Omega}_{k6} = l_k \cdot (\mathbf{\Sigma}_u + \mathbf{\Sigma}_y) + \mathbf{\Omega}_0$
  - (7) $\mathbf{\Omega}_{k7} = l_k \cdot (\mathbf{\Sigma}_x + \mathbf{\Sigma}_u + \mathbf{\Sigma}_y) + \mathbf{\Omega}_0$

and:

$$\mathbf{\Sigma}_u = \frac{h_u^2}{m \cdot \pi_u}\begin{bmatrix}(\alpha_{y\to x} \cdot q_y + q_x)^2 & (\alpha_{y\to x} \cdot q_y + q_x) \cdot (\alpha_{x\to y} \cdot q_x + q_y) \\ (\alpha_{y\to x} \cdot q_y + q_x) \cdot (\alpha_{x\to y} \cdot q_x + q_y) & (\alpha_{x\to y} \cdot q_x + q_y)^2\end{bmatrix}$$

$$\mathbf{\Sigma}_x = \frac{h_x^2}{m \cdot \pi_x}\begin{bmatrix}1 & \alpha_{x\to y} \\ \alpha_{x\to y} & {\alpha_{x\to y}}^2\end{bmatrix}$$

$$\mathbf{\Sigma}_y = \frac{h_y^2}{m \cdot \pi_y}\begin{bmatrix}{\alpha_{y\to x}}^2 & \alpha_{y\to x} \\ \alpha_{y\to x} & 1\end{bmatrix}$$

$$\Sigma_0 = \begin{bmatrix} \frac{i_x}{n_x} & \rho_{x\cap y} \\ \rho_{x\cap y} & \frac{i_y}{n_y} \end{bmatrix}$$

Since the sum of two independent normally distributed random variables is Gaussian, when calculating the distribution of $\begin{pmatrix} \beta_k^x \\ \beta_k^y \end{pmatrix}$ for each component, the terms $\langle \vec{\rho}_k \odot \vec{\xi}_a, \vec{\kappa}_a \rangle$ are either $\vec{0}$ or independent Gaussian distributions (as affecting $X$, $Y$ or $U$ is independent). Therefore, the sum of $\langle \vec{\rho}_k \odot \vec{\xi}_a, \vec{\kappa}_a \rangle$ terms is a Gaussian distribution, making each component modeled with a Gaussian distribution. It is worth mentioning that, although each component is modeled with a Gaussian distribution, the global model is not Gaussian. This is not an issue since likelihoods and posterior probabilities are calculated independently for each component.

We chose to use a model with 8 nested components, whereas it would be possible to use a simpler model with just 4 components ($O$, $X$, $Y$, and $U$). However, $U$ is a latent variable and $\pi_u$ is an unknown prior. We do not want to risk masking effects on $X$ and $Y$ because of $U$. For example, let us consider a genetic variant $v$ with causal effects on $X$ and $U$. With a 4-components model, $v$ would have a high probability to belong to component $X$ and to belong to component $U$. However, we would have no way to interpret these probabilities. We would not be able to compare probabilities to conclude if $v$ affects $X$, $U$, or both, nor to conclude on a type of pleiotropy. In the 8-component model, $v$ will have a high probability of belonging to model (4), $X\&U$, that we choose to classify as "direct pleiotropy". In others words, this is an ingenious way of performing a fuzzy classification where categories are not strictly defined. This is why we preferred the more complex model, which can handle more subtle effects.

To account for the added complexity of the multi-effect components, we penalize each component with the prior probability $\omega_j$ for a variant to belong to the component. The more complex the component, the lower its prior, resulting in a greater penalty.

#### PRISM - Pleiotropic Relationships to Infer the SNP Model.

##### Part 4: Calculating posterior probabilities and scores for variants.

Parameters $\theta = i_x, i_y, \pi_x, \pi_y, h_x^2, h_y^2, q_x, q_y, \alpha_{x\to y}, \alpha_{y\to x}, \rho_{x\cap y}$ are estimated from the observed association summary statistics, using LHC-MR. Parameters $n_x, n_y, m$ are known, $h_u^2$ is a latent parameter fixed to $1$, $\pi_u$ is a latent parameter fixed to $10^{-5}$.

We chose $\pi_u = 10^{-5}$ based on simulations indicating that using a very low value for $\pi_u$ had no adverse effects. As shown in Supplementary Fig. 33, setting a very low value for $\pi_u$ significantly improved the precision in detecting direct variants, regardless of the actual polygenicity of the confounder.

Applying Bayes' theorem, the probability that a given variant $k$ belongs to the Gaussian component $j$ is determined by the following posterior probability:

$$\hat{P}(k \in j | \hat{\beta}_k^x, \hat{\beta}_k^y) = \frac{\phi(\hat{\beta}_k^x, \hat{\beta}_k^y | 0, \widehat{\mathbf{\Omega}}_\mathbf{j}) \hat{P}(k \in j)}{\sum_{i=0}^{7} \phi(\hat{\beta}_k^x, \hat{\beta}_k^y | 0, \widehat{\mathbf{\Omega}}_\mathbf{j}) \hat{P}(k \in j)}$$

Here, $\widehat{\mathbf{\Omega}}_\mathbf{j}$ denotes the variance-covariance matrix of Gaussian component $j$. Function $\phi$ represents the joint probability density function of the bivariate normal distribution, and $\hat{P}(k \in j) = \omega_j$ is the prior probability of variant $k$ to belong to component $j$, conditional on parameters $\theta$. These probabilities will be use to differentiate between direct and pleiotropic effects in Part 6.

The probabilities for variant $k$ to belong to the 8 Gaussian components are converted into scores, indicating whether $k$ has no effect (denoted $O$), or an effect on $X$, or on $Y$. No Effect: the score indicating no effect of variant $k$ on neither $X$ nor $Y$ corresponds to the probability of belonging to component (0): $\hat{S}_k^O = \hat{P}(k \in \{\text{component } 0\} | \hat{\beta}_k^x, \hat{\beta}_k^y)$

Effect on $X$: the score indicating an effect of variant $k$ on $X$ corresponds to the highest probability among the components that include an effect on $X$, specifically components (1), (4), (5) and (7): $\hat{S}_k^X = \hat{P}(k \in \{\text{component } 1{,}4{,}5{,}7\} | \hat{\beta}_k^x, \hat{\beta}_k^y)$

Effect on $Y$: the score indicating an effect of variant $k$ on $Y$ corresponds to the highest probability among components indicating an effect on $Y$, specifically components (3), (5), (6) and (7): $\hat{S}_k^Y = \hat{P}(k \in \{\text{component } 3{,}5{,}6{,}7\} | \hat{\beta}_k^x, \hat{\beta}_k^y)$

$\vec{S}_k = (\hat{S}_k^O \quad \hat{S}_k^X \quad \hat{S}_k^Y) \in \mathbb{R}^3$ is a vector of dimension 3 that contains these scores, for each variant $k$.

$$\hat{\mathbf{S}} = \begin{pmatrix} \hat{S}_1^O & \hat{S}_1^X & \hat{S}_1^Y \\ \hat{S}_2^O & \hat{S}_2^X & \hat{S}_2^Y \\ \vdots & \vdots & \vdots \\ \hat{S}_m^O & \hat{S}_m^X & \hat{S}_m^Y \end{pmatrix} \in \mathbb{R}^{m \times 3}$$

$\hat{\mathbf{S}}$ is a matrix of dimensions $m \times 3$ stemming from the concatenation of all $m$ $\vec{S}_k$ vectors.

**Part 5: Trait-wise workflow of PRISM and classification of the genetic variants.**

We detailed the process to obtain $\hat{\mathbf{S}}$ for two complex traits $X$ and $Y$, using their GWAS summary statistics. The idea of PRISM is to apply this process to a large number of traits $T$ pairwise. Consequently, $\frac{T(T-1)}{2}$ different $\hat{\mathbf{S}}$ matrices are obtained from the pairwise pipeline.

Then, for each variant $k$, we extract the scores $\hat{S}$ corresponding to each trait. For example, let us consider the variant-trait association of variant $k$ on a new trait $A$, included in PRISM workflow. We previously observed the effect of variant $k$ on trait $A$ in $T-1$ contexts, trait $A$ paired with all the other traits. So, we aggregate all previously calculated scores $\hat{S}_k^A$ and $\hat{S}_k^O$ from all $\hat{\mathbf{S}}$ matrices containing $A$. As a result, we obtain $\vec{E}_k^A = (\hat{S}_k^{A_1} \quad \hat{S}_k^{A_2} \quad \cdots \quad \hat{S}_k^{A_{T-1}}) \in \mathbb{R}^{T-1}$ and $\vec{E}_k^O = (\hat{S}_k^{O_1} \quad \hat{S}_k^{O_2} \quad \cdots \quad \hat{S}_k^{O_{T-1}}) \in \mathbb{R}^{T-1}$. Vector $\vec{E}_k^A$ is the collection of $T-1$ observations of the score of the very same variant $k$ to have a causal effect on trait $A$. Vector $\vec{E}_k^O$ is the collection of $T-1$ observations of the score of the very same variant $k$ to have no effect on trait $A$.

**Part 6: Statistical test of the variant-trait effect consistency.**

Next, we compare these values using a one-sided paired sign test. Specifically, we denote the sets of observed values for variant $k$ as $S_k^A$ and $S_k^O$. The null hypothesis of this sign test is that the median difference between $S_k^A$ and $S_k^O$ is zero, while the alternative hypothesis is that $S_k^A$ has a larger median than $S_k^O$.

By applying this test to all $k$ variants, we obtain one p-value per variant. Variant $k$ is considered significant if p-value$_k < \frac{5\times10^{-8}}{T-1}$. This threshold corresponds to a Bonferroni correction in addition to the usual GWAS significance threshold of $\mathrm{p} < 5 \times 10^{-8}$.

Then, to label significant variants for trait $A$ according to whether their effect is direct or pleiotropic, we follow a three-step procedure.

1) A significant variant is flagged with vertical pleiotropy on trait $A$ if any trait $B$ is causal to trait $A$, and $\hat{P}(k \in \{\text{component } 3\} | \hat{\beta}_k^x, \hat{\beta}_k^y) > \hat{P}(k \in \{\text{component } 0,1,2,4,5,6,7\} | \hat{\beta}_k^x, \hat{\beta}_k^y)$ in the causal inference model for pair $(A, B)$. This means that the probability that the genetic variant $k$ has an effect only on trait $B$ is higher than all other possibilities.

2) A significant variant is flagged with confounder pleiotropy on trait $A$ if, in at least one causal inference model involving trait $A$, $\hat{P}(k \in \{\text{component } 2\} | \hat{\beta}_k^x, \hat{\beta}_k^y) > \hat{P}(k \in \{\text{component } 0,1,3,4,5,6,7\} | \hat{\beta}_k^x, \hat{\beta}_k^y)$. This means that the probability that the genetic variant $k$ has an effect on $A$ only through a confounder is higher than all other possibilities.

3) All other significant variants are considered to have a direct effect on trait $A$.

The same procedure is applied to each trait.

**Part 7: Construction of the raw and deconvoluted variant networks.**

Finally, for each variant, we construct a raw network by creating a graph where nodes represent the variant and traits, and edges represent the relationships inferred from PRISM (direct, vertical, or confounder effects). The raw network is then pruned by removing vertical edges between the variant and specific traits, conditioned on other traits involved in vertical relationships. Indeed, although PRISM uses all the traits to test for consistency of effect, the labels themselves (direct, vertical, or confounder effects) are provided from a given pair of traits. Therefore, this step integrates labels on all traits and ensures that the causal pathways are accurately represented. Let us take a concrete example, we observe three vertical effects:

1) variant → Trait A → Trait B;

2) variant → Trait A → Trait C;

3) variant → Trait B → Trait C.

We remove edges that are null, conditioned on other traits. Since, the effect of the variant on Trait B can be explained by a mediated effect from Trait A, we remove the edge variant → Trait B (from vertical effects 1 and 3). In addition, since the effect of Trait A on Trait C can be explained by Trait B, we remove the edge Trait A → Trait C (from vertical effect 2). Therefore, the raw graph is pruned into variant → Trait A → Trait B → Trait C. Additionally, when a variant shows vertical effects on multiple traits through a common causal trait, we eliminate redundant edges. In our example, there are two edges variant → Trait A (from vertical effects 1 and 2), we simply remove one of the two.

A similar procedure has been employed in Mendelian randomization to prune trait causal graphs[35].

#### GWAS simulation

We constructed a pleiotropic network consisting of 60 simulated traits organized into four subnetworks with similar structure (See Fig. 6). Each subnetwork consisted of four groups of traits $A$, $B$, $C$, $D$ and $E$. $A$ connected directly to $B_3$ and $B_4$, and $B_1$ and $B_2$ had mutual causal influences while $D$ was causal to $A$. $B_4$ also connected to traits from the $C$ group, which made $B_4$ the most pivotal trait in our subnetwork. Traits from group $E$ had no causal relationships with any other traits. In addition a confounder was simulated for each pair of traits in the subnetwork. To link the subnetworks, a confounder was simulated between each pair of traits $B_4$ across subnetworks. 100,000 genetic variants were simulated for each of the 60 traits generating GWAS summary statistics. We tested 48 different scenarios, varying parameters for heritability, polygenicity, and causal relationships, as detailed in Supplementary Table 1. The sample size was set to $N = 361{,}194$, matching the sample size of UK Biobank GWAS round 2. To approximate a genome-wide model while maintaining computational feasibility, we opted for sets of $m = 100{,}000$ simulated variants. The standardized effects of all these variants on all the traits were simulated, taking into account the network relationships between the traits. First, for each trait, we randomly selected genetic variants to have a true direct effect on trait $X$ and on the confounders between trait $X$ and other traits. In addition to randomly selecting direct variants for each trait independently, we also randomly selected four causal variants to have direct effects on two traits, thereby introducing directed horizontal pleiotropy. Specifically, we introduced horizontal pleiotropy for the following trait pairs: $B_3$ and $C_2$, $C_3$ and $E_4$, $B_4$ and $E_5$, $E_2$ and $E_3$. This approach ensured a guaranteed direct effect of genetic variants on two traits, even in low polygenicity settings. Then, for all selected direct genetic variants, the true effects were drawn from a Gaussian distribution with parameters depending on the trait and the scenario. Specifically, true direct effects of all $m$ variants on trait $X$ were computed as:

$$\vec{\gamma}_x = \vec{\xi}_x \odot \vec{\kappa}_x$$

Here, $\xi_x \sim \mathcal{B}(m, \pi_x)$ consisted of binary elements (0 or 1) indicating the presence or absence of a genetic variant effect on trait $X$. The effect $\kappa_x \sim \mathcal{N}(0, \frac{h_x^2}{m\lambda_x})$ followed a Gaussian distribution with variance depending on the heritability of $X$, $h_x^2$, and on the total number of variants with effect on $X$, $m\lambda_x$. Similarly, the true confounder effects of all $m$ variants on all confounders $U$ were computed separately as:

$$\vec{\gamma}_u = \vec{\xi}_u \odot \vec{\kappa}_u$$

Similarly, $\xi_u \sim \mathcal{B}(m, \pi_u)$ consisted of binary elements (0 or 1) indicating the presence or absence of a genetic variant effect on $U$. The effect $\kappa_u \sim \mathcal{N}(0, \frac{h_u^2}{m\lambda_u})$ followed a Gaussian distribution with variance depending on the heritability of $U$, $h_u^2 = 1$, and on the total number of variants with effect on $U$, $m\lambda_u$.

Then, the true effects were propagated to the other $T$ traits through vertical and confounder pleiotropy. Specifically,

$$\vec{\beta}_x = \vec{\gamma}_x + \sum_{y=1}^{T} \left( (q_x + bq_y) \odot \vec{\gamma}_{u_{xy}} \right) + b\vec{\gamma}_y$$

Additionally, the effects were propagated according to the LD structure of each variant. For example, for variant $k$ in LD with all $J$ variants with coefficients denoted $r_j^2$:

$$B_k^x = \beta_k^x + \sum_{j=1}^{J} \beta_j^x \, r_j^2$$

Finally, an error term was added:

$$\overrightarrow{\widehat{\beta_x}} = \vec{B}_x + \mathcal{N}(\vec{O}_m, med(\frac{\sqrt{(1 - B_x^2)}}{\sqrt{N}})I_m)$$

This gives us, for all genetic variants, standardized effects $\overrightarrow{\widehat{\beta_x}}$ on all traits, and LD scores. The LD score of a variant is the sum of its LD with all variants (including itself). The LD structure that we used was derived from 1000 Genomes[36] LD data from chromosome 1. We chose to simulate small independent LD blocks, rather large LD blocks deemed computationally prohibitive for extensive simulations.
